# Dihydroartemisinin-piperaquine versus sulfadoxine-pyrimethamine for intermittent preventive treatment of malaria in pregnancy: a systematic review and individual participant data meta-analysis

**DOI:** 10.1101/2024.11.23.24315401

**Authors:** Michelle E. Roh, Julie Gutman, Maxwell Murphy, Jenny Hill, Mywayiwawo Madanitsa, Abel Kakuru, Hellen C. Barsosio, Simon Kariuki, John P.A. Lusingu, Frank Mosha, Richard Kajubi, Moses R. Kamya, Don Mathanga, Jobiba Chinkhumba, Miriam K. Laufer, Eulambius Mlugu, Appolinary A.R. Kamuhabwa, Eleni Aklillu, Omary Minzi, Roland Nnaemeka Okoro, Ado Danazumi Geidam, John David Ohieku, Meghna Desai, Prasanna Jagannathan, Grant Dorsey, Feiko O. ter Kuile

## Abstract

**Background:** High-grade *Plasmodium falciparum* resistance to sulfadoxine-pyrimethamine in East and Southern Africa has prompted numerous trials evaluating intermittent preventive treatment in pregnancy (IPTp) with dihydroartemisinin-piperaquine as an alternative to sulfadoxine-pyrimethamine.

**Methods:** We conducted individual participant data meta-analyses of randomised trials comparing IPTp with dihydroartemisinin-piperaquine to sulfadoxine-pyrimethamine on maternal, birth, and infant outcomes. We searched the WHO International Clinical Trials Registry Platform, ClinicalTrials.Gov, PubMed, and the Malaria in Pregnancy Consortium Library. Eligible trials enrolled HIV-uninfected pregnant women, followed participants to delivery, included participants with no prior IPTp use during the current pregnancy, and were conducted in areas with high-level parasite resistance to sulfadoxine-pyrimethamine (i.e., PfDHPS 540E≥90% and/or 581G>0%). Only singleton pregnancies were analysed. Meta-analyses used a two-stage approach: first, study-specific estimates were generated and then pooled using a random-effects model. Gravidity subgroup analyses were performed. Causal mediation analyses were used to investigate the maternal mechanisms underlying the effect of IPTp regimens on birth outcomes. The meta-analysis is registered in PROSPERO (CRD42020196127).

**Findings:** Of 85 screened records, six trials (one multi-country trial) contributed data on 6646 pregnancies. Compared to sulfadoxine-pyrimethamine, dihydroarteminsinin-piperaquine was associated with a 69% [95% CI: 45%–82%] lower incidence of clinical malaria during pregnancy, a 62% [37%– 77%] lower risk of placental parasitaemia, and a 17% [0%–31%] lower incidence of moderate maternal anaemia (Hb<9 g/dL). In contrast, sulfadoxine-pyrimethamine was associated with higher mean weekly maternal weight gain (34 grams/week [17–51]). There were no statistically significant differences in the composite adverse pregnancy outcome between the two IPTp regimens (RR=1·05 [95% CI: 0·92–1·19]; *I*^2^=48%), although the risk of small-for-gestational-age was 15% [3%–24%] lower in the sulfadoxine-pyrimethamine arm. Among multigravidae, participants of the sulfadoxine-pyrimethamine arm were 20% [8%–30%] and 35% [17%–49%] less likely to have stunted and underweight infants by two months compared to the dihydroartemisinin-piperaquine arm. Infant wasting by two months was 13% [3%–22%] lower in the sulfadoxine-pyrimethamine arm, regardless of gravidity. Mediation analyses indicated that 15% [0%–19%] of sulfadoxine-pyrimethamine’s superior effect on reducing small-for-gestational-age risk was mediated by its greater impact on gestational weight gain.

**Interpretation:** In areas of high *P. falciparum* sulfadoxine-pyrimethamine resistance, dihydroartemisinin-piperaquine is a more efficacious antimalarial than sulfadoxine-pyrimethamine. However, replacing sulfadoxine-pyrimethamine with dihydroartemisinin-piperaquine alone will not result in better maternal, birth, or infant outcomes. It could increase the risk of SGA, since much of the effect of sulfadoxine-pyrimethamine may be exerted through non-malarial mechanisms. Future research evaluating the alternative strategies for IPTp are needed, including with the combination of sulfadoxine-pyrimethamine and dihydroartemisinin-piperaquine.

**Funding:** This work was supported by the Bill and Melinda Gates Foundation and Eunice Kennedy Shriver National Institute of Child Health and Human Development.

**Research in context:** *Evidence before this study:* We searched the World Health Organization International Clinical Trials Registry Platform, ClinicalTrials.Gov, PubMed, and the Malaria in Pregnancy Consortium Library for randomised trials comparing intermittent preventive treatment in pregnancy (IPTp) with dihydroartemisinin-piperaquine to sulfadoxine-pyrimethamine, using the search term: (“intermittent preventive treatment” OR “IPTp”) AND ((“sulfadoxine-pyrimethamine” OR “sulphadoxine-pyrimethamine”) AND (“dihydroartemisinin-piperaquine”)). The initial search was conducted on July 30, 2020, and updated on September 24, 2024, without any restrictions on publication date, peer-review status, or language. We found eight studies, of which six were eligible for inclusion in this meta-analysis. Two previous meta-analyses had been conducted: a 2018 review by Desai et al that included the first two trials, and a subsequent pooled analysis by Roh et al in 2020 that included the first three trials and focused disentangling the antimalarial and non-malarial effects of sulfadoxine-pyrimethamine versus dihydroartemisinin-piperaquine. These reviews highlighted the superior antimalarial efficacy of dihydroartemisinin-piperaquine compared to sulfadoxine-pyrimethamine, but also suggested the potential superior non-malarial benefits of sulfadoxine-pyrimethamine. A recent meta-analysis by Muthoka et al evaluated the safety of IPTp with dihydroartemisinin-piperaquine in pregnancy. However, an updated meta-analysis comparing the efficacy of all currently completed trials of IPTp with dihydroartemisinin-piperaquine versus sulfadoxine-pyrimethamine has not been conducted.

*Added value of this study:* This study represents the first and only meta-analysis using individual participant data from all six available trials conducted in areas with high sulfadoxine-pyrimethamine resistance. By pooling data from 6646 pregnancies across multiple African countries, we were able to conduct a more robust and nuanced analysis comparing the efficacy of dihydroartemisinin-piperaquine to sulfadoxine-pyrimethamine for IPTp. Our findings confirm the superior antimalarial efficacy of dihydroartemisinin-piperaquine but also reveal that sulfadoxine-pyrimethamine is associated with better birth and infant outcomes, particularly in reducing the risk of small-for-gestational age and infant malnutrition. This meta-analysis provides strong evidence for the existence of non-malarial benefits of sulfadoxine-pyrimethamine in pregnancy, which appear to outweigh its reduced antimalarial efficacy in terms of pregnancy outcomes, even in areas of high resistance.

*Implications of all the available evidence:* Based on our comprehensive analysis, we recommend against switching from sulfadoxine-pyrimethamine to dihydroartemisinin-piperaquine for IPTp, even in areas with very high sulfadoxine-pyrimethamine resistance. Such a change would likely reduce gestational weight gain, lower mean newborn birthweights, increased risk of SGA, and poor early infant growth. Instead, we recommend further studies combining sulfadoxine-pyrimethamine with dihydroartemisinin-piperaquine (or another potent malaria strategy) to harness the non-malarial benefits of sulfadoxine-pyrimethamine and target the malaria-associated causes of adverse pregnancy outcomes. Additionally, more research is needed to better understand the mechanisms underlying the non-malarial effects of these drugs, including their direct antimicrobial activity, effects on gut and vaginal health, and/or influence on maternal systemic inflammation. This research is crucial for optimising malaria prevention strategies in pregnancy and improving maternal and neonatal outcomes in malaria-endemic regions.

## Introduction

In sub-Saharan Africa, malaria infection during pregnancy poses substantial risks for both the mother and foetus. In moderate-to-high transmission settings, infection with the *Plasmodium falciparum* parasite is associated with maternal anaemia, miscarriage, stillbirth, preterm birth (PTB), intrauterine growth restriction, low birthweight (LBW), and neonatal mortality.^1^ In 2022, nearly 13 million pregnant women in the WHO African region, which accounts for 94% of *P. falciparum* cases, were exposed to malaria.^2^ To avert the consequences of malaria during pregnancy, the World Health Organization (WHO) recommends intermittent preventive treatment of malaria in pregnancy (IPTp).^3^ This strategy involves administering full treatment courses of a long-acting antimalarial starting the second trimester of pregnancy up to delivery, with doses given at least one month apart. Currently, 35 African countries have adopted IPTp into their national malaria policy.^2^

Since its initial recommendation in 1998, sulfadoxine-pyrimethamine has been the only antimalarial recommended for IPTp. Over the past 30 years, its widespread use has led to the emergence of parasite resistance to sulfadoxine-pyrimethamine, particularly in East and Southern Africa.^4,5^ Concerns over the limited antimalarial efficacy of sulfadoxine-pyrimethamine has prompted researchers to evaluate alternative regimens for IPTp. Of the numerous antimalarial combinations studied, dihydroartemisinin-piperaquine has been the most promising candidate to replace sulfadoxine-pyrimethamine due to its excellent efficacy, long prophylactic period, and safety profile for pregnant women. A 2018 meta-analysis^6^ of the first two trials comparing dihydroartemisinin-piperaquine to sulfadoxine-pyrimethamine^7,8^ found that dihydroartemisinin-piperaquine was associated with a significantly lower incidence of clinical malaria, placental malaria, maternal anaemia, and foetal loss.^6^ However, impacts on LBW, PTB, and small-for-gestational age (SGA) did not statistically significantly differ between regimens. Thus, the WHO recommended further research to determine whether dihydroartemisinin-piperaquine could be a viable replacement for sulfadoxine-pyrimethamine.^9^

Since then, four additional trials from Uganda, Kenya, Malawi, Tanzania, and Nigeria have been published,^10–13^ three of which were conducted in areas with high *P. falciparum* resistance to sulfadoxine-pyrimethamine.^10–12^ While results from these trials consistently demonstrated dihydroartemisinin-piperaquine’s superior effect on malaria outcomes, findings were mixed regarding its impact on birth outcomes. Moreover, some trials showed that compared to dihydroartemisinin-piperaquine, sulfadoxine-pyrimethamine exhibited a greater effect on mean birthweight,^12,14^ mean maternal mid-upper arm circumference (MUAC), and gestational weight gain (GWG).^12^ However, these outcomes were not consistently reported across trials, highlighting the need for further assessment.

A recent meta-analysis evaluated the safety of IPTp with dihydroartemisinin-piperaquine in pregnancy.^15^ The aim of the current systematic review and meta-analysis was to provide an updated and comprehensive review of trials conducted in areas of high *P. falciparum* resistance that compared the efficacy of IPTp with dihydroartemisinin-piperaquine to sulfadoxine-pyrimethamine across maternal, birth, and infant outcomes.

## Methods

### Search strategy and selection criteria

The systematic review and meta-analysis were conducted in accordance with the Preferred Reporting Items for Systematic Reviews and Meta-Analyses Statement (**Appendix 1, pp 3–6**). We searched the WHO International Clinical Trials Registry Platform, ClinicalTrials.Gov, PubMed, and the Malaria in Pregnancy Consortium Library database for original articles, abstracts, reports, or protocols using the search term: (“intermittent preventive treatment” OR “IPTp”) AND ((“sulfadoxine-pyrimethamine” OR “sulphadoxine-pyrimethamine”) AND (“dihydroartemisinin-piperaquine”)). The search was conducted on July 30, 2020, and updated on September 24, 2024, without restrictions to publication date, peer-review status, or language.

Trials were eligible if they met the following inclusion criteria: randomised HIV-uninfected pregnant women to either IPTp with dihydroartemisinin-piperaquine or sulfadoxine-pyrimethamine; followed participants to delivery to assess malaria and delivery outcomes; enrolled women with no prior use of IPTp during their current pregnancy; and were conducted in areas with high-level parasite resistance to sulfadoxine-pyrimethamine (*P. falciparum* dihydropteroate synthase (PfDHPS) 540E mutation prevalence ≥90% and/or 581G mutation >0%). Data on PfDHPS 540E and 581G prevalence were obtained directly from studies or nearby sites if unavailable. Treatment arms were excluded if dosing schedules differed between arms and/or study drugs were co-administered with another intervention (e.g., azithromycin or metronidazole). Non-singleton pregnancies were excluded from our analyses.

### Data extraction and quality assessment

Screening was conducted by two independent reviewers (MER and JG). Any uncertainties or discrepancies were resolved through discussion with a third reviewer (FtOK) or by contacting trial authors for clarification. For each eligible trial, chief investigators were invited to collaborate and contribute their individual participant data. Up to three attempts were made to contact authors to participate in the meta-analysis. A description of the available study outcomes from each study is provided in **Appendix 2** (**pp 7–10**). The risk of bias was assessed using The Cochrane Risk of Bias tool for randomised trials version 2 (RoB2).^16^ The meta-analysis is registered in PROSPERO (CRD42020196127).

### Study endpoints

Definitions of study endpoints are provided in **Appendix 3** (**pp 11–13**). The primary endpoint was defined as the risk of any adverse pregnancy outcome, a composite outcome of either miscarriage (foetal loss <28 gestational weeks), stillbirth (foetal loss ≥28 gestational weeks), PTB (delivery <37 gestational weeks), SGA (birthweight <10^th^ percentile for gestational age using INTERGROWTH-21^st^ standards^17^); LBW (birthweight <2500 grams), and neonatal loss (newborn death within the first 28 days of life). PTB, SGA, LBW, and neonatal loss were only assessed among live births. Secondary endpoints included the individual components of the primary outcome; mean birthweight in grams, gestational age at birth in weeks, birthweight-for-gestational age (BWGA) z-scores using INTERGROWTH 21^st^ standards^17^; incidence of clinical malaria during pregnancy; measures of placental malaria; maternal peripheral malaria infection at delivery; measures of maternal anaemia during pregnancy; maternal MUAC at delivery; and GWG per week in grams.

Post-hoc analyses were performed to evaluate differences in infant anthropometric measures between IPTp regimens. Infant outcomes included cumulative incidence of stunting, wasting, and underweight measured from birth to approximately two months of life, and mean differences in length-for-age, weight-for-age, and weight-for-length z-scores at approximately two months of life. Z-scores were calculated according to age and sex based on the 2006 WHO Child Growth Standards^18^ using the zscorer R package.^19^ Stunting, underweight, and wasting were defined as <2 standard deviations below median WHO standards for length-for-age, weight-for-age, and weight-for-length z-scores, respectively.

### Statistical analysis

The study employed a two-stage, individual participant data meta-analysis. In the first stage, individual-level data were analysed to generate study-specific estimates. In the second stage, study-specific estimates were pooled to generate summary estimates using restricted maximum likelihood estimation random-effects models. Between-study heterogeneity was assessed using the *I*^2^ statistic. Prediction intervals were reported for each outcome. Meta-analyses were conducted using the meta R package;^20^ forest plots were generated using the metafor R package.^21^

Study-specific estimates were computed using unadjusted models, except for maternal weight gain and MUAC outcomes which adjusted for enrolment values. Binary outcomes were modelled using log-binomial regression to estimate risk ratios. Modified Poisson regression with robust standard errors^22^ was used if log-binomial models did not converge. Continuous outcomes were modelled using linear regression to compute mean differences. Incidence rate ratios were estimated using Poisson regression with an offset term of the number of days at-risk between the first day study drugs were given to the last day of the pregnancy period (for maternal outcomes). For all outcomes, we reported subgroup analyses by gravidity (primi-versus multi-gravidae). P-values testing for subgroup differences (p_subgroup_) were based on comparing differences in the Q statistic.

Mediation analyses were conducted to examine the extent to which differences in birth outcomes between IPTp regimens were influenced by maternal outcomes that statistically significantly differed between arms. Mediation analyses were carried out following a potential outcomes framework and used targeted minimum loss estimation to estimate natural indirect (mediated) and direct (non-mediated) effects. Separate analyses were conducted for each mediator using the medoutcon R package.^23^ Further details of the analytic approach are described in **Appendix 4** (**pp 14–15**). All analyses were conducted using Stata 16.1 (StataCorp, College Station, TX, USA) and R (version 4.3.2; R Project for Statistical Computing; http://www.r-project.org/).

## Results

### Description of studies

Our search yielded 154 records (**Figure 1**); one additional study (PACTR201701001982152) was found outside the search strategy. After removing duplicates, 85 records were screened, identifying eight randomised controlled trials. All but one study (PACTR201808204807776) provided individual-level data. One study from Nigeria (Okoro 2023)^13^ was excluded from the meta-analysis due to its location in an area with low sulfadoxine-pyrimethamine resistance (PfDHPS 437G mutation prevalence=35% with no evidence of the 540E or 581G mutation^24^); results from this trial were presented separately in **Appendix 9** (**pp 54–58**). The remaining six trials (five published^7,8,10–12^ and one unpublished^25^) were conducted in Kenya (n=2), Malawi (n=2), Uganda (n=2), and Tanzania (n=2), where PfDHPS 540E and 581G mutation prevalence ranged from 52%–99% and 0%–40%, respectively (**Appendix 2, pp 7–8**). The Madanitsa 2023 trial^12^ was conducted in three countries (Kenya, Malawi, and Tanzania); thus, country-specific estimates were reported separately and treated as three distinct studies, bringing the total to eight studies. Individual participant data were obtained from 6723 participants. After excluding 77 non-singleton pregnancies, the final analytic sample comprised 6646 singleton pregnancies. Six of the eight studies were scored as having a low risk of bias and two as having some concerns as ultrasound was not used for gestational age dating (**Appendix 5, p 16**).

**Figure 1.**
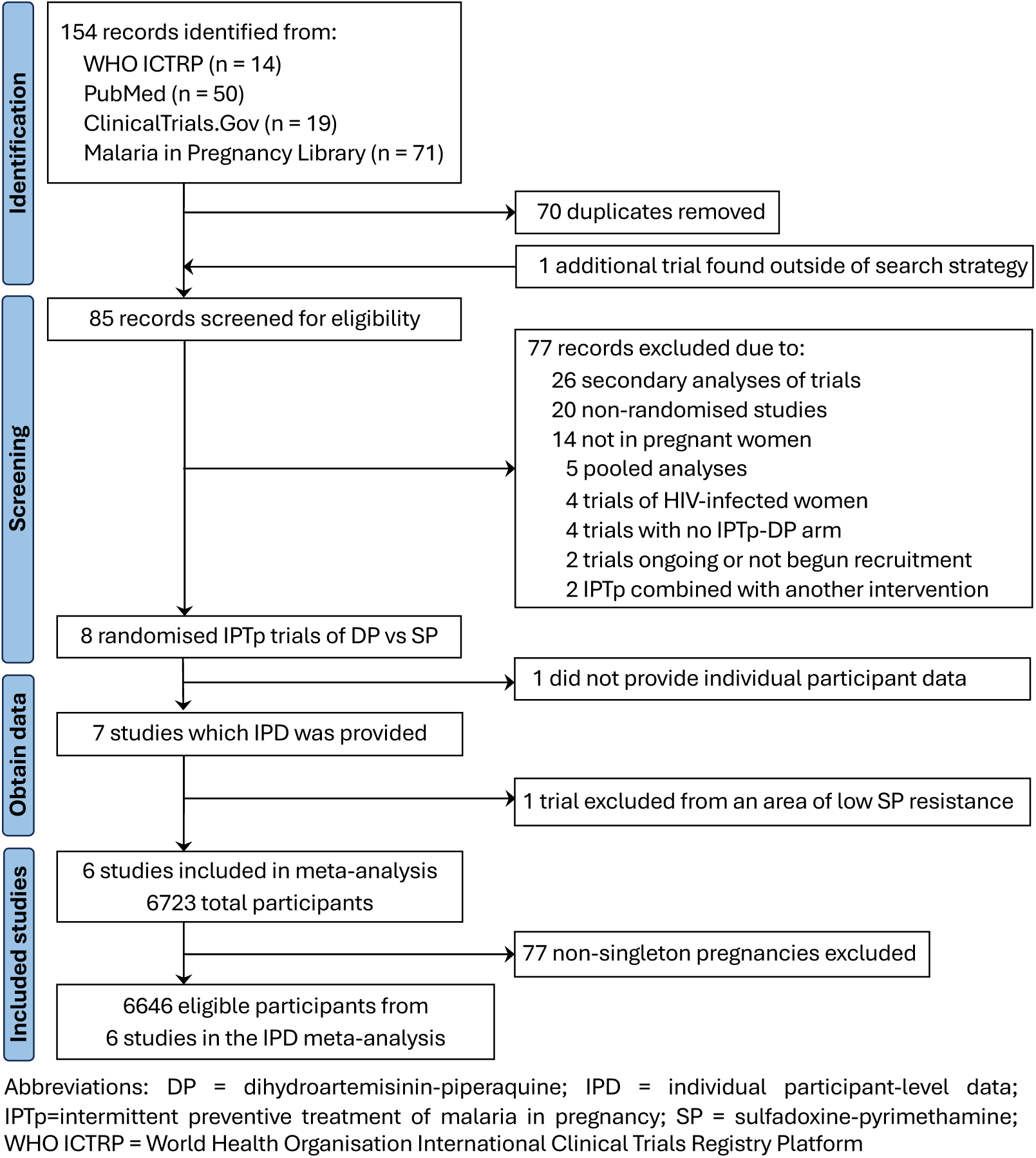
PRISMA flow diagram of included studies and participants.

Across studies, enrolment characteristics were balanced between arms (**Appendix 6, pp 17–24**). LAMP/PCR positivity at enrolment ranged from 11%–81% across studies. The median number of IPTp courses was 4 [interquartile range (IQR): 3–5] in studies that administered IPTp every four weeks (n=6 studies^10–12,25^), 2 [IQR: 2–3] in those that administered IPTp every antenatal care visit spaced ≥1 month apart (n=1 study^7^), and 3 [2–3] in those that administered every eight weeks (n=1 study^8^). In all trials, participants received insecticide-treated nets at enrolment.

### Birth outcomes

Data on the primary endpoint (a composite of any adverse pregnancy outcome) was available from all eight studies (N=6153 pregnancies). Across studies, the risk of experiencing any adverse pregnancy outcome ranged from 16%–33% in the sulfadoxine-pyrimethamine arm and 14%–34% in the dihydroartemisinin-piperaquine arm. The pooled RR comparing the risk of any adverse pregnancy outcome between arms was 1·05 [95% CI: 0·92–1·19] (p=0·50). The *I*^2^ statistic was 48%, indicating moderate between-study heterogeneity. Pooled RRs of the individual components of the primary outcome showed no statistically significant differences in the risk of foetal loss, PTB, LBW, or neonatal death between arms (**Figure 2A**; **Appendix 7, pp 25–30**). However, the risk of SGA was statistically significantly higher in the dihydroartemisinin-piperaquine arm compared to sulfadoxine-pyrimethamine (pooled RR=1·17 [95% CI: 1·03–1·32]; p=0·016; *I*^2^=3%). This effect was mainly seen in multigravidae (pooled RR_multi_=1·28 [95% CI: 1·10–1·49] versus pooled RR_primi_=1·09 [95% CI: 0·92–1·30]), though testing of subgroup differences did not reach statistical significance (p_subgroup_=0·18). The directions for the overall and gravidity subgroup analyses were similar for LBW, except for the Mlugu 2021 study, where LBW risk was statistically significantly lower in the dihydroartemisinin-piperaquine arm (RR=0·51 [95% CI: 0·31–0·84]) (**Appendix 7, p 29**).

**Figure 2.**
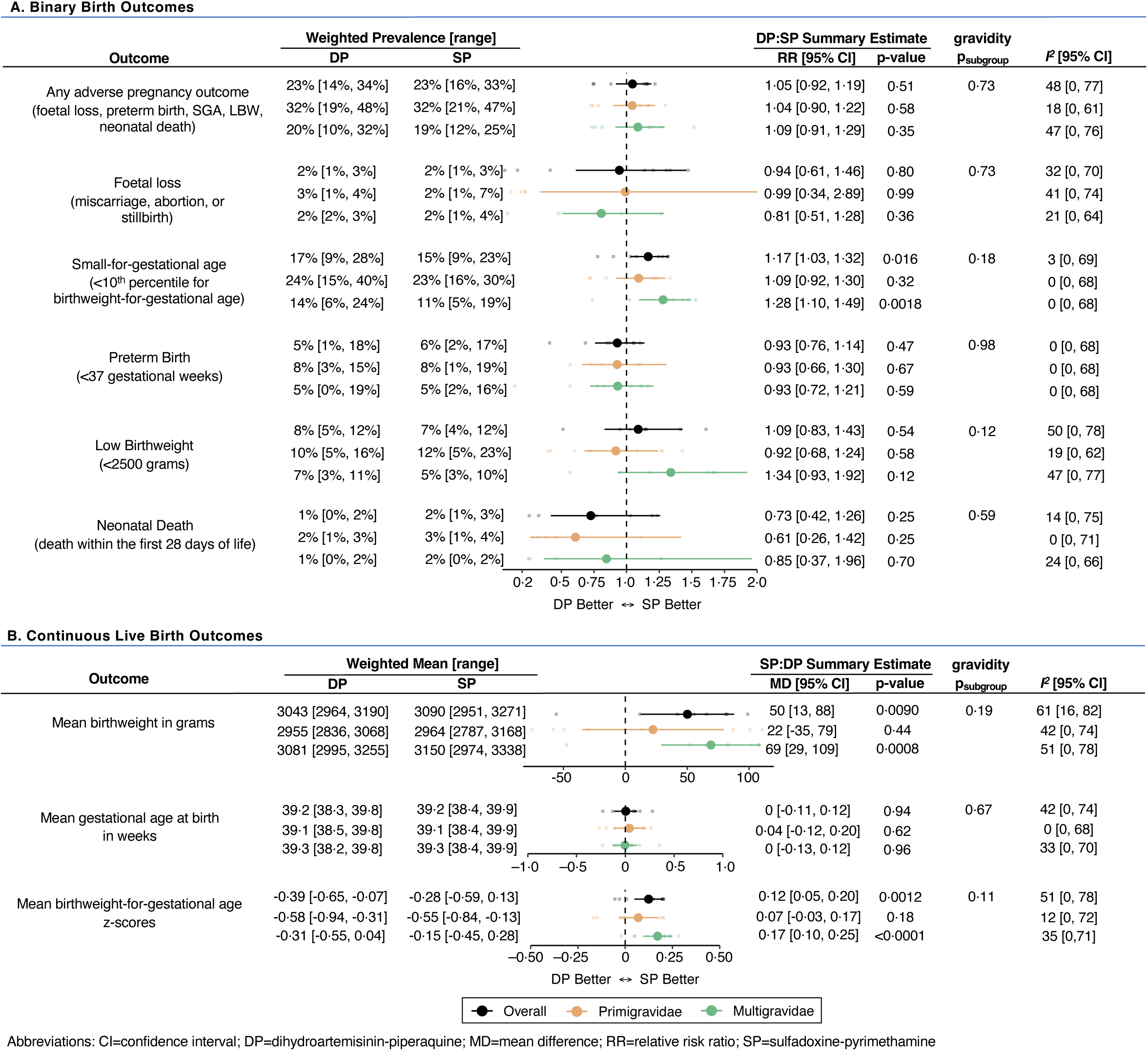
Forest plot comparing binary (A) and continuous live (B) birth outcomes between IPTp regimens. All estimates reflect unadjusted differences between arms. Weighted prevalences and means for each outcome were calculated using a restricted maximum likelihood random-effects model.

Pooled estimates of continuous live birth outcomes showed that compared to dihydroartemisinin-piperaquine, sulfadoxine-pyrimethamine was associated with higher mean newborn birthweight (mean difference (MD)=50 grams [95% CI: 13–88]; p=0·0090, *I*^2^=61%) and BWGA z-scores (MD=0·12 [95% CI: 0·05–0·20]; p=0·0010, *I*^2^=51%), but not gestational age at birth (MD=0 weeks [95% CI: –0·11–0·12]; p=0·94; *I*^2^=42%) (**Figure 2B**; **Appendix 7, pp 31–33**). While study-specific estimates varied for primigravidae, the direction of effect estimates for multigravidae was consistent in all studies except for the Mlugu 2021 study, which found newborn birthweight and gestational age at birth was higher in the dihydroartemisinin-piperaquine arm, regardless of gravidity.

### Maternal outcomes

All studies evaluated the following malaria endpoints: incidence of clinical malaria during pregnancy, presence of parasitaemia in placental tissue and/or blood, and peripheral parasitaemia. Pooled estimates of malaria endpoints showed that compared to sulfadoxine-pyrimethamine, dihydroartemisinin-piperaquine was associated with a 69% [95% CI: 45–82] lower risk of clinical malaria, 34% [95% CI: 20–45] lower risk of placental pigmentation (past infection), 62% [95% CI: 37–77] lower risk of placental parasitaemia at delivery (active or chronic infection), and 61% [95% CI: 45–73] lower risk of maternal peripheral malaria at delivery (**Figure 3**; **Appendix 7, pp 34–38**). While substantial heterogeneity was observed between studies (range of *I*^2^ values: 64%– 81%), estimates generally favoured dihydroartemisinin-piperaquine for malaria prevention. Subgroup analyses revealed that although the risks of clinical malaria and active placental malaria infection at delivery were nearly two-fold higher in primigravidae, effect sizes were similar between gravidity subgroups, except for preventing placental pigmentation (RR_primi_=0·85 [95% CI: 0·74–0·98] versus RR_multi_=0·53 [95% CI: 0·38–0·74]; p_subgroup_=0·011). In addition to its superior effects on malaria prevention, dihydroartemisinin-piperaquine was associated with a lower risk of moderate anaemia (pooled RR=0·83 [95% CI: 0·69–1·00]; p=0·050; *I*^2^=41%) (**Figure 4A**; **Appendix 7, p 40**).

**Figure 3.**
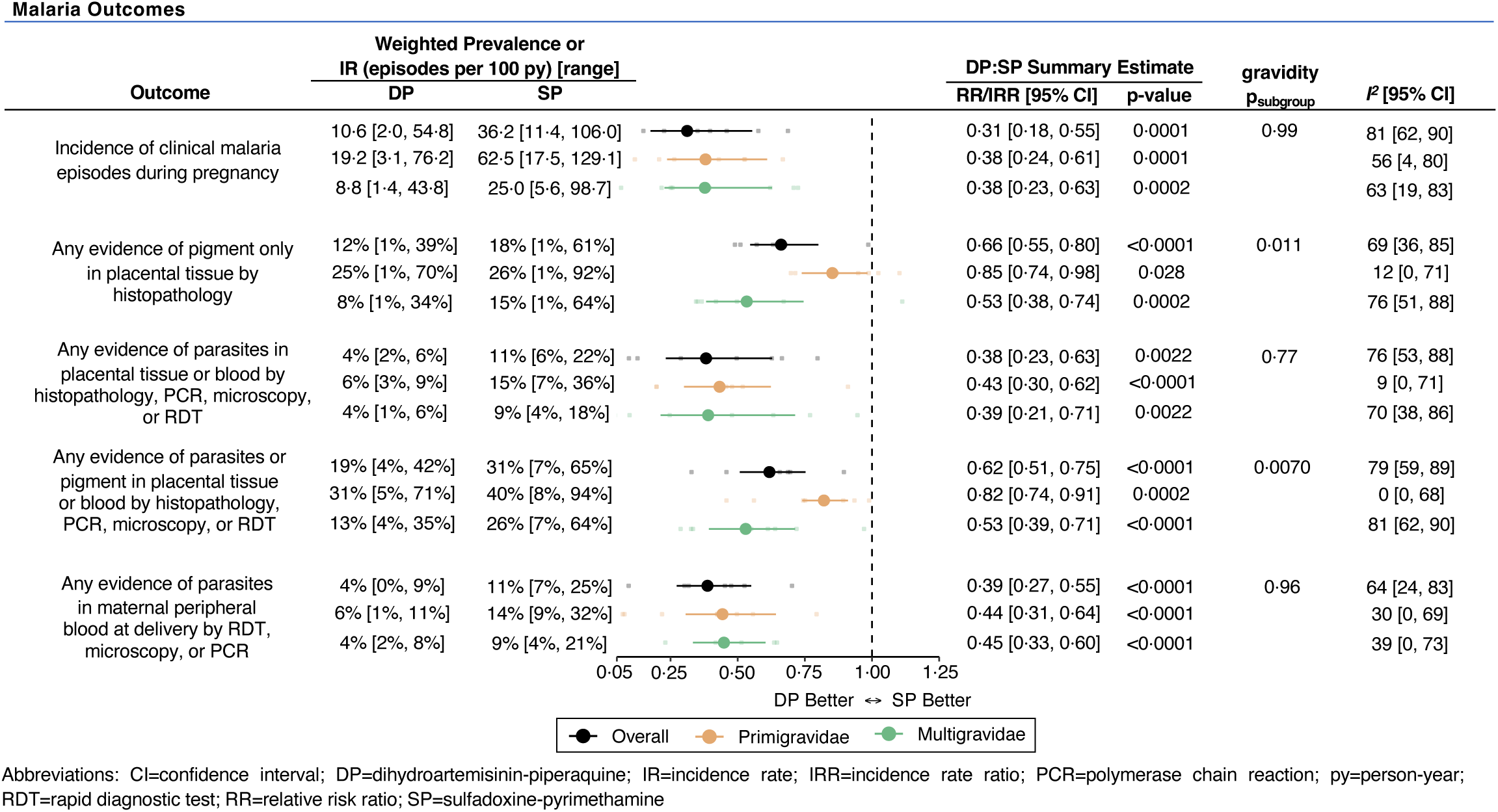
Forest plot comparing malaria outcomes between IPTp regimens. All estimates reflect unadjusted differences between arms. Weighted prevalence and incidence rates for each outcome were calculated using a restricted maximum likelihood random-effects model.

**Figure 4.**
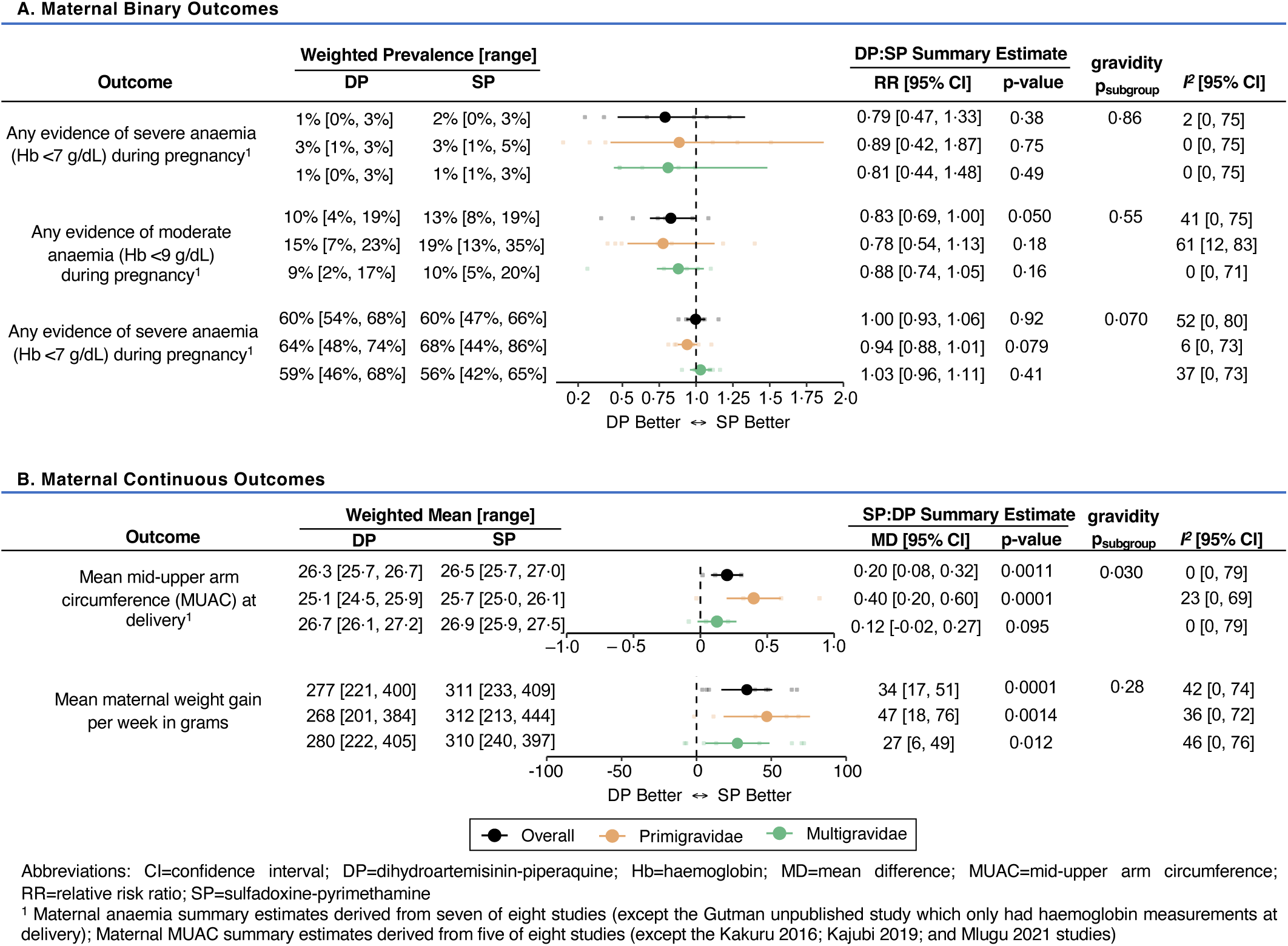
Forest plot comparing binary (A) and continuous (B) maternal outcomes between IPTp regimens. All estimates reflect unadjusted differences between arms, except for mean MUAC and gestational weight gain, which adjusted for enrolment values. Weighted prevalence and means for each outcome were calculated using a restricted maximum likelihood random-effects model.

Compared to dihydroartemisinin-piperaquine, sulfadoxine-pyrimethamine was associated with higher mean maternal MUAC at delivery (pooled MD=0·20 cm [95% CI: 0·08–0·32]; p=0·0011; *I*^2^=0%) with the greatest difference in primigravidae (MD_primi_=0·40 cm [95% CI: 0·20–0·60] versus MD_multi_=0·12 cm [95% CI: –0·02–0·27]; p_subgroup_=0·030) (**Figure 4B**; **Appendix 7, p 42**). Sulfadoxine-pyrimethamine was also associated with greater GWG (pooled MD=34 grams/week [95% CI: 17–51]; p=0·0001; *I*^2^=42%), with similar effects in primigravidae (MD_primi_=47 grams/week [95% CI: 18–76]) and multigravidae (MD_multi_=27 grams/week [95% CI: 6–49]; p_subgroup_=0·28).

### Infant anthropometric outcomes

Post-hoc analyses from seven of the eight studies showed that among multigravidae, the risks of stunting and underweight among infants followed from birth up to approximately two months of life were 1·25 [95% CI: 1⋅09–1⋅43] and 1·54 [95% CI: 1⋅20–1⋅98] times higher in mothers randomised to dihydroartemisinin-piperaquine arm compared to sulfadoxine-pyrimethamine (**Figure 5A**; **Appendix 7, pp 44–46**). The risk of early wasting was higher in infants born to mothers randomised to dihydroartemisinin-piperaquine arm, regardless of gravidity (RR=1·15 [95% CI: 1·03–1·29]). At approximately two months of life, mean infant length-for-age and weight-for-age z-scores were higher in the sulfadoxine-pyrimethamine arm. However, mean weight-for-length z-scores were higher in the dihydroartemisinin-piperaquine arm, especially among multigravidae (MD_multi_=0·13 [95% CI: 0·02–0·25]) (**Figure 5B**; **Appendix 7, pp 47**–**49**).

**Figure 5.**
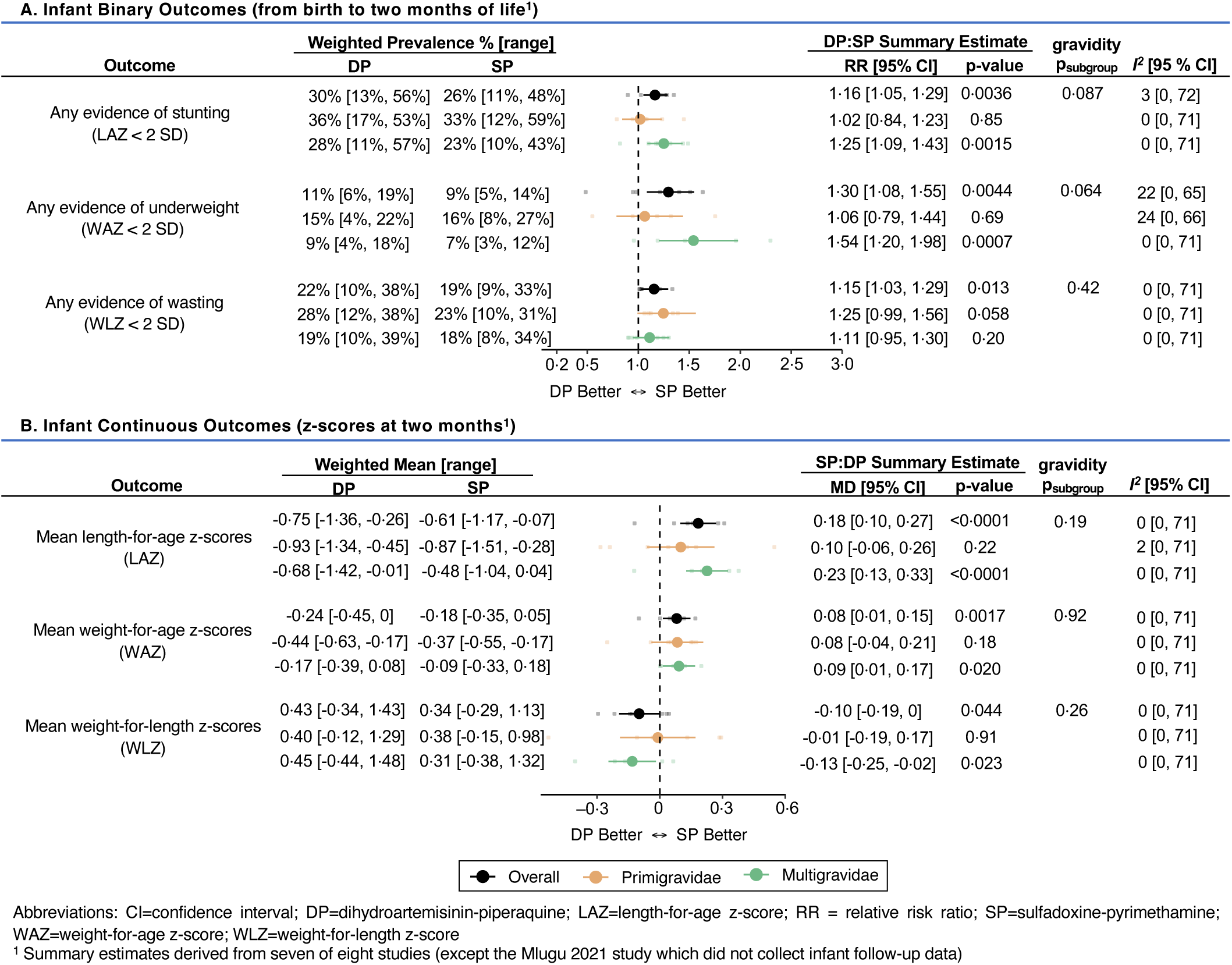
Forest plot comparing binary (A) and continuous (B) infant outcomes between IPTp regimens. All estimates reflect unadjusted differences between arms. Weighted prevalence and means for each outcome were calculated using a restricted maximum likelihood random-effects model.

### Mediation analyses

Given sulfadoxine-pyrimethamine’s greater benefit on newborn birthweight (but not gestational age), we conducted mediation analyses to examine the extent to which differences in BWGA z-scores between regimens were mediated by variations in the incidence of clinical malaria, placental malaria (defined as any evidence of parasites or pigment), GWG, and maternal MUAC (**Appendix 8, pp 50**–**53**). Pooled estimates showed that dihydroartemisinin-piperaquine’s superior effect on preventing placental malaria infection contributed a relatively small proportion to improving BWGA z-scores, especially compared to sulfadoxine’s superior ‘non-malarial’ effect (dihydroartemisinin-piperaquine’s indirect, “antimalarial” effect=0·01 [95% CI: 0–0·02] versus sulfadoxine-pyrimethamine’s direct, “non-malarial” effect=0·15 [95% CI: 0·07–0·23]). Dihydroartemisinin-piperaquine’s antimalarial effect was greatest in the Kajubi 2019 study (indirect effect=0·10 [95% CI: 0·03–0·17]), where malaria burden was exceptionally high (81% of women had detectable parasitaemia by PCR at enrolment). Similar associations were seen when incidence of clinical malaria during pregnancy was used as the mediating variable (**Appendix 8, p 50**).

Notably, we found that 15% of sulfadoxine-pyrimethamine’s superior effects on BWGA z-scores was mediated by its superior effects on GWG (pooled indirect effect=0·02 [95% CI: 0–0·04] and pooled direct effect = 0·11 [95% CI: 0·05–0·17]). Of the five studies that measured MUAC at delivery, summary estimates showed differences in maternal MUAC mediated a relatively small proportion (2%) of the superior effect of sulfadoxine-pyrimethamine on BWGA z-scores (pooled indirect effect=0·003 [95% CI: –0·004–0·010] and pooled direct effect=0·16 [95% CI: 0·11– 0·22]).

## Discussion

In this comprehensive meta-analysis of six randomised controlled trials of IPTp, we found that in areas of high *P. falciparum* resistance to sulfadoxine-pyrimethamine, dihydroartemisinin-piperaquine was associated with markedly lower risks of clinical, placental, and peripheral malaria infection during pregnancy. Despite superior malaria prevention, summary estimates showed that the composite risk of adverse pregnancy outcomes did not differ between regimens. Analyses of the individual components of the composite outcome revealed that infants born to women randomised to sulfadoxine-pyrimethamine had a lower risk of being SGA and had higher mean birthweights, particularly among multigravidae. No statistically significant differences were seen in foetal loss, neonatal death, or gestational age at birth, suggesting that the superior effect of sulfadoxine-pyrimethamine is likely through improving foetal growth rather than premature delivery. Our findings were generally consistent across studies, except for the Mlugu 2021 study, where dihydroartemisinin-piperaquine was associated with a lower risk of LBW and PTB than sulfadoxine-pyrimethamine. Further analyses of maternal outcomes showed that compared to dihydroartemisinin-piperaquine, sulfadoxine-pyrimethamine was associated with modestly higher maternal MUAC and GWG, while dihydroartemisinin-piperaquine was associated with a lower risk of moderate anaemia. Interestingly, the benefits of sulfadoxine-pyrimethamine extended into early infancy whereby infants born to women in this group were less likely to experience stunting, underweight, or wasting in the first two months of life—a critical period with limited interventions for promoting growth.^26^ Collectively, these findings support the continued use of sulfadoxine-pyrimethamine for IPTp but suggest that in areas of high *P. falciparum* sulfadoxine-pyrimethamine resistance, additional interventions are needed to prevent malaria.

Our gravidity subgroup analyses revealed primigravidae and their infants consistently experienced poorer health outcomes than multigravidae. In primigravidae, the comparison of sulfadoxine-pyrimethamine to dihydroartemisinin-piperaquine for SGA risk was closer to the null than in multigravidae. This weaker effect is likely attributable to the stronger impact of dihydroartemisinin-piperaquine in preventing placental malaria, as primigravidae have not yet acquired parity-dependent malarial-immunity.^27^ Despite this, we recommend against gravidity-dependent approaches to IPTp (i.e., adding dihydroartemisinin-piperaquine or another malaria prevention approach to sulfadoxine-pyrimethamine for primigravidae only), as protecting against placental malaria in the first pregnancy could hinder immunity acquisition and increase risks in subsequent pregnancies. Additionally, a gravidity-specific strategy would be logistically more complex to implement.

Our mediation analyses confirm results from prior studies demonstrating sulfadoxine-pyrimethamine’s potent ‘non-malarial’ effect^14^ and its impacts on increasing GWG and maternal MUAC.^12,28^ While dihydroartemisinin-piperaquine exhibited superior effects on preventing placental malaria, its contribution to increasing BWGA was relatively modest compared to sulfadoxine-pyrimethamine’s non-malarial effect, except in the Kajubi 2019 study, where malaria burden was especially high. Importantly, our GWG results support earlier findings from a secondary analysis of the Gutman unpublished trial^28^ and the Madanitsa 2023 trial.^12^ Given its broad-spectrum activity, the precise mechanisms by which sulfadoxine-pyrimethamine enhances foetal and infant growth (either through or independent of GWG) likely involve multiple pathways. Several studies have demonstrated these mechanisms may include: impact on enteroaggregative *Escherichia coli*,^28^ febrile respiratory illnesses,^29^ maternal nutrient absorption,^30^ and changes in maternal inflammatory responses.^26^ In contrast, sulfadoxine-pyrimethamine’s non-malarial effects were absent in the Kakuru 2016^8^ and Mlugu 2021^11^ trials, which may suggest that these mechanisms were less prominent in these trial populations or that dihydroartemisinin-piperaquine could provide comparable non-malarial benefits, although other explanations are possible. Notably, IPTp dosing in the Kakuru 2016 trial^8^ was less frequent (every eight weeks), compared to most other trials, suggesting that the non-malarial effects may follow a dose-response relationship. Further studies on the effects of these regimens on non-malarial infections, the gut and vaginal microbiome, and maternal inflammation may offer deeper insights.

This meta-analysis had several strengths, including its diverse evaluation across multiple countries, comprehensive assessment of maternal, birth, and infant outcomes, and inclusion of mediation analyses and gravidity subgroup analyses, which provided valuable and nuanced insights into the antimalarial and non-malarial benefits of IPTp. However, certain limitations should be considered. First, the small number of included trials restricted our ability to conduct meta-regression analyses and assess for small-study effects or publication bias. Moreover, the reported *I*^2^ statistics, which can be biased with a small number of studies,^31^ should be interpreted cautiously. We were also likely underpowered to detect true differences between gravidity subgroups. Second, as with all meta-analyses, larger studies had a greater influence on the summary estimate, which may limit the generalizability of our findings, particularly in the presence of substantial heterogeneity. Third, separate mediation analyses were conducted for each mediator, limiting our understanding of how these mediators function independently or in combination. Fourth, our mediation estimates may be subject to unmeasured mediator-outcome confounding and measurement error and should be interpreted cautiously. Finally, infant outcomes were only assessed up to two months of life and further research is needed to understand longer-term impacts.

In conclusion, our meta-analyses showed that, in areas with high *P. falciparum* resistance to sulfadoxine-pyrimethamine, dihydroartemisinin-piperaquine was more efficacious in preventing malaria and maternal anaemia. However, suppose the goal of IPTp is to improve overall maternal, foetal, and infant health outcomes. In that case, replacing sulfadoxine-pyrimethamine with dihydroartemisinin-piperaquine is unlikely to be beneficial and could increase the risk of SGA and poor infant growth early in life. This may be because sulfadoxine-pyrimethamine offers ‘non-malarial’ benefits on maternal nutrition and foetal growth that, in some settings, may outweigh the antimalarial benefits of dihydroartemisinin-piperaquine. Therefore, future studies should evaluate the combined IPTp regimen of sulfadoxine-pyrimethamine and dihydroartemisinin-piperaquine (or another effective malaria prevention strategy) and investigate the ‘non-malarial’ mechanisms by which these regimens affect maternal and infant outcomes.

## Article information

### Disclaimer

The findings and conclusions in this publication are those of the authors and do not necessarily represent the views of the US Centers for Disease Control and Prevention, the US Department of Health and Human Services, or the National Institutes of Health.

### Contributors

MER, JG, and FOtK conceived the idea for the study. MER, JG, and FOtK wrote the protocol. JG, MMa, AK, HCB, SK, JL, FM, RK, MRK, DM, JC, MKL, EM, AARK, EA, OM, RNO, ADG, JDO, JH, MD, PJ, GD, and FOtK collected the original data and provided individual participant data. MER, JG, and FOtK contributed to data acquisition. MER and JG conducted the search and identifed studies based on the selection criteria; FOtK served as the tiebreaker. MER conducted the bias assessment, with support from FOtK. MER abstracted all the data in collaboration with the investigators of the original trials. MER performed the statistical analysis with inputs from JG, FOtK, and MMu. MER, JG, and FOtK wrote the first draft of the manuscript. All authors interpreted the data and critically reviewed the manuscript.

### Data sharing

Individual participant data from the source trials are available from the investigators from the source trials and will be uploaded onto the Worldwide Antimalarial Resistance Network (WWARN) repository approximately three months after publication.

## Acknowledgements

This study received financial support from The Bill and Melinda Gates Foundation (Award Number OPP1181807). MER is supported by the Eunice Kennedy Shriver National Institute Of Child Health & Human Development of the National Institutes of Health (Award Number K99HD111572). We are grateful to the study participants and dedicated study staff and researchers for their contributions to the study. We thank Carole Khairallah and James Dodd for supporting data acquisition from the original trials.

