## Appendix for "Dihydroartemisinin-piperaquine versus sulfadoxine-pyrimethamine for intermittent preventive treatment of malaria in pregnancy: a systematic review and individual participant data meta-analysis"

Figure S-23. Mean length-for-age z-score at two months of life ... 47
Figure S-24. Mean weight-for-age z-score at two months of life ... 48
Figure S-25. Mean weight-for-length z-score at two months of life ... 49

**Appendix 8. Study-specific effect estimates from mediation analyses ... 50**

Figure S-26. IPTp differences in BWGA z-scores mediated by incidence of clinical malaria during pregnancy ... 50
Figure S-27. IPTp differences in BWGA z-scores mediated by placental malaria infection defined as any evidence of pigment or parasites detected by histopathology, microscopy, PCR, or RDT ... 51
Figure S-28. IPTp differences in BWGA z-scores mediated by gestational weight gain ... 52
Figure S-29. IPTp differences in BWGA z-scores mediated by MUAC ... 53

**Appendix 9. Results for excluded Okoro et al, Nigeria (2023) trial ... 54**

Table S-9. Description of study characteristics and outcomes collected ... 55
Figure S-30. Birth outcomes ... 57
Figure S-31. Malaria Outcomes ... 58

**Appendix 10. Appendix References ... 59**

### Appendix 1. PRISMA 2020 Checklist

| Section and Topic | Item # | Checklist item | Location where item is reported |
|---|---|---|---|
| **TITLE** | | | |
| Title | 1 | Identify the report as a systematic review. | Paper title |
| **ABSTRACT** | | | |
| Abstract | 2 | See the PRISMA 2020 for Abstracts checklist. | N/A |
| **INTRODUCTION** | | | |
| Rationale | 3 | Describe the rationale for the review in the context of existing knowledge. | Paragraphs 2-3 |
| Objectives | 4 | Provide an explicit statement of the objective(s) or question(s) the review addresses. | Paragraph 4 |
| **METHODS** | | | |
| Eligibility criteria | 5 | Specify the inclusion and exclusion criteria for the review and how studies were grouped for the syntheses. | "Search strategy and selection criteria" section |
| Information sources | 6 | Specify all databases, registers, websites, organisations, reference lists and other sources searched or consulted to identify studies. Specify the date when each source was last searched or consulted. | |
| Search strategy | 7 | Present the full search strategies for all databases, registers and websites, including any filters and limits used. | |
| Selection process | 8 | Specify the methods used to decide whether a study met the inclusion criteria of the review, including how many reviewers screened each record and each report retrieved, whether they worked independently, and if applicable, details of automation tools used in the process. | "Data extraction and quality assessment" section |
| Data collection process | 9 | Specify the methods used to collect data from reports, including how many reviewers collected data from each report, whether they worked independently, any processes for obtaining or confirming data from study investigators, and if applicable, details of automation tools used in the process. | |
| Data items | 10a | List and define all outcomes for which data were sought. Specify whether all results that were compatible with each outcome domain in each study were sought (e.g. for all measures, time points, analyses), and if not, the methods used to decide which results to collect. | "Study endpoints" section; Appendices 2-4 |
| | 10b | List and define all other variables for which data were sought (e.g. participant and intervention characteristics, funding sources). Describe any assumptions made about any missing or unclear information. | Appendices 2-4 |
| Study risk of bias assessment | 11 | Specify the methods used to assess risk of bias in the included studies, including details of the tool(s) used, how many reviewers assessed each study and whether they worked independently, and if applicable, details of automation tools used in the process. | "Data extraction and quality assessment" section; Appendix 5 |

| Section and Topic | Item # | Checklist item | Location where item is reported |
|---|---|---|---|
| Effect measures | 12 | Specify for each outcome the effect measure(s) (e.g. risk ratio, mean difference) used in the synthesis or presentation of results. | “Statistical analysis” section |
| Synthesis methods | 13a | Describe the processes used to decide which studies were eligible for each synthesis (e.g. tabulating the study intervention characteristics and comparing against the planned groups for each synthesis (item #5)). | N/A |
| | 13b | Describe any methods required to prepare the data for presentation or synthesis, such as handling of missing summary statistics, or data conversions. | N/A |
| | 13c | Describe any methods used to tabulate or visually display results of individual studies and syntheses. | “Statistical analysis” section |
| | 13d | Describe any methods used to synthesize results and provide a rationale for the choice(s). If meta-analysis was performed, describe the model(s), method(s) to identify the presence and extent of statistical heterogeneity, and software package(s) used. | “Statistical analysis” section |
| | 13e | Describe any methods used to explore possible causes of heterogeneity among study results (e.g. subgroup analysis, meta-regression). | “Statistical analysis” section |
| | 13f | Describe any sensitivity analyses conducted to assess robustness of the synthesized results. | N/A |
| Reporting bias assessment | 14 | Describe any methods used to assess risk of bias due to missing results in a synthesis (arising from reporting biases). | N/A |
| Certainty assessment | 15 | Describe any methods used to assess certainty (or confidence) in the body of evidence for an outcome. | “Statistical analysis” section; Appendix 4 |
| **RESULTS** | | | |
| Study selection | 16a | Describe the results of the search and selection process, from the number of records identified in the search to the number of studies included in the review, ideally using a flow diagram. | “Description of studies” section; Figure 1 |
| | 16b | Cite studies that might appear to meet the inclusion criteria, but which were excluded, and explain why they were excluded. | “Description of studies” section |
| Study characteristics | 17 | Cite each included study and present its characteristics. | “Description of studies” section; Appendix 2 |
| Risk of bias in studies | 18 | Present assessments of risk of bias for each included study. | “Description of studies” section |
| Results of | 19 | For all outcomes, present, for each study: (a) summary statistics for each group (where appropriate) and (b) | Figures 2-6; Results |

| Section and Topic | Item # | Checklist item | Location where item is reported |
|---|---|---|---|
| individual studies | | an effect estimate and its precision (e.g. confidence/credible interval), ideally using structured tables or plots. | section |
| Results of syntheses | 20a | For each synthesis, briefly summarise the characteristics and risk of bias among contributing studies. | "Description of studies" section |
| | 20b | Present results of all statistical syntheses conducted. If meta-analysis was done, present for each the summary estimate and its precision (e.g. confidence/credible interval) and measures of statistical heterogeneity. If comparing groups, describe the direction of the effect. | All pooled estimates are presented with 95% CIs and $I^2$ statistic for in Results section + Figures 2-6 |
| | 20c | Present results of all investigations of possible causes of heterogeneity among study results. | N/A |
| | 20d | Present results of all sensitivity analyses conducted to assess the robustness of the synthesized results. | N/A |
| Reporting biases | 21 | Present assessments of risk of bias due to missing results (arising from reporting biases) for each synthesis assessed. | N/A |
| Certainty of evidence | 22 | Present assessments of certainty (or confidence) in the body of evidence for each outcome assessed. | 95% CIs are presented with all outcomes |
| **DISCUSSION** | | | |
| Discussion | 23a | Provide a general interpretation of the results in the context of other evidence. | Paragraph 1-3 |
| | 23b | Discuss any limitations of the evidence included in the review. | Paragraph 4 |
| | 23c | Discuss any limitations of the review processes used. | Paragraph 4 |
| | 23d | Discuss implications of the results for practice, policy, and future research. | Paragraph 5 |
| **OTHER INFORMATION** | | | |
| Registration and protocol | 24a | Provide registration information for the review, including register name and registration number, or state that the review was not registered. | "Data extraction and quality assessment" section of Methods |
| | 24b | Indicate where the review protocol can be accessed, or state that a protocol was not prepared. | |
| | 24c | Describe and explain any amendments to information provided at registration or in the protocol. | N/A |
| Support | 25 | Describe sources of financial or non-financial support for the review, and the role of the funders or sponsors in the review. | Summary, "Role of funding source" section of Methods, "Acknowledgement" |

| Section and Topic | Item # | Checklist item | Location where item is reported |
|---|---|---|---|
| | | | section |
| Competing interests | 26 | Declare any competing interests of review authors. | "Declaration of interests" section |
| Availability of data, code and other materials | 27 | Report which of the following are publicly available and where they can be found: template data collection forms; data extracted from included studies; data used for all analyses; analytic code; any other materials used in the review. | "Data sharing statement" section |

*From:* Page MJ, McKenzie JE, Bossuyt PM, Boutron I, Hoffmann TC, Mulrow CD, et al. The PRISMA 2020 statement: an updated guideline for reporting systematic reviews. BMJ 2021;372:n71. doi: 10.1136/bmj.n71

### Appendix 2. Description of study characteristics and outcomes for each study

| Study Information | Desai 2015[1] | Kakuru 2016[2] | Kajubi 2019[3] | Mlugu 2021[4] | Madanitsa 2023[5] studies | Gutman unpublished[6] |
|---|---|---|---|---|---|---|
| **Study Details** | | | | | | |
| Source | PMID: 26429700 | PMID: 26962728 | PMID: 30910321 | PMID: 33891721 | PMID: 36913959 | ClinicalTrials.Gov: NCT03009526 |
| Study site(s) | 4 health facilities in Siaya County, Kenya | Tororo District Hospital in Tororo District, E. Uganda | Masafu General Hospital in Busia District, E. Uganda | Kibiti District Center in Kibiti District, SE. Tanzania | 12 ANC clinics in W. Kenya (n=4) S. Malawi (n=5), NE Tanzania (n=3) | Machinga District Hospital in Liwonde, S. Malawi |
| Prevalence of PfDHPS 540E mutation, %* | 96% | 85% | 98% | 90% | Kenya: 65%; Malawi: 90%; Tanzania: 52% | 99% |
| Prevalence of PfDHPS 581G mutation, %* | 6% | 0% | 3% | 1% | Kenya: 11%; Malawi: 8%; Tanzania: 40% | 8% |
| Number of Participants Randomized (Among Singleton Pregnancies) | 1012 | 194 | 769 | 956 | Kenya: 992; Malawi: 938; Tanzania: 1192 | 593 |
| Sulfadoxine-Pyrimethamine | 508 | 104 | 381 | 478 | Kenya: 495; Malawi: 469; Tanzania: 597 | 297 |
| Dihydroartemisinin-Piperaquine | 504 | 90 | 388 | 478 | Kenya: 497; Malawi: 469; Tanzania: 595 | 296 |
| IPTp dosing regimen | every ANC | every 8 weeks | every 4 weeks | every 4 weeks | every 4 weeks | every 4 weeks |
| Number of IPTp doses, median (IQR) | 2 (2-3) | 3 (3-3) | 6 (5-6) | 3 (2-4) | Kenya: 4 (4-5); Malawi: 4 (4-5); Tanzania: 5 (4-6) | 4 (3-5) |

| Study Information | Desai 2015[1] | Kakuru 2016[2] | Kajubi 2019[3] | Mlugu 2021[4] | Madanitsa 2023[5] studies | Gutman unpublished[6] |
|---|---|---|---|---|---|---|
| PCR positivity at enrolment, % | 32% | 58% | 81% | 14% | Kenya: 15%; Malawi: 11%; Tanzania: 18% | 11% |
| **Birth outcomes** | | | | | | |
| Foetal Loss | Available | Available | Available | Available | Available | Available |
| Small-for-Gestational Age | Available† | Available | Available | Available† | Available | Available |
| Preterm Delivery | Available† | Available | Available | Available† | Available | Available |
| Low Birthweight | Available | Available | Available | Available | Available | Available |
| Neonatal Death | Available | Available | Available | Available (no cases) | Available | Available |
| **Continuous Birth Outcomes** | | | | | | |
| Mean Birthweight | Available | Available | Available | Available | Available | Available |
| Mean Gestational Age at Delivery | Available† | Available | Available | Available† | Available | Available |
| Mean Birthweight-for-Gestational Age Z-scores | Available† | Available | Available | Available† | Available | Available |
| **Malaria Outcomes** | | | | | | |
| Incidence of Clinical Malaria Episodes in Pregnancy | Available | Available | Available | Available | Available | Available |
| Any Evidence of Pigment Only in Placental Tissue by Histopathology | Available | Available | Available | Available | Available | Available |

| Study Information | Desai 2015[1] | Kakuru 2016[2] | Kajubi 2019[3] | Mlugu 2021[4] | Madanitsa 2023[5] studies | Gutman unpublished[6] |
|---|---|---|---|---|---|---|
| Any Evidence of Parasites in Placental Tissue or Blood by Histopathology, PCR, Microscopy, or RDT | Available | Available; RDT not done on placental blood | Available; RDT not done on placental blood | Available; RDT not done on placental blood | Available | Available |
| Any Evidence of Parasites or Pigment in Placental Tissue or Blood by Histopathology, PCR, Microscopy, or RDT | Available | Available; RDT not done on placental blood | Available; RDT not done on placental blood | Available; RDT not done on placental blood | Available | Available |
| Any Evidence of Parasites in Maternal Peripheral Blood at Delivery by RDT, Microscopy, or PCR | Available | Available; RDT not done on maternal blood | Available; RDT and PCR not done on maternal blood | Available | Available | Available; PCR not done on maternal blood |
| **Maternal Outcomes** | | | | | | |
| Any Evidence of Severe Anaemia (Hb <7 g/dl) During Pregnancy | Available; Assessed at routine visits | Available; Assessed at routine visits | Available; Assessed at routine visits | Available; Assessed at routine visits | Available; Assessed at routine visits | Not Available; Measured at Delivery Only |
| Any Evidence of Moderate Anaemia (Hb <9 g/dl) During Pregnancy | Available; Assessed at routine visits | Available; Assessed at routine visits | Available; Assessed at routine visits | Available; Assessed at routine visits | Available; Assessed at routine visits | Not Available; Measured at Delivery Only |
| Any Evidence of Mild Anaemia (Hb <11 g/dl) During Pregnancy | Available; Assessed at routine visits | Available; Assessed at routine visits | Available; Assessed at routine visits | Available; Assessed at routine visits | Available; Assessed at routine visits | Not Available; Measured at Delivery Only |
| MUAC at Delivery | Available; but unadjusted estimates b/c enrolment data was unavailable | Not available | Not available | Not available | Available | Available |
| Maternal weight gain per week[‡] | Available | Available | Available | Available | Available | Available |

| Study Information | Desai 2015[1] | Kakuru 2016[2] | Kajubi 2019[3] | Mlugu 2021[4] | Madanitsa 2023[5] studies | Gutman unpublished[6] |
|---|---|---|---|---|---|---|
| **Infant Outcomes** | | | | | | |
| Any Evidence of Stunting (LAZ <2 SD) from Birth to 2 Months of Life | Available; Follow up data up to 6-8 weeks | Available; Follow-up data up to 8 weeks | Available; Follow-up data up to 8 weeks | Not available | Available; Follow up data up to 6-8 weeks | Available; Follow up data up to10 weeks |
| Any Evidence of Underweight (WAZ <2 SD) from Birth to 2 Months of Life | Available; Follow up data up to 6-8 weeks | Available; Follow-up data up to 8 weeks | Available; Follow-up data up to 8 weeks | Not available | Available; Follow up data up to 6-8 weeks | Available; Follow up data up to10 weeks |
| Any Evidence of Wasting (WLZ <2 SD) from Birth to 2 Months of Life | Available; Follow up data up to 6-8 weeks | Available; Follow-up data up to 8 weeks | Available; Follow-up data up to 8 weeks | Not available | Available; Follow up data up to 6-8 weeks | Available; Follow up data up to10 weeks |
| Mean LAZ at 2 Months of Life | Available; Follow up data up to 6-8 weeks | Available; Follow-up data up to 8 weeks | Available; Follow-up data up to 8 weeks | Not available | Available; Follow up data up to 6-8 weeks | Available; Follow up data up to10 weeks |
| Mean WAZ at 2 Months of Life | Available; Follow up data up to 6-8 weeks | Available; Follow-up data up to 8 weeks | Available; Follow-up data up to 8 weeks | Not available | Available; Follow up data up to 6-8 weeks | Available; Follow up data up to10 weeks |
| Mean WLZ at 2 Months of Life | Available; Follow up data up to 6-8 weeks | Available; Follow-up data up to 8 weeks | Available; Follow-up data up to 8 weeks | Not available | Available; Follow up data up to 6-8 weeks | Available; Follow up data up to10 weeks |

Abbreviations: ANC = antenatal care visit; Hb = haemoglobin; IQR = interquartile range; LAZ = length-for-age z-score; MUAC = mid-upper arm circumference; WAZ = weight-for-age z-score; WLZ = weight-for-length z-score

* Prevalence of polymorphisms were reported as part of the published findings of the trial, except for the Kakuru et al (2016) and Kajubi et al (2019) studies which were reported separately in Conrad et al (2017)[7] NS Nayebare et al (2020)[8], respectively. Data were not available from Gutman et al (unpublished) and we therefore used estimates from a separate study (Gutman et al (2015)[9]) which was conducted at the same site eight years earlier.

† Gestational age dating not confirmed by ultrasound

‡ Maternal weight gain per week calculated using the following formula: $\frac{Weight_{last\ ANC\ visit\ before\ delivery} - Weight_{enrollment}}{\#\ of\ weeks\ between\ enrollment\ and\ last\ ANC\ visit}$

### Appendix 3. Definition of outcomes

| Outcome | Description |
|---|---|
| **Birth outcomes** | |
| Any Adverse Pregnancy Outcome | Binary variable defined as a composite outcome of any one of the following conditions: miscarriage (stillbirth (foetal loss ≥28 gestational weeks), preterm birth (PTB; delivery <37 gestational weeks), small-for-gestational age (SGA; birthweight <10$^{th}$ percentile for gestational age using the INTERGROWTH-21$^{st}$ standard[10]); low birthweight (LBW; birthweight <2,500 grams), and neonatal loss (newborn death within the first 28 days of life). |
| Foetal Loss | Binary variable defined as a composite outcome of miscarriage or abortion that occurred at less than 28 gestational weeks |
| Small-for-Gestational Age | Binary variable defined as birthweight below the 10$^{th}$ percentile for a given gestational age and sex using the INTERGROWTH-21$^{st}$ standard[10] |
| Preterm Delivery | Binary variable defined as delivery occurring less than 37 gestational weeks among live births |
| Low Birthweight | Binary variable defined as newborn birthweight that was less than 2500 grams. Birthweights were assessed among live births. Corrected birthweights were used when available. |
| Neonatal Death | Defined as a binary variable indicating the death of live newborn within the first 28 days of life |
| **Continuous Birth Outcomes** | |
| Birthweight | Continuous measure of newborn birthweight in grams. Birthweights were assessed among live births. Corrected birthweights were used when available. |
| Gestational Age at Birth | Continuous measure of duration of gestation in weeks. Gestational age at birth was estimated using the gestational age at enrolment assessed by ultrasound, except for Desai 2015 and Mlugu 2021. Desai 2015 used the Ballard score, while Mlugu 2021 used last menstrual period to estimate gestational age at delivery. |
| Birthweight-for-Gestational Age Z-scores | Continuous measure of birthweight-for-gestational age z-scores. Z-scores were calculated based on INTERGROWTH-21$^{st}$ standards[10] |
| **Malaria outcomes** | |
| Incidence of Clinical Malaria Episodes in Pregnancy | Count outcome defined as the number of symptomatic malaria episodes experienced during the pregnancy follow-up period. Clinical malaria was defined as presence of documented fever (≥37·5°C or defined by study-specific definitions) or recent history of fever in the last 48 hours (or as defined by study-specific definitions) and a positive diagnosis of malaria by either rapid diagnostic test or blood smear microscopy. |
| Any Evidence of Pigment Only in Placental Tissue by Histopathology | Binary variable defined as the presence of pigment (but not parasites) in the placental tissue assessed by histopathology. For this outcome, active and chronic placental infections as defined by Rogerson et al[11] were excluded from the analysis. |

| | |
|---|---|
| Any Evidence of Parasites in Placental Tissue or Blood by Histopathology, PCR, Microscopy, or RDT | Binary variable defined as the presence of parasites in the placental tissue or blood assessed by histopathology, PCR, microscopy, or RDT. For this outcome, samples containing pigment with no indication of parasites in the placental tissue or blood were considered in the "No" category. |
| Any Evidence of Parasites or Pigment in Placental Tissue or Blood by Histopathology, PCR, Microscopy, or RDT | Binary variable defined as the presence of parasites or pigment in the placental tissue or blood assessed by histopathology, PCR, microscopy, or RDT. For this outcome, samples containing pigment with no indication of parasites in the placental tissue or blood were considered in the "Yes" category. |
| Any Evidence of Parasites in Maternal Peripheral Blood at Delivery by RDT, Microscopy, or PCR | Binary variable defined as the presence of parasites detected in maternal peripheral blood at delivery by PCR, microscopy, or RDT. |
| **Maternal outcomes** | |
| Any Evidence of Severe Anaemia (Hb <7 g/dl) During Pregnancy | Binary variable defined as the incidence of a severe anaemic episode detected during routine and/or unscheduled visits during pregnancy follow-up period. Severe anaemia was defined as a haemoglobin measurement that fell below 7 g/dL. |
| Any Evidence of Moderate Anaemia (Hb <9 g/dl) During Pregnancy | Binary variable defined as the incidence of a moderate anaemic episode detected during routine and/or unscheduled visits during pregnancy follow-up period. Moderate anaemia was defined as a haemoglobin measurement that fell below 9 g/dL. |
| Any Evidence of Mild Anaemia (Hb <11 g/dl) During Pregnancy | Binary variable defined as the incidence of a moderate anaemic episode detected during routine and/or unscheduled visits during pregnancy follow-up period. Moderate anaemia was defined as a haemoglobin measurement that fell below 11 g/dL. |
| MUAC at Delivery | Continuous measure of maternal mid-upper arm circumference measured at delivery. |
| Maternal weight gain per week[‡] | Continuous variable defined as the average weight gained per week from enrolment to last day of antenatal care visit before delivery. The following formula was used to define this outcome: $\frac{Weight_{last\ ANC\ visit\ before\ delivery} - Weight_{enrollment}}{\#\ of\ weeks\ between\ enrollment\ and\ last\ ANC\ visit}$ |
| **Infant Outcomes** | |
| Any Evidence of Stunting (LAZ <2 SD) from Birth to 2 Months of Life | Binary variable defined as a length-for-age z-score that fell below two standard deviations below the median 2006 WHO Child Growth Standards[12] at birth and at routine scheduled neonatal care visits up to approximately two months of life. Infant follow-up periods varied between studies: six-eight weeks in the Desai 2015 and Madanitsa 2023 studies, eight weeks in the Kakuru 2016 and Kajubi 2019 studies, and ten weeks in the Gutman unpublished study. |
| Any Evidence of Underweight (WAZ <2 SD) from Birth to 2 Months of Life | Binary variable defined as a weight-for-age z-score that fell below two standard deviations below the median 2006 WHO Child Growth Standards[12] at birth and at routine scheduled neonatal care visits up to approximately two months of life. Infant follow-up |

| | |
|---|---|
| | periods varied between studies: six-eight weeks in the Desai 2015 and Madanitsa 2023 studies, eight weeks in the Kakuru 2016 and Kajubi 2019 studies, and ten weeks in the Gutman unpublished study. |
| Any Evidence of Wasting (WLZ <2 SD) from Birth to 2 Months of Life | Binary variable defined as a weight-for-length z-score that fell below two standard deviations below the median 2006 WHO Child Growth Standards[12] at birth and at routine scheduled neonatal care visits up to approximately two months of life. Infant follow-up periods varied between studies: six-eight weeks in the Desai 2015 and Madanitsa 2023 studies, eight weeks in the Kakuru 2016 and Kajubi 2019 studies, and ten weeks in the Gutman unpublished study. |
| LAZ at 2 Months of Life | Continuous variable defined as a length-for-age z-score based on 2006 WHO Child Growth Standards.[12] |
| WAZ at 2 Months of Life | Continuous variable defined as a weight-for-age z-score based on 2006 WHO Child Growth Standards.[12] |
| WLZ at 2 Months of Life | Continuous variable defined as a weight-for-length z-score based on 2006 WHO Child Growth Standards.[12] |

### Appendix 4. Details of causal mediation analyses methods

Causal mediation analyses were performed to quantify the contributions of placental malaria, gestational weight gain (GWG), and maternal mid-upper arm circumference (MUAC) on the differential impact of IPTp regimens on birthweight-for-gestational age z-scores.

**Brief summary of causal mediation analysis.** Mediation analyses were conducted following the "potential outcomes" framework as described by Rubin[13] and Pearl[14,15] and extended for mediation analyses by Imai.[16] Under this framework, potential outcomes are defined for each individual based on their counterfactual treatment condition. Let A, Y, and W denote the treatment, outcome, and confounder for each individual *i*. In the scenario where treatment is binary, an individual would have two potential outcomes: one had they been treated ($Y_i(a=1)$) and another if they had not been treated ($Y_i(a=0)$). The causal effect of the treatment on the outcome for individual *i* is defined as $Y_i(a=1)$ - $Y_i(a=0)$ and E[$Y_i(a=1)$ - $Y_i(a=0)$] is defined as the average causal effect.

For mediation analyses, this framework is extended to further decompose the above causal effect (also known as the "total effect") into effects that are mediated and not mediated through some intermediate variable of interest, M. This requires the specification four potential outcomes for each individual:

$$Y_i(a=0, M_i(a=0)) \qquad Y_i(a=0, M_i(a=1))$$
$$Y_i(a=1, M_i(a=0)) \qquad Y_i(a=1, M_i(a=1))$$

where $Y_i(a, M_i(a))$ represents the outcome value for individual *i* had treatment been set to some value *a* and the mediator value was set to the value it would have taken on under *a*. For example, $Y_i(a=1, M_i(a=1))$ represents the outcome value for individual *i* had they been treated and their mediator value takes on what it would have "naturally" taken on had the individual been treated. By estimating potential mediator values, $M_i(a=0)$ and $M_i(a=1)$, we can separately estimate the **natural indirect effect (NIE)**, where NIE = $Y_i(a, M_i(a=1))$ - $Y_i(a, M_i(a=0))$ and is defined as the effect of the treatment on the outcome that is mediated through M (i.e., mediated effect). The **natural direct effect (NDE)** is defined as the effect of the treatment on the outcome via pathways not through mediator M (i.e., non-mediated effect), where NDE = $Y_i(a=1, M_i(a))$ - $Y_i(a=0, M_i(a))$.

Of these four potential outcomes, two can never be observed under real world conditions (i.e., $Y_i(a=1, M_i(a=0))$ and $Y_i(a=0, M_i(a=1))$) and depending the individual's actual treatment value, either one of the two remaining potential outcome values are actually observed ($Y_i(a=1, M_i(a=1))$ or $Y_i(a=0, M_i(a=0))$). However, these values are needed to derive NIE AND NDE; therefore, unobserved values must be estimated from data.

**Required assumptions.** To interpret NIE and NDE estimates causally, several assumptions need to be met[17], including:

- No unmeasured confounding between treatment-outcome, treatment-mediator, and mediator-outcome
- Treatment and mediator positivity
- Well-defined treatment and mediator
- Mediator and outcome values are unaffected by another individual's treatment and mediator condition
- No presence of a mediator-outcome confounder that is itself affected by the treatment

**Targeted Maximum Likelihood Estimation Approach to Causal Mediation Analysis.** We used the semi-parametric, doubly robust targeted maximum likelihood estimation (TMLE) approach to estimate NIE and

NDE. TMLE offers advantages over traditional parametric approaches by imposing fewer assumptions about the underlying data generating process. It does so by allowing data-adaptive, ensemble machine-learning approaches (e.g., SuperLearner via the sl3 package[18]) to flexibility accommodate interactions and non-linear relationships between variables, thereby reducing the risk of model misspecification.

A detailed description of the TMLE approach is provided in Zheng and van Der Laan (2012).[19] In brief, the observed data is used to generate an initial outcome model estimating the expected outcome conditional on treatment, mediator, and confounders (i.e., $Q[Y|A, M, W]$). The initial outcome estimates are then updated using "clever covariates" derived from two propensity score models: one estimating the conditional probability given confounders ($g(A|W)$) and one estimating the conditional probability of treatment given mediator and confounders ($g(A|M, W)$). This process, known as the "targeting step" of TMLE, is performed to ensure that the final estimates optimize the bias-variance trade-off, thereby enhancing the precision, efficiency, and robustness of causal effect estimates. Finally, this updated estimate is used to generate updated estimates of the four potential outcomes and NIE and NDE are calculated as the mean difference across these potential outcomes (using prior formulas).

**Mediation analysis methods.** The directed acyclic graph (DAG) on the right was used to represent our assumptions of the causal relationships between treatment mediator, outcome, and confounders. Mediators were analyses separately to prevent overly complex models given our small sample size and to reduce the risk of violating the treatment and mediator positivity assumption.

Directed Acyclic Graph

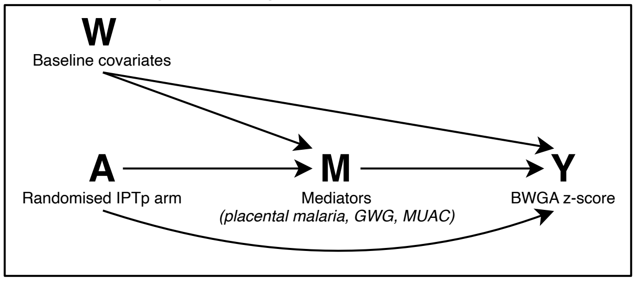

The `medoutcon` package in R[20] was used to estimate NIE, NDE, and the corresponding 95% confidence intervals. Definitions of the treatment, mediator, outcome, and confounders used in the analyses are provided in the table below. The algorithm library for the initial outcome model included elastic net regularization, lasso (L-1 penalized) regression, generalized linear regression, generalized additive models, and extreme gradient boosting. The algorithm library for propensity score models included simple intercept models, generalized additive models, and gradient boosted decision tree models. 10-fold cross-validation was used all applications. Observations with missing values were excluded from the analysis.

**Variable definitions**

| Variable | Description |
|---|---|
| Treatment (A) | Randomized assignment to IPTp with sulfadoxine-pyrimethamine (A=1) or dihydroartemisinin-piperaquine (A=0) |
| Mediators (M) | • **Placental malaria**, defined as any evidence of pigment or parasites in the placental tissue or blood detected by histopathology, RDT, microscopy, and/or PCR<br>• **Gestational weight gain**, defined as mean weight gain per week from enrolment to last antenatal care visit before delivery<br>• **Maternal mid-upper arm circumference** in cm measured at delivery |
| Outcome (Y) | Birthweight-for-gestational age z-score using INTERGROWTH-21$^{st}$ birthweight standards |
| Confounders (W) *measured at enrolment* | Maternal age, BMI, weight, and gestational age; highest schooling level completed by mother (none, primary, secondary, or higher); and infant sex |

### Appendix 5. Bias assessment using the Cochrane Risk of Bias Tool 2.0

**Summary.** The Cochrane risk of bias tool for randomized trials (RoB 2.0) was applied to each study. As the primary outcome of the individual trials often differed from the primary outcome of our meta-analysis and varied between studies, we conducted the risk of bias assessment based on the primary outcome of the meta-analysis. Overall, six of the eight trials were considered to have a low risk of bias; two studies (Desai et al, 2015 and Mlugu et al, 2021) had some concerns regarding the primary outcome because gestational age dating was not confirmed by ultrasound. This may have resulted in misclassification of small-for-gestational age and preterm birth, both of which are included as components in the composite primary outcome. Detailed visualizations from the RoB 2.0 assessment are provided below. Plots were generated using the `robvis` package in R.[21]

##### Risk of bias traffic light plot

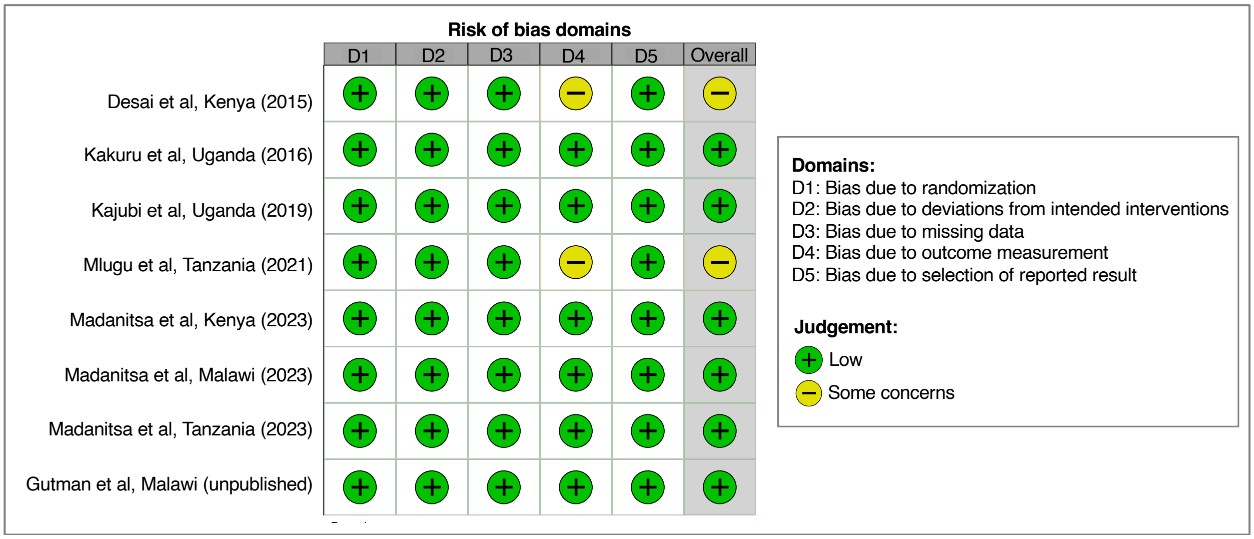

##### Risk of bias summary plot

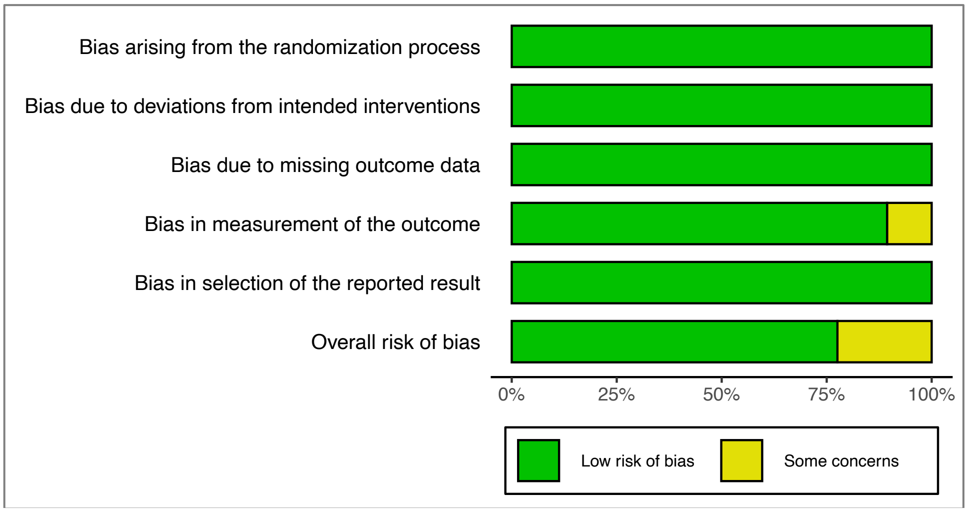

### Appendix 6. Participant characteristics at enrolment by arm

Table S-1. Desai et al, Kenya (2015)

| Enrolment characteristics | Total N=1,012 | IPTp-SP N=508 | IPTp-DP N=504 |
|---|---|---|---|
| **Maternal age in years, mean (SD)** | 23·4 (5·7) | 23·5 (5·9) | 23·4 (5·5) |
| **Gestational age in weeks, mean (SD)** | 22·9 (4·8) | 22·8 (4·9) | 23·0 (4·8) |
| **Gravidity categories, n/N (%)** | | | |
| Primigravidae | 336/1012 (33%) | 178/508 (35%) | 158/504 (31%) |
| Secundigravidae | 215/1012 (21%) | 112/508 (22%) | 103/504 (20%) |
| Multigravidae (3+) | 461/1012 (46%) | 218/508 (43%) | 243/504 (48%) |
| **Weight in kg, mean (SD)** | 61·6 (9·2) | 61·5 (9·1) | 61·7 (9·4) |
| **Height in cm, mean (SD)** | 164·3 (6·8) | 164·3 (6·9) | 164·3 (6·7) |
| **Maternal MUAC in cm, mean (SD)** | -- | -- | -- |
| **Highest level of schooling completed, n/N (%)** | | | |
| None | 229/1004 (23%) | 114/505 (23%) | 115/499 (23%) |
| Primary | 569/1004 (57%) | 283/505 (56%) | 286/499 (57%) |
| Secondary | 183/1004 (18%) | 96/505 (19%) | 87/499 (17%) |
| Higher | 23/1004 (2%) | 12/505 (2%) | 11/499 (2%) |
| **Wealth index tertiles, n/N (%)** | | | |
| Lowest tertile | 328/1006 (33%) | 172/505 (34%) | 156/501 (31%) |
| Middle tertile | 331/1006 (33%) | 164/505 (32%) | 167/501 (33%) |
| Highest tertile | 347/1006 (34%) | 169/505 (33%) | 178/501 (36%) |
| **Slept under a bed net last night, n/N (%)** | 725/1012 (72%) | 364/508 (72%) | 361/504 (72%) |
| **Microscopy positivity, n/N (%)** | 154/984 (16%) | 79/499 (16%) | 75/485 (15%) |
| **PCR/LAMP positivity, n/N (%)** | 322/1002 (32%) | 167/504 (33%) | 155/498 (31%) |

Abbreviations: LAMP = loop-mediated isothermal amplification method MUAC = mid-upper arm circumference; PCR = polymerase chain reaction

#### Table S-2. Kakuru et al, Uganda (2016)

| Enrolment characteristics | Total<br>N=194 | IPTp-SP<br>N=104 | IPTp-DP<br>N=90 |
|---|---|---|---|
| **Maternal age in years, mean (SD)** | 21·7 (4·0) | 21·3 (3·6) | 22·2 (4·3) |
| **Gestational age in weeks, mean (SD)** | 15·3 (2·0) | 15·2 (2·0) | 15·4 (2·0) |
| **Gravidity categories, n/N (%)** | | | |
| Primigravidae | 73/194 (38%) | 42/104 (40%) | 31/90 (34%) |
| Secundigravidae | 58/194 (30%) | 30/104 (29%) | 28/90 (31%) |
| Multigravidae (3+) | 63/194 (32%) | 32/104 (31%) | 31/90 (34%) |
| **Weight in kg, mean (SD)** | 55·6 (6·9) | 55·5 (6·8) | 55·7 (7·1) |
| **Height in cm, mean (SD)** | 162·7 (6·8) | 162·8 (6·8) | 162·5 (6·9) |
| **Maternal MUAC in cm, mean (SD)** | -- | -- | -- |
| **Highest level of schooling completed, n/N (%)** | | | |
| None | 9/194 (5%) | 6/104 (6%) | 3/90 (3%) |
| Primary | 143/194 (74%) | 74/104 (71%) | 69/90 (77%) |
| Secondary | 39/194 (20%) | 22/104 (21%) | 17/90 (19%) |
| Higher | 3/194 (2%) | 2/104 (2%) | 1/90 (1%) |
| **Wealth index tertiles, n/N (%)** | | | |
| Lowest tertile | 67/194 (35%) | 38/104 (37%) | 29/90 (32%) |
| Middle tertile | 66/194 (34%) | 31/104 (30%) | 35/90 (39%) |
| Highest tertile | 61/194 (31%) | 35/104 (34%) | 26/90 (29%) |
| **Slept under a bed net last night, n/N (%)** | 174/194 (90%) | 91/104 (88%) | 83/90 (92%) |
| **Microscopy positivity, n/N (%)** | -- | -- | -- |
| **PCR/LAMP positivity, n/N (%)** | 111/193 (58%) | 58/104 (56%) | 53/89 (60%) |

Abbreviations: LAMP = loop-mediated isothermal amplification method MUAC = mid-upper arm circumference; PCR = polymerase chain reaction

Table S-3. Kajubi et al, Uganda (2019)

| Enrolment characteristics | Total<br>N=769 | IPTp-SP<br>N=381 | IPTp-DP<br>N=388 |
|---|---|---|---|
| **Maternal age in years, mean (SD)** | 23·7 (5·8) | 23·8 (5·9) | 23·7 (5·6) |
| **Gestational age in weeks, mean (SD)** | 15·5 (2·4) | 15·5 (2·4) | 15·4 (2·3) |
| **Gravidity categories, n/N (%)** | | | |
| Primigravidae | 193/769 (25%) | 100/381 (26%) | 93/388 (24%) |
| Secundigravidae | 187/769 (24%) | 83/381 (22%) | 104/388 (27%) |
| Multigravidae (3+) | 389/769 (51%) | 198/381 (52%) | 191/388 (49%) |
| **Weight in kg, mean (SD)** | 55·9 (7·6) | 56·1 (7·7) | 55·6 (7·6) |
| **Height in cm, mean (SD)** | 158·6 (6·0) | 158·8 (6·2) | 158·4 (5·9) |
| **Maternal MUAC in cm, mean (SD)** | | | |
| **Highest level of schooling completed, n/N (%)** | | | |
| None | 63/769 (8%) | 37/381 (10%) | 26/388 (7%) |
| Primary | 512/769 (67%) | 248/381 (65%) | 264/388 (68%) |
| Secondary | 175/769 (23%) | 89/381 (23%) | 86/388 (22%) |
| Higher | 19/769 (2%) | 7/381 (2%) | 12/388 (3%) |
| **Wealth index tertiles, n/N (%)** | | | |
| Lowest tertile | 251/740 (34%) | 122/366 (33%) | 129/374 (34%) |
| Middle tertile | 248/740 (34%) | 116/366 (32%) | 132/374 (35%) |
| Highest tertile | 241/740 (33%) | 128/366 (35%) | 113/374 (30%) |
| **Slept under a bed net last night, n/N (%)** | 242/740 (33%) | 122/366 (33%) | 120/374 (32%) |
| **Microscopy positivity, n/N (%)** | 394/769 (51%) | 192/381 (50%) | 202/388 (52%) |
| **PCR/LAMP positivity, n/N (%)** | 622/769 (81%) | 313/381 (82%) | 309/388 (80%) |

Abbreviations: LAMP = loop-mediated isothermal amplification method MUAC = mid-upper arm circumference; PCR = polymerase chain reaction

Table S-4. Mlugu et al, Tanzania (2021)

| **Enrolment characteristics** | **Total**<br>**N=956** | **IPTp-SP**<br>**N=478** | **IPTp-DP**<br>**N=478** |
|---|---|---|---|
| **Maternal age in years, mean (SD)** | 26·6 (7·1) | 26·5 (7·1) | 26·6 (7·1) |
| **Gestational age in weeks, mean (SD)** | 21·5 (3·4) | 21·4 (3·5) | 21·7 (3·3) |
| **Gravidity categories, n/N (%)** | | | |
| Primigravidae | 243/956 (25%) | 128/478 (27%) | 115/478 (24%) |
| Secundigravidae | 213/956 (22%) | 105/478 (22%) | 108/478 (23%) |
| Multigravidae (3+) | 500/956 (52%) | 245/478 (51%) | 255/478 (53%) |
| **Weight in kg, mean (SD)** | 55·2 (8·9) | 54·8 (8·5) | 55·6 (9·3) |
| **Height in cm, mean (SD)** | 152·2 (3·1) | 152·1 (3·1) | 152·4 (3·0) |
| **Maternal MUAC in cm, mean (SD)** | -- | -- | -- |
| **Highest level of schooling completed, n/N (%)** | | | |
| None | 183/956 (19%) | 94/478 (20%) | 89/478 (19%) |
| Primary | 657/956 (69%) | 336/478 (70%) | 321/478 (67%) |
| Secondary | 116/956 (12%) | 48/478 (10%) | 68/478 (14%) |
| Higher | 0/956 (0%) | 0/478 (0%) | 0/478 (0%) |
| **Wealth index tertiles, n/N (%)** | | | |
| Lowest tertile | 251/740 (34%) | 122/366 (33%) | 129/374 (34%) |
| Middle tertile | 248/740 (34%) | 116/366 (32%) | 132/374 (35%) |
| Highest tertile | 241/740 (33%) | 128/366 (35%) | 113/374 (30%) |
| **Slept under a bed net last night, n/N (%)** | 695/956 (73%) | 345/478 (72%) | 350/478 (73%) |
| **Microscopy positivity, n/N (%)** | -- | -- | -- |
| **PCR/LAMP positivity, n/N (%)** | 136/956 (14%) | 63/478 (13%) | 73/478 (15%) |

Abbreviations: LAMP = loop-mediated isothermal amplification method MUAC = mid-upper arm circumference; PCR = polymerase chain reaction

Table S-5. Madanitsa et al, Kenya (2023)

| Enrolment characteristics | Total N=992 | IPTp-SP N=495 | IPTp-DP N=497 |
|---|---|---|---|
| **Maternal age in years, mean (SD)** | 23·8 (5·3) | 23·8 (5·4) | 23·8 (5·3) |
| **Gestational age in weeks, mean (SD)** | 21·5 (3·6) | 21·8 (3·7) | 21·3 (3·4) |
| **Gravidity categories, n/N (%)** | | | |
| Primigravidae | 355/992 (36%) | 178/495 (36%) | 177/497 (36%) |
| Secundigravidae | 268/992 (27%) | 131/495 (26%) | 137/497 (28%) |
| Multigravidae (3+) | 369/992 (37%) | 186/495 (38%) | 183/497 (37%) |
| **Weight in kg, mean (SD)** | 65·1 (11·2) | 66·1 (11·1) | 64·1 (11·2) |
| **Height in cm, mean (SD)** | 162·7 (6·8) | 163·4 (6·7) | 162·0 (6·9) |
| **Maternal MUAC in cm, mean (SD)** | 27·0 (3·2) | 27·2 (3·3) | 26·8 (3·2) |
| **Highest level of schooling completed, n/N (%)** | | | |
| None | 57/992 (6%) | 28/495 (6%) | 29/497 (6%) |
| Primary | 454/992 (46%) | 218/495 (44%) | 236/497 (47%) |
| Secondary | 334/992 (34%) | 170/495 (34%) | 164/497 (33%) |
| Higher | 147/992 (15%) | 79/495 (16%) | 68/497 (14%) |
| **Wealth index tertiles, n/N (%)** | | | |
| Lowest tertile | 46/992 (5%) | 17/495 (3%) | 29/497 (6%) |
| Middle tertile | 473/992 (48%) | 242/495 (49%) | 231/497 (46%) |
| Highest tertile | 473/992 (48%) | 236/495 (48%) | 237/497 (48%) |
| **Slept under a bed net last night, n/N (%)** | 895/900 (99%) | 440/445 (99%) | 455/455 (100%) |
| **Microscopy positivity, n/N (%)** | 127/981 (13%) | 61/490 (12%) | 66/491 (13%) |
| **PCR/LAMP positivity, n/N (%)** | 127/865 (15%) | 61/434 (14%) | 66/431 (15%) |

Abbreviations: LAMP = loop-mediated isothermal amplification method MUAC = mid-upper arm circumference; PCR = polymerase chain reaction

Table S-6. Madanitsa et al, Malawi (2023)

| Enrolment characteristics | Total N=938 | IPTp-SP N=469 | IPTp-DP N=469 |
|---|---|---|---|
| **Maternal age in years, mean (SD)** | 24·5 (5·9) | 24·3 (5·9) | 24·7 (5·9) |
| **Gestational age in weeks, mean (SD)** | 21·3 (3·1) | 21·2 (3·1) | 21·3 (3·1) |
| **Gravidity categories, n/N (%)** | | | |
| Primigravidae | 287/936 (31%) | 151/469 (32%) | 136/467 (29%) |
| Secundigravidae | 238/936 (25%) | 112/469 (24%) | 126/467 (27%) |
| Multigravidae (3+) | 411/936 (44%) | 206/469 (44%) | 205/467 (44%) |
| **Weight in kg, mean (SD)** | 58·7 (9·1) | 58·4 (8·8) | 59·0 (9·4) |
| **Height in cm, mean (SD)** | 157·3 (6·3) | 157·2 (6·1) | 157·5 (6·6) |
| **Maternal MUAC in cm, mean (SD)** | 26·3 (3·1) | 26·2 (3·1) | 26·4 (3·1) |
| **Highest level of schooling completed, n/N (%)** | | | |
| None | 90/938 (10%) | 42/469 (9%) | 48/469 (10%) |
| Primary | 586/938 (62%) | 306/469 (65%) | 280/469 (60%) |
| Secondary | 236/938 (25%) | 107/469 (23%) | 129/469 (28%) |
| Higher | 26/938 (3%) | 14/469 (3%) | 12/469 (3%) |
| **Wealth index tertiles, n/N (%)** | | | |
| Lowest tertile | 649/938 (69%) | 331/469 (71%) | 318/469 (68%) |
| Middle tertile | 165/938 (18%) | 78/469 (17%) | 87/469 (19%) |
| Highest tertile | 124/938 (13%) | 60/469 (13%) | 64/469 (14%) |
| **Slept under a bed net last night, n/N (%)** | 580/597 (97%) | 283/293 (97%) | 297/304 (98%) |
| **Microscopy positivity, n/N (%)** | 104/934 (11%) | 52/467 (11%) | 52/467 (11%) |
| **PCR/LAMP positivity, n/N (%)** | 159/867 (18%) | 79/433 (18%) | 80/434 (18%) |

Abbreviations: LAMP = loop-mediated isothermal amplification method MUAC = mid-upper arm circumference; PCR = polymerase chain reaction

Table S-7. Madanitsa et al, Tanzania (2023)

| Enrolment characteristics | Total<br>N=1,192 | IPTp-SP<br>N=597 | IPTp-DP<br>N=595 |
|---|---|---|---|
| **Maternal age in years, mean (SD)** | 26·5 (6·5) | 26·5 (6·6) | 26·5 (6·5) |
| **Gestational age in weeks, mean (SD)** | 20·0 (3·4) | 19·9 (3·3) | 20·1 (3·5) |
| **Gravidity categories, n/N (%)** | | | |
| Primigravidae | 324/1187 (27%) | 164/594 (28%) | 160/593 (27%) |
| Secundigravidae | 260/1187 (22%) | 130/594 (22%) | 130/593 (22%) |
| Multigravidae (3+) | 603/1187 (51%) | 300/594 (51%) | 303/593 (51%) |
| **Weight in kg, mean (SD)** | 59·5 (12·1) | 59·5 (12·4) | 59·5 (11·8) |
| **Height in cm, mean (SD)** | 155·9 (6·1) | 155·8 (6·0) | 156·1 (6·3) |
| **Maternal MUAC in cm, mean (SD)** | 27·5 (3·9) | 27·6 (4·0) | 27·5 (3·9) |
| **Highest level of schooling completed, n/N (%)** | | | |
| None | 128/1189 (11%) | 65/596 (11%) | 63/593 (11%) |
| Primary | 702/1189 (59%) | 366/596 (61%) | 336/593 (57%) |
| Secondary | 326/1189 (27%) | 147/596 (25%) | 179/593 (30%) |
| Higher | 33/1189 (3%) | 18/596 (3%) | 15/593 (3%) |
| **Wealth index tertiles, n/N (%)** | | | |
| Lowest tertile | 336/1192 (28%) | 177/597 (30%) | 159/595 (27%) |
| Middle tertile | 415/1192 (35%) | 200/597 (34%) | 215/595 (36%) |
| Highest tertile | 441/1192 (37%) | 220/597 (37%) | 221/595 (37%) |
| **Slept under a bed net last night, n/N (%)** | 932/956 (97%) | 451/466 (97%) | 481/490 (98%) |
| **Microscopy positivity, n/N (%)** | 70/1192 (6%) | 38/597 (6%) | 32/595 (5%) |
| **PCR/LAMP positivity, n/N (%)** | 114/1082 (11%) | 56/543 (10%) | 58/539 (11%) |

Abbreviations: LAMP = loop-mediated isothermal amplification method MUAC = mid-upper arm circumference; PCR = polymerase chain reaction

Table S-8. Gutman et al, unpublished

| Enrolment characteristics | Total N=593 | IPTp-SP N=297 | IPTp-DP N=296 |
|---|---|---|---|
| **Maternal age in years, mean (SD)** | 24·6 (6·2) | 24·5 (6·4) | 24·7 (6·1) |
| **Gestational age in weeks, mean (SD)** | 20·1 (3·2) | 20·1 (3·2) | 20·1 (3·1) |
| **Gravidity categories, n/N (%)** | | | |
| Primigravidae | 184/589 (31%) | 102/295 (35%) | 82/294 (28%) |
| Secundigravidae | 146/589 (25%) | 73/295 (25%) | 73/294 (25%) |
| Multigravidae (3+) | 259/589 (44%) | 120/295 (41%) | 139/294 (47%) |
| **Weight in kg, mean (SD)** | 58·4 (10·0) | 58·6 (9·4) | 58·3 (10·6) |
| **Height in cm, mean (SD)** | 157·4 (6·2) | 157·3 (6·6) | 157·5 (5·7) |
| **Maternal MUAC in cm, mean (SD)** | 26·2 (3·1) | 26·4 (3·2) | 26·1 (3·1) |
| **Maternal education level, n/N (%)** | | | |
| None | 286/593 (48%) | 146/297 (49%) | 140/296 (47%) |
| Primary | 209/593 (35%) | 97/297 (33%) | 112/296 (38%) |
| Secondary | 89/593 (15%) | 49/297 (16%) | 40/296 (14%) |
| Higher | 9/593 (2%) | 5/297 (2%) | 4/296 (1%) |
| **Wealth index tertiles, n/N (%)** | | | |
| Lowest tertile | 200/593 (34%) | 93/297 (31%) | 107/296 (36%) |
| Middle tertile | 196/593 (33%) | 105/297 (35%) | 91/296 (31%) |
| Highest tertile | 197/593 (33%) | 99/297 (33%) | 98/296 (33%) |
| **Slept under a bed net last night, n/N (%)** | 932/956 (97%) | 451/466 (97%) | 481/490 (98%) |
| **Microscopy positivity, n/N (%)** | -- | -- | -- |
| **PCR/LAMP positivity, n/N (%)** | 143/593 (24%) | 66/297 (22%) | 77/296 (26%) |

Abbreviations: LAMP = loop-mediated isothermal amplification method MUAC = mid-upper arm circumference; PCR = polymerase chain reaction

### Appendix 7. Forest plot of study-specific estimates

#### Figure S-1. Any composite adverse birth outcome

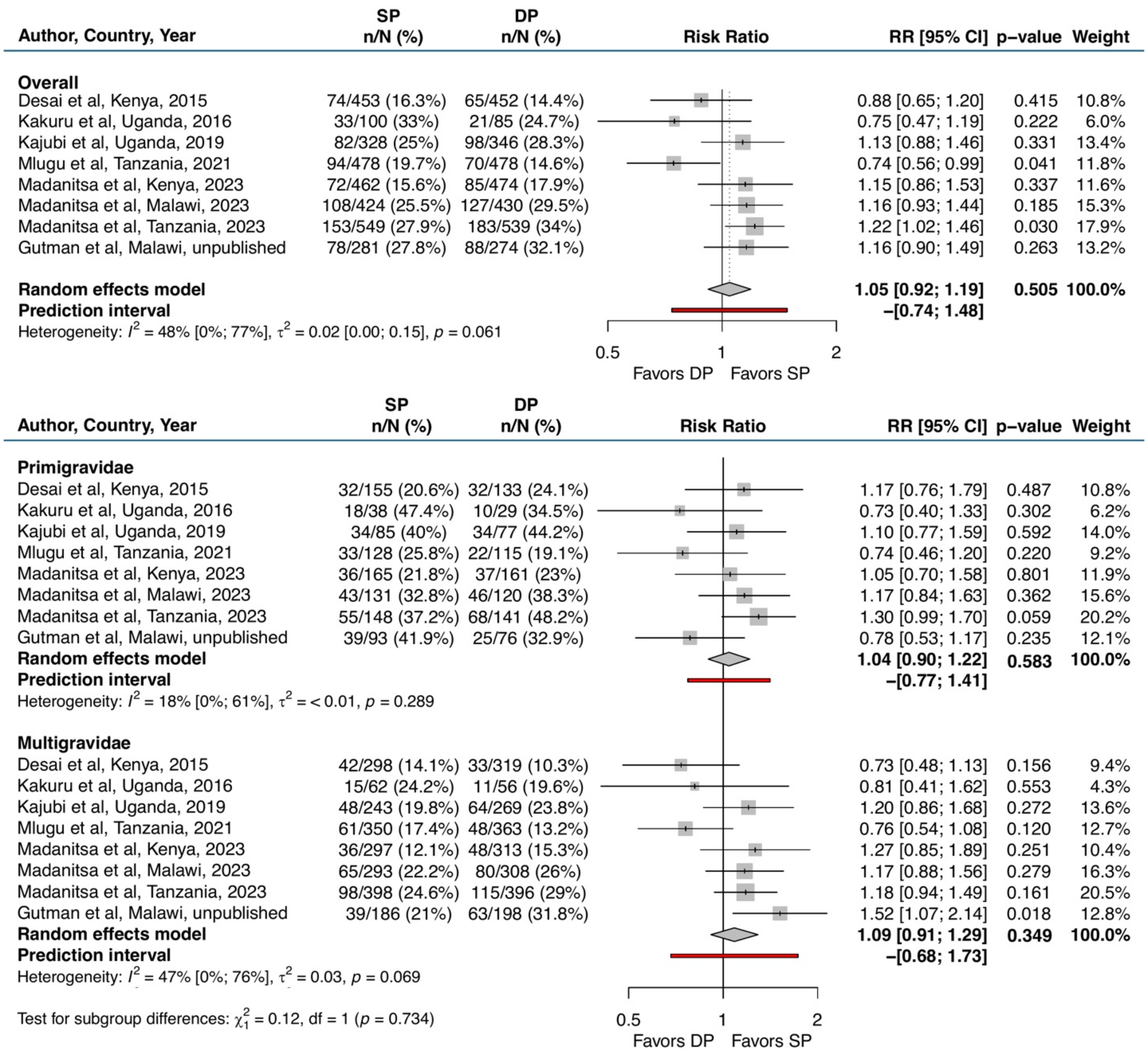

Abbreviations: CI = confidence interval; DP = dihydroartemisinin-piperaquine; LBW = low birthweight; RR = risk ratio; SGA = small-for-gestational age; SP = sulfadoxine-pyrimethamine

### Figure S-2. Foetal loss

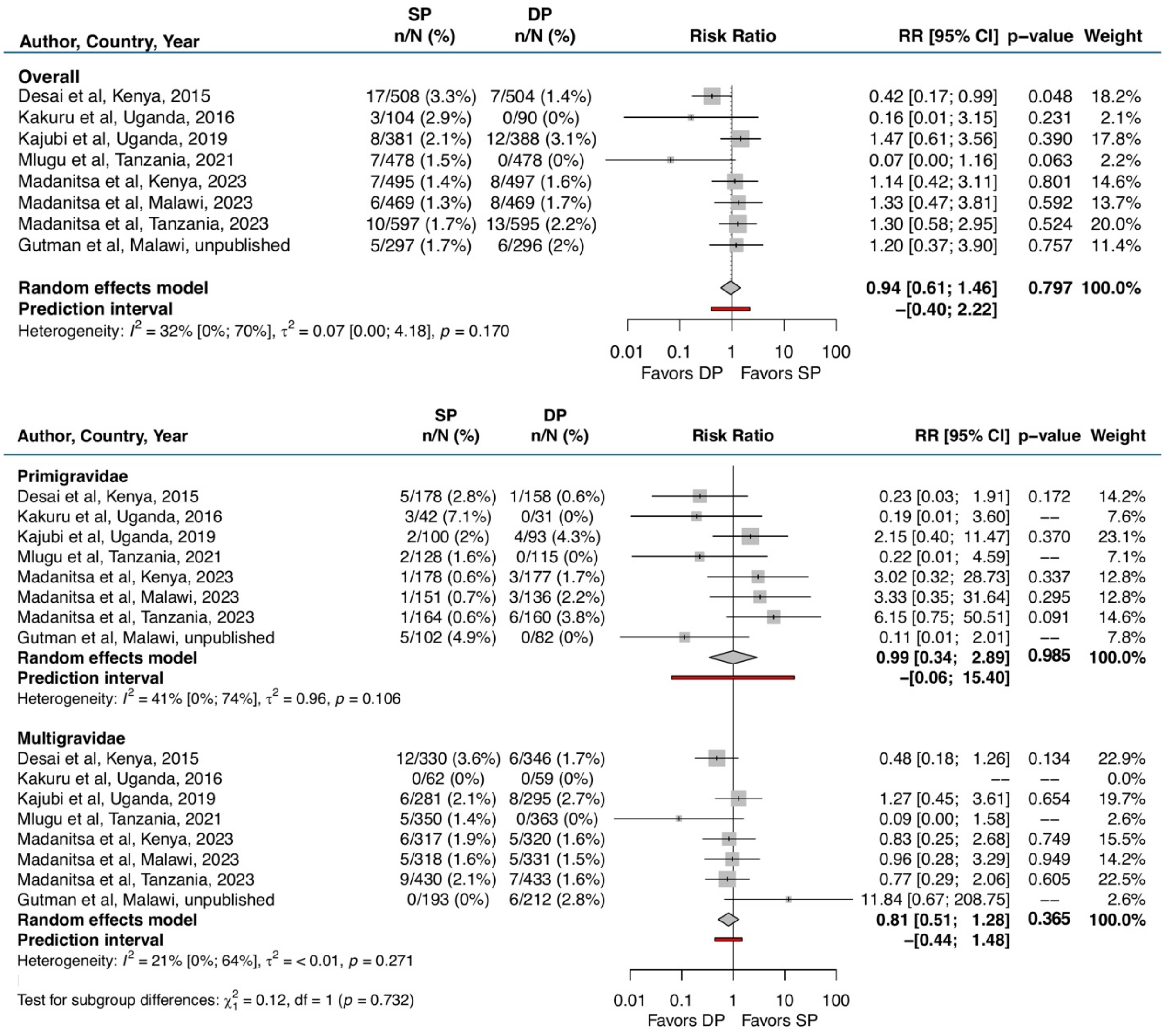

Abbreviations: CI = confidence interval; DP = dihydroartemisinin-piperaquine; RR = risk ratio; SP = sulfadoxine-pyrimethamine

Figure S-3. Small-for-gestational age (<10th percentile for birthweight-for-gestational age)

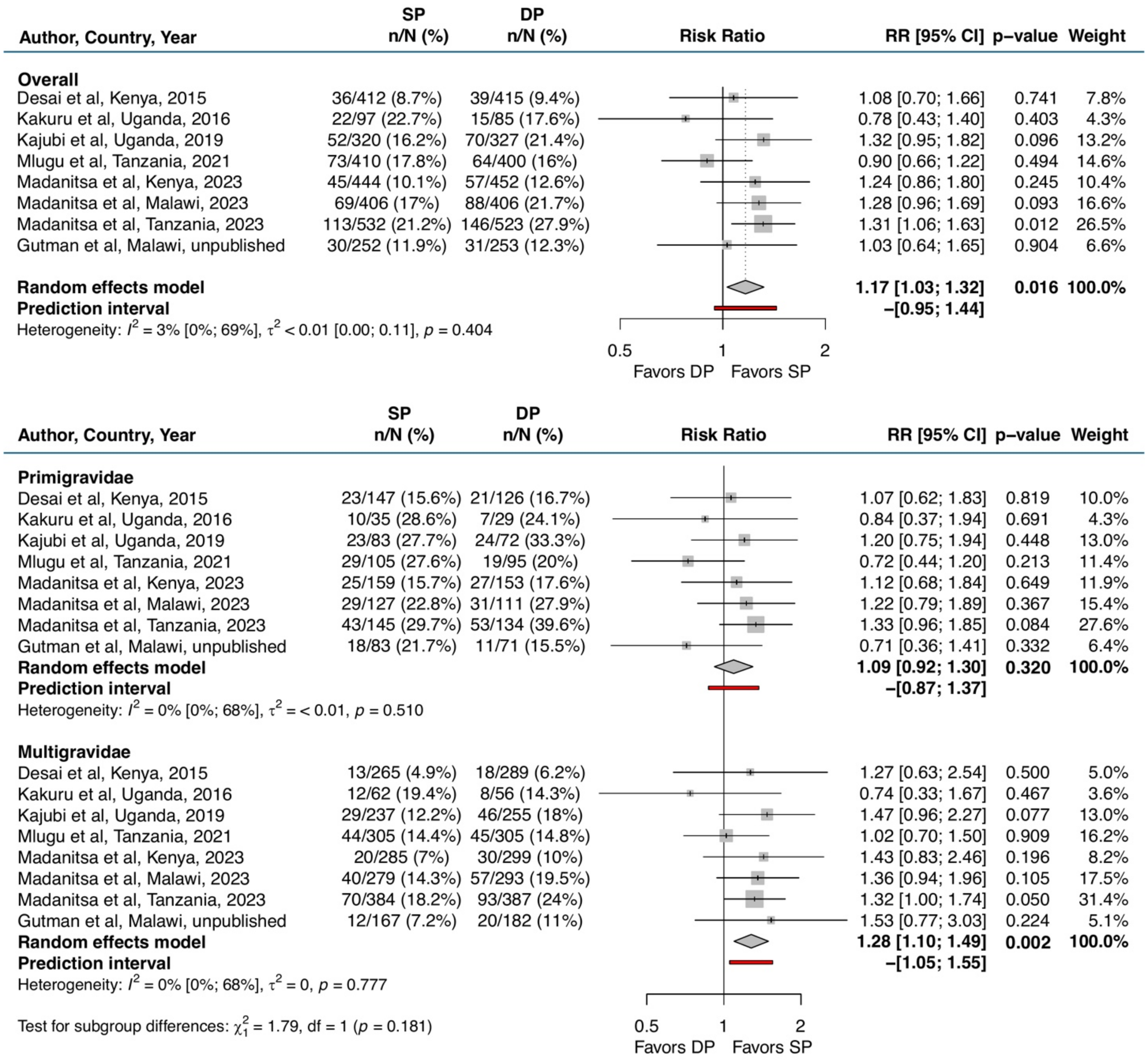

Abbreviations: CI = confidence interval; DP = dihydroartemisinin-piperaquine; RR = risk ratio; SP = sulfadoxine-pyrimethamine

### Figure S-4. Preterm birth (<37 gestational weeks)

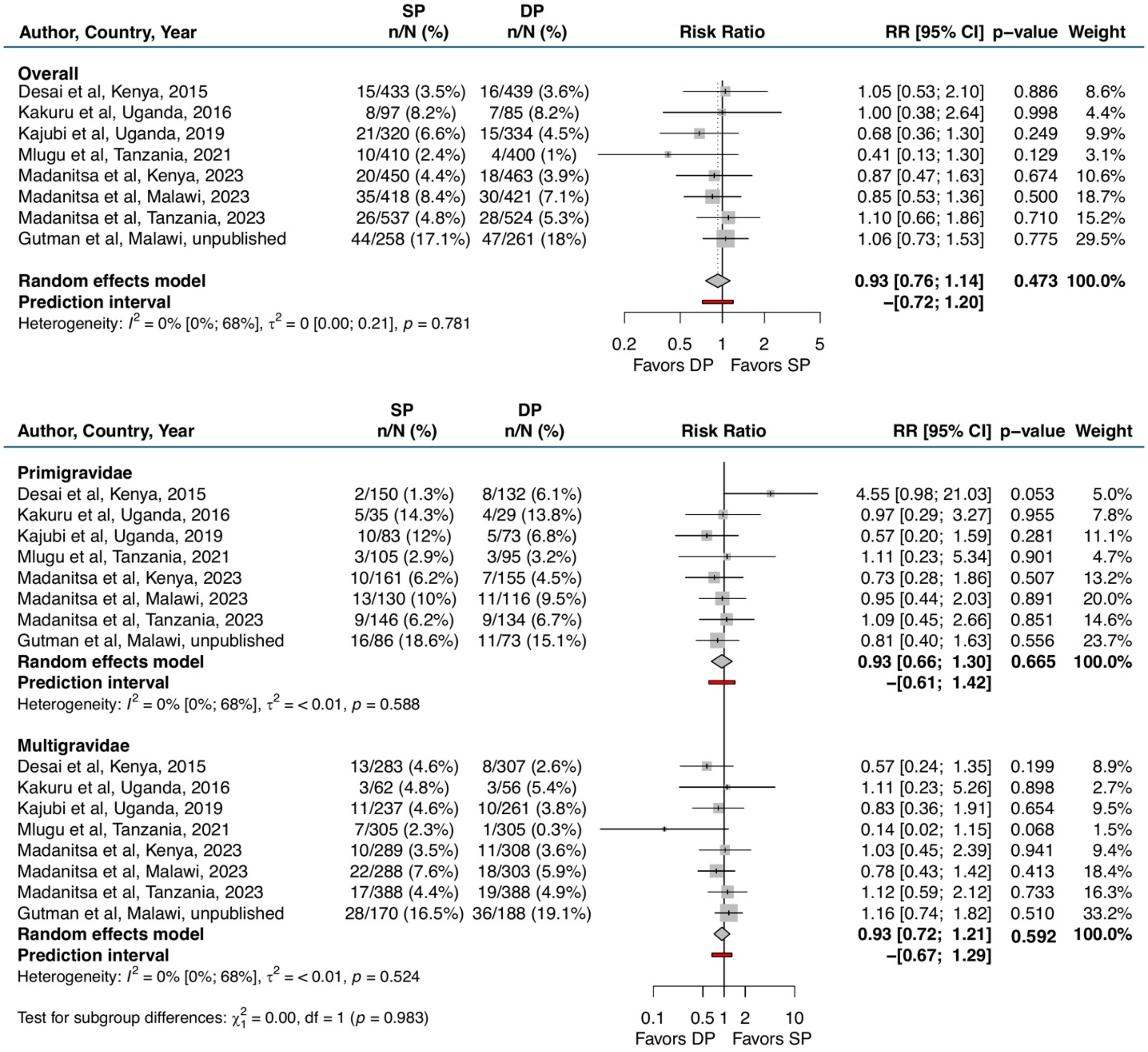

Abbreviations: CI = confidence interval; DP = dihydroartemisinin-piperaquine; RR = risk ratio; SP = sulfadoxine-pyrimethamine

### Figure S-5. Low birthweight

| Author, Country, Year | SP n/N (%) | DP n/N (%) | Risk Ratio | RR [95% CI] | p-value | Weight |
|---|---|---|---|---|---|---|
| **Overall** | | | | | | |
| Desai et al, Kenya, 2015 | 19/412 (4.6%) | 22/415 (5.3%) | | 1.15 [0.63; 2.09] | 0.648 | 11.3% |
| Kakuru et al, Uganda, 2016 | 12/97 (12.4%) | 10/85 (11.8%) | | 0.95 [0.43; 2.09] | 0.900 | 8.1% |
| Kajubi et al, Uganda, 2019 | 21/320 (6.6%) | 22/327 (6.7%) | | 1.03 [0.58; 1.83] | 0.933 | 11.8% |
| Mlugu et al, Tanzania, 2021 | 44/410 (10.7%) | 22/400 (5.5%) | | 0.51 [0.31; 0.84] | 0.008 | 13.8% |
| Madanitsa et al, Kenya, 2023 | 19/444 (4.3%) | 22/452 (4.9%) | | 1.14 [0.62; 2.07] | 0.674 | 11.3% |
| Madanitsa et al, Malawi, 2023 | 31/407 (7.6%) | 50/408 (12.3%) | | 1.61 [1.05; 2.46] | 0.029 | 15.5% |
| Madanitsa et al, Tanzania, 2023 | 41/532 (7.7%) | 57/523 (10.9%) | | 1.41 [0.96; 2.07] | 0.076 | 16.8% |
| Gutman et al, Malawi, unpublished | 19/253 (7.5%) | 22/255 (8.6%) | | 1.15 [0.64; 2.07] | 0.644 | 11.5% |
| **Random effects model** | | | | **1.09 [0.83; 1.43]** | **0.537** | **100.0%** |
| **Prediction interval** | | | | **–[0.51; 2.31]** | | |

Heterogeneity: $I^2 = 50\%$ [0%; 78%], $\tau^2 = 0.08$ [0.00; 0.42], $p = 0.049$

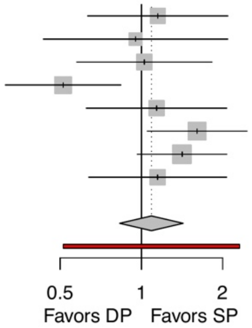

| Author, Country, Year | SP n/N (%) | DP n/N (%) | Risk Ratio | RR [95% CI] | p-value | Weight |
|---|---|---|---|---|---|---|
| **Primigravidae** | | | | | | |
| Desai et al, Kenya, 2015 | 12/147 (8.2%) | 13/126 (10.3%) | | 1.26 [0.60; 2.67] | 0.539 | 14.3% |
| Kakuru et al, Uganda, 2016 | 8/35 (22.9%) | 4/29 (13.8%) | | 0.60 [0.20; 1.80] | 0.366 | 8.6% |
| Kajubi et al, Uganda, 2019 | 11/83 (13.3%) | 4/72 (5.6%) | | 0.42 [0.14; 1.26] | 0.121 | 8.5% |
| Mlugu et al, Tanzania, 2021 | 14/105 (13.3%) | 5/95 (5.3%) | | 0.39 [0.15; 1.05] | 0.064 | 10.0% |
| Madanitsa et al, Kenya, 2023 | 8/159 (5%) | 11/153 (7.2%) | | 1.43 [0.59; 3.46] | 0.428 | 11.6% |
| Madanitsa et al, Malawi, 2023 | 16/127 (12.6%) | 18/112 (16.1%) | | 1.28 [0.68; 2.38] | 0.444 | 17.5% |
| Madanitsa et al, Tanzania, 2023 | 16/145 (11%) | 16/134 (11.9%) | | 1.08 [0.56; 2.08] | 0.813 | 16.7% |
| Gutman et al, Malawi, unpublished | 13/84 (15.5%) | 8/72 (11.1%) | | 0.72 [0.32; 1.63] | 0.430 | 12.8% |
| **Random effects model** | | | | **0.92 [0.68; 1.24]** | **0.578** | **100.0%** |
| **Prediction interval** | | | | **–[0.56; 1.51]** | | |
| Heterogeneity: $I^2 = 19\%$ [0%; 62%], $\tau^2 = 0.02$, $p = 0.278$ | | | | | | |
| **Multigravidae** | | | | | | |
| Desai et al, Kenya, 2015 | 7/265 (2.6%) | 9/289 (3.1%) | | 1.18 [0.45; 3.12] | 0.740 | 8.9% |
| Kakuru et al, Uganda, 2016 | 4/62 (6.5%) | 6/56 (10.7%) | | 1.66 [0.49; 5.58] | 0.412 | 6.4% |
| Kajubi et al, Uganda, 2019 | 10/237 (4.2%) | 18/255 (7.1%) | | 1.67 [0.79; 3.55] | 0.180 | 12.4% |
| Mlugu et al, Tanzania, 2021 | 30/305 (9.8%) | 17/305 (5.6%) | | 0.57 [0.32; 1.01] | 0.052 | 16.5% |
| Madanitsa et al, Kenya, 2023 | 11/285 (3.9%) | 11/299 (3.7%) | | 0.95 [0.42; 2.16] | 0.909 | 11.2% |
| Madanitsa et al, Malawi, 2023 | 15/280 (5.4%) | 32/294 (10.9%) | | 2.03 [1.13; 3.67] | 0.019 | 16.0% |
| Madanitsa et al, Tanzania, 2023 | 25/384 (6.5%) | 41/387 (10.6%) | | 1.63 [1.01; 2.62] | 0.045 | 19.2% |
| Gutman et al, Malawi, unpublished | 6/167 (3.6%) | 14/183 (7.7%) | | 2.13 [0.84; 5.41] | 0.112 | 9.4% |
| **Random effects model** | | | | **1.34 [0.93; 1.92]** | **0.115** | **100.0%** |
| **Prediction interval** | | | | **–[0.50; 3.57]** | | |
| Heterogeneity: $I^2 = 47\%$ [0%; 77%], $\tau^2 = 0.13$, $p = 0.065$ | | | | | | |

Test for subgroup differences: $\chi^2_1 = 2.45$, df = 1 ($p = 0.118$)

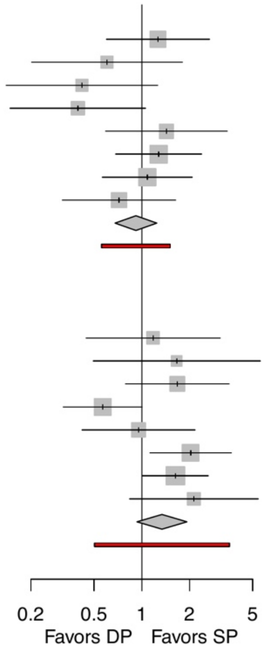

Abbreviations: CI = confidence interval; DP = dihydroartemisinin-piperaquine; RR = risk ratio; SP = sulfadoxine-pyrimethamine

### Figure S-6. Neonatal death

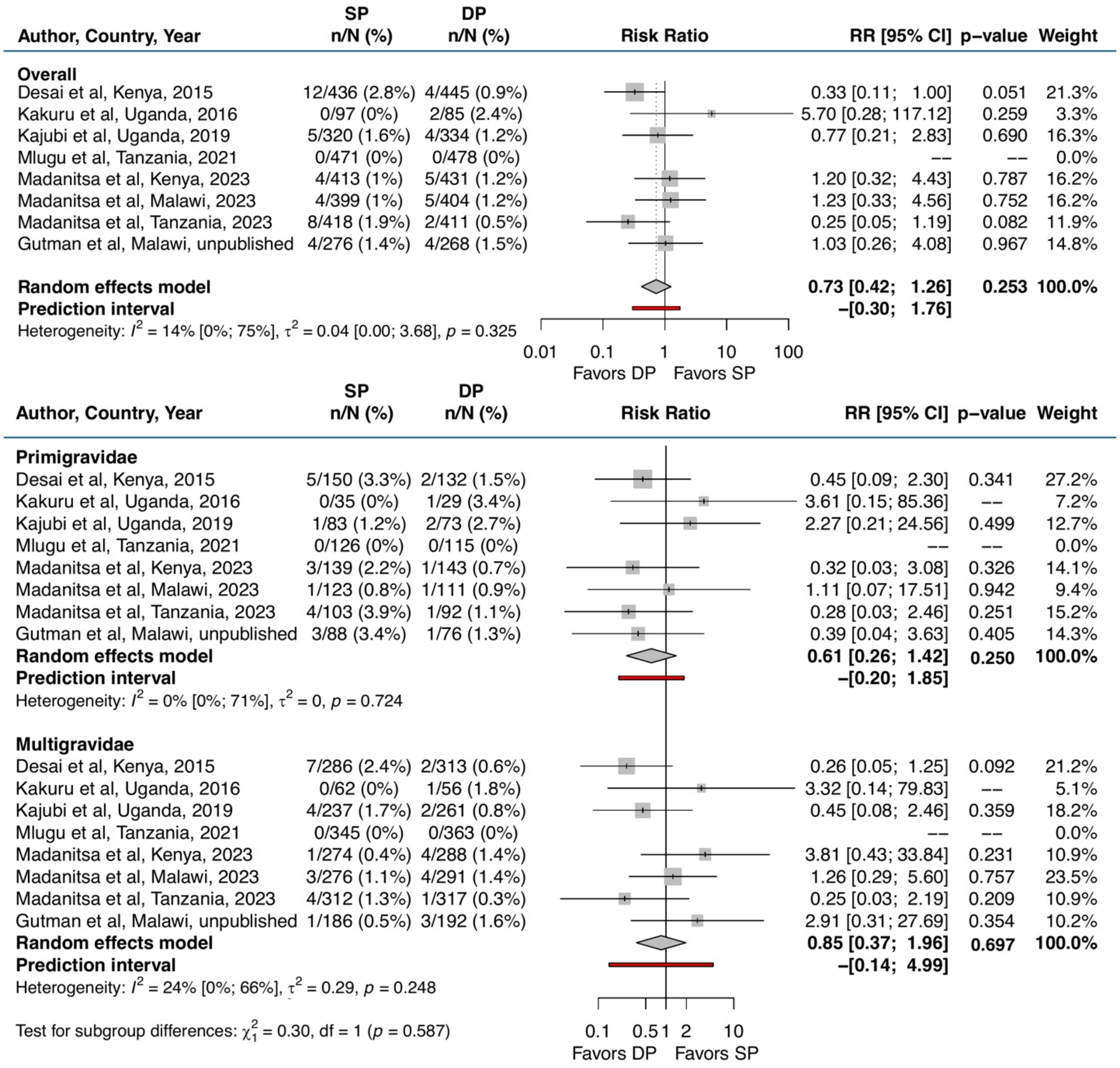

Abbreviations: CI = confidence interval; DP = dihydroartemisinin-piperaquine; RR = risk ratio; SP = sulfadoxine-pyrimethamine

### Figure S-7. Mean birthweight in grams

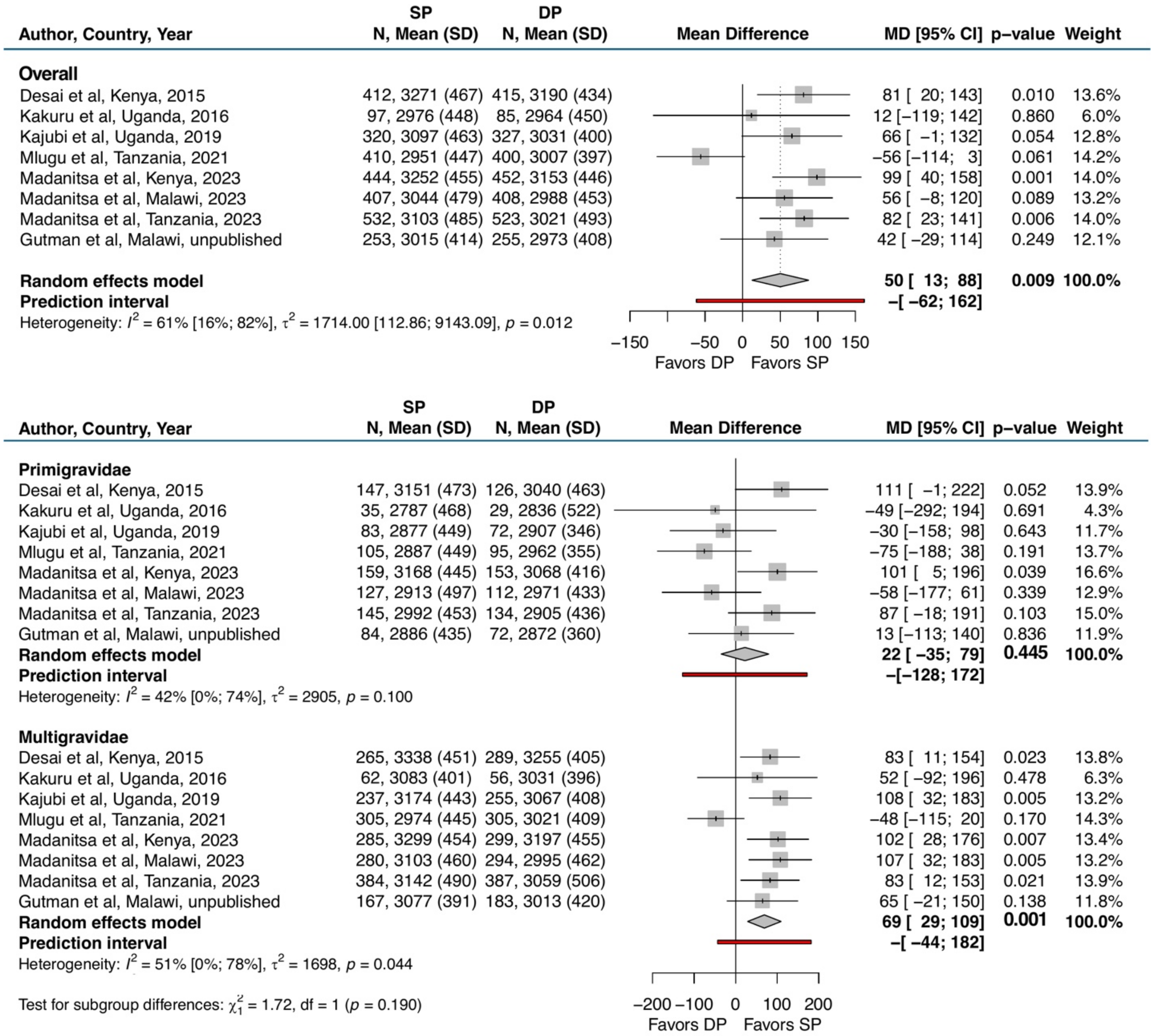

Abbreviations: CI = confidence interval; DP = dihydroartemisinin-piperaquine; MD = mean difference; SD = standard deviation; SP = sulfadoxine-pyrimethamine

Figure S-8. Mean gestational age at birth in weeks

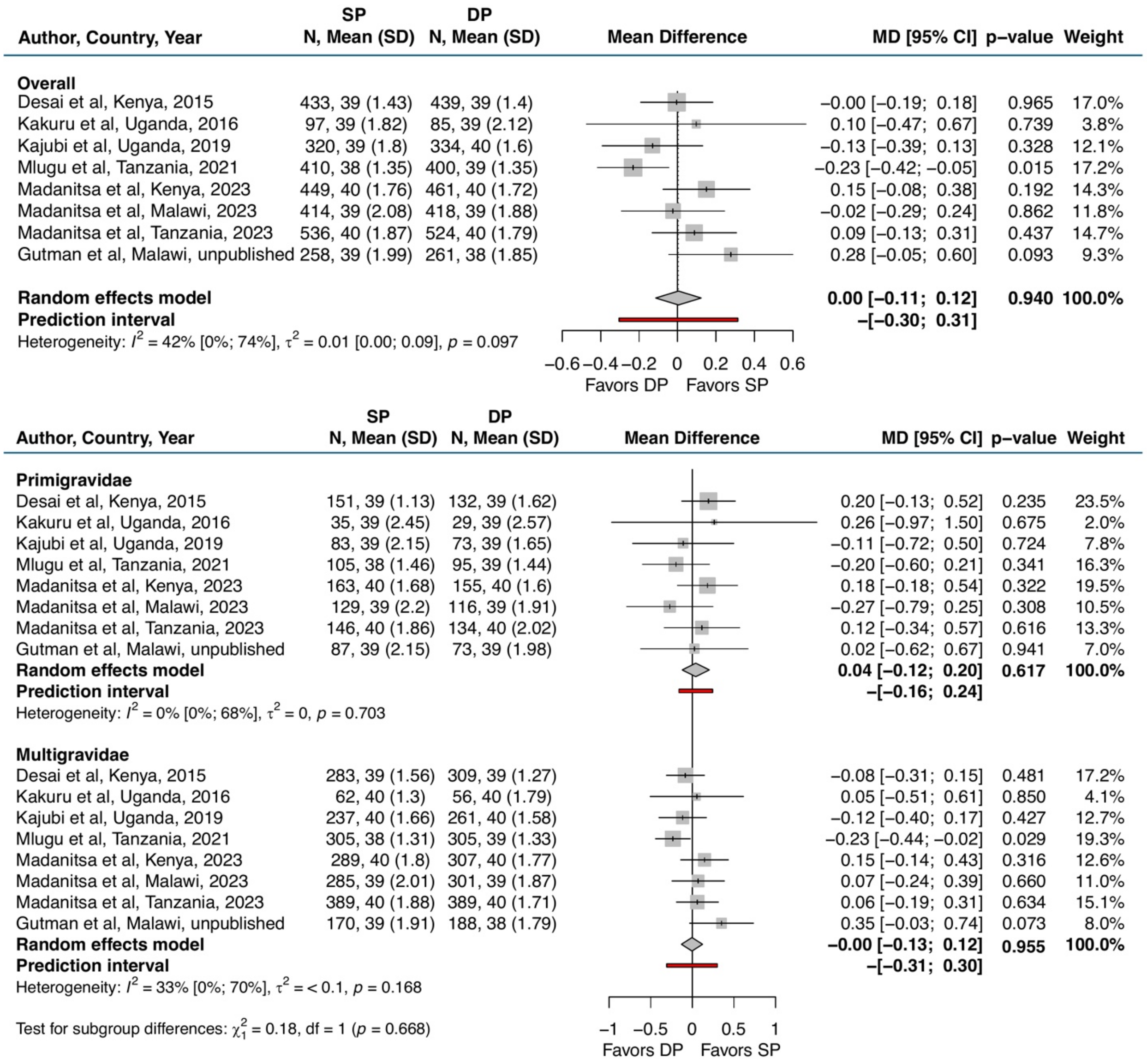

Abbreviations: CI = confidence interval; DP = dihydroartemisinin-piperaquine; MD = mean difference; SD = standard deviation; SP = sulfadoxine-pyrimethamine

### Figure S-9. Mean birthweight-for-gestational age z-scores

| Author, Country, Year | SP N, Mean (SD) | DP N, Mean (SD) | MD [95% CI] | p–value | Weight |
|---|---|---|---|---|---|
| **Overall** | | | | | |
| Desai et al, Kenya, 2015 | 412, 0.13 (1.02) | 415, −0.07 (0.94) | 0.20 [ 0.07; 0.33] | 0.003 | 13.9% |
| Kakuru et al, Uganda, 2016 | 97, −0.59 (0.89) | 85, −0.56 (0.82) | −0.03 [−0.28; 0.22] | 0.814 | 6.6% |
| Kajubi et al, Uganda, 2019 | 320, −0.35 (0.96) | 327, −0.55 (0.92) | 0.20 [ 0.05; 0.34] | 0.008 | 12.9% |
| Mlugu et al, Tanzania, 2021 | 410, −0.33 (1.07) | 400, −0.28 (1.02) | −0.05 [−0.20; 0.09] | 0.458 | 13.0% |
| Madanitsa et al, Kenya, 2023 | 444, −0.1 (0.92) | 452, −0.29 (0.9) | 0.20 [ 0.08; 0.32] | 0.001 | 15.3% |
| Madanitsa et al, Malawi, 2023 | 406, −0.36 (1.06) | 406, −0.51 (1.06) | 0.15 [ 0.00; 0.29] | 0.045 | 12.8% |
| Madanitsa et al, Tanzania, 2023 | 532, −0.44 (1.01) | 523, −0.65 (1.06) | 0.20 [ 0.08; 0.33] | 0.001 | 14.7% |
| Gutman et al, Malawi, unpublished | 252, −0.2 (0.96) | 253, −0.21 (0.99) | 0.01 [−0.16; 0.18] | 0.937 | 10.9% |
| **Random effects model** | | | **0.12 [ 0.05; 0.20]** | **0.001** | **100.0%** |
| **Prediction interval** | | | **–[−0.09; 0.33]** | | |

Heterogeneity: $I^2$ = 51% [0%; 78%], $\tau^2$ < 0.01 [0.00; 0.05], $p$ = 0.045

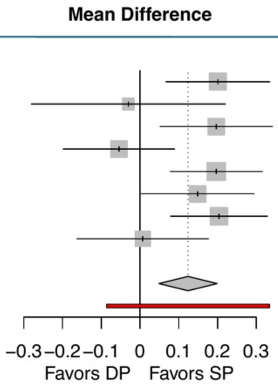

| Author, Country, Year | SP N, Mean (SD) | DP N, Mean (SD) | MD [95% CI] | p–value | Weight |
|---|---|---|---|---|---|
| **Primigravidae** | | | | | |
| Desai et al, Kenya, 2015 | 147, −0.13 (1.07) | 126, −0.31 (0.97) | 0.18 [−0.06; 0.42] | 0.149 | 14.5% |
| Kakuru et al, Uganda, 2016 | 35, −0.84 (0.87) | 29, −0.67 (0.66) | −0.17 [−0.55; 0.22] | 0.397 | 6.8% |
| Kajubi et al, Uganda, 2019 | 83, −0.78 (0.89) | 72, −0.78 (0.95) | −0.00 [−0.29; 0.29] | 0.999 | 11.0% |
| Mlugu et al, Tanzania, 2021 | 105, −0.51 (1.15) | 95, −0.36 (0.97) | −0.15 [−0.44; 0.15] | 0.328 | 10.6% |
| Madanitsa et al, Kenya, 2023 | 159, −0.3 (0.91) | 153, −0.5 (0.95) | 0.20 [−0.01; 0.40] | 0.063 | 18.4% |
| Madanitsa et al, Malawi, 2023 | 127, −0.6 (1.03) | 111, −0.59 (1.18) | −0.00 [−0.29; 0.28] | 0.974 | 11.6% |
| Madanitsa et al, Tanzania, 2023 | 145, −0.74 (0.92) | 134, −0.94 (0.97) | 0.20 [−0.02; 0.42] | 0.072 | 16.7% |
| Gutman et al, Malawi, unpublished | 83, −0.55 (1) | 71, −0.53 (0.89) | −0.02 [−0.32; 0.28] | 0.879 | 10.3% |
| **Random effects model** | | | **0.07 [−0.03; 0.17]** | 0.184 | **100.0%** |
| **Prediction interval** | | | **–[−0.12; 0.25]** | | |
| Heterogeneity: $I^2$ = 12% [0%; 72%], $\tau^2$ = < 0.01, $p$ = 0.335 | | | | | |
| **Multigravidae** | | | | | |
| Desai et al, Kenya, 2015 | 265, 0.28 (0.96) | 289, 0.04 (0.92) | 0.24 [ 0.09; 0.40] | 0.002 | 14.2% |
| Kakuru et al, Uganda, 2016 | 62, −0.45 (0.88) | 56, −0.5 (0.89) | 0.05 [−0.27; 0.37] | 0.750 | 5.0% |
| Kajubi et al, Uganda, 2019 | 237, −0.2 (0.94) | 255, −0.48 (0.91) | 0.28 [ 0.12; 0.44] | 0.001 | 13.4% |
| Mlugu et al, Tanzania, 2021 | 305, −0.27 (1.03) | 305, −0.25 (1.03) | −0.02 [−0.18; 0.14] | 0.814 | 13.4% |
| Madanitsa et al, Kenya, 2023 | 285, 0.02 (0.9) | 299, −0.19 (0.85) | 0.21 [ 0.06; 0.35] | 0.004 | 15.7% |
| Madanitsa et al, Malawi, 2023 | 279, −0.25 (1.06) | 293, −0.49 (1) | 0.23 [ 0.07; 0.40] | 0.007 | 12.9% |
| Madanitsa et al, Tanzania, 2023 | 384, −0.34 (1.02) | 387, −0.55 (1.08) | 0.21 [ 0.06; 0.36] | 0.005 | 15.0% |
| Gutman et al, Malawi, unpublished | 167, −0.03 (0.9) | 182, −0.08 (1) | 0.05 [−0.15; 0.25] | 0.639 | 10.4% |
| **Random effects model** | | | **0.17 [ 0.10; 0.25]** | **< 0.001** | **100.0%** |
| **Prediction interval** | | | **–[−0.01; 0.36]** | | |
| Heterogeneity: $I^2$ = 35% [0%; 71%], $\tau^2$ = < 0.01, $p$ = 0.152 | | | | | |

Test for subgroup differences: $\chi^2_1$ = 2.51, df = 1 ($p$ = 0.113)

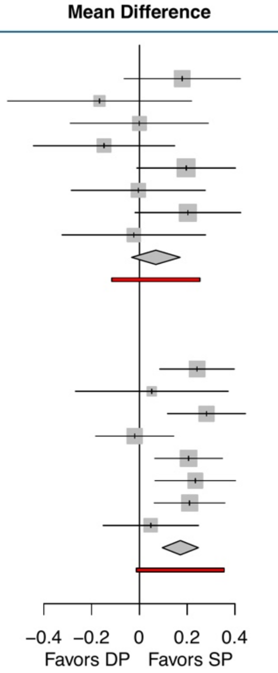

Abbreviations: CI = confidence interval; DP = dihydroartemisinin-piperaquine; MD = mean difference; SD = standard deviation; SP = sulfadoxine-pyrimethamine

### Figure S-10. Incidence of clinical malaria during pregnancy

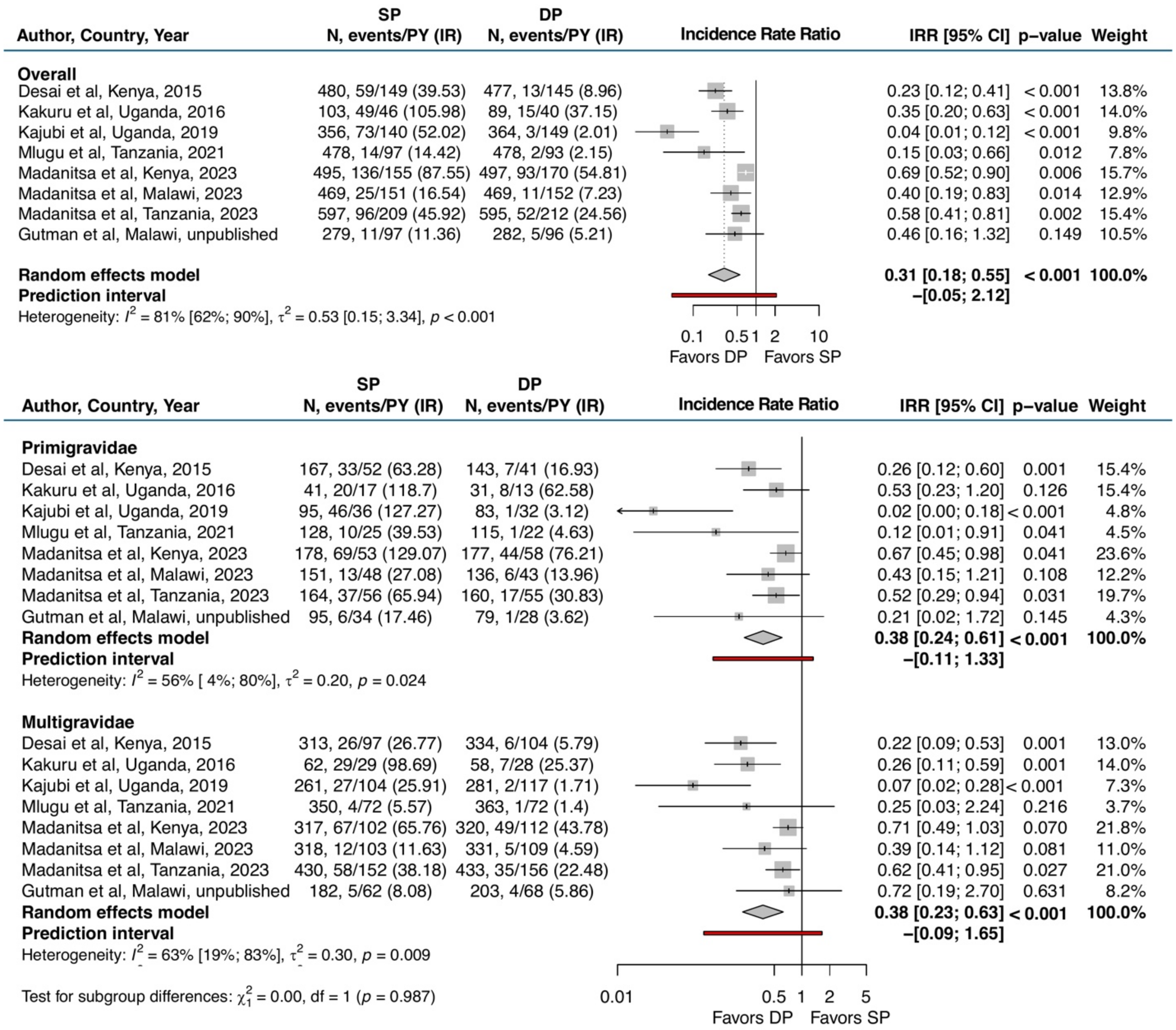

Abbreviations: CI = confidence interval; DP = dihydroartemisinin-piperaquine; IR = incidence rate (episodes per 100 person-years); IRR = incidence rate ratio; PY = person-years; SP = sulfadoxine-pyrimethamine

Figure S-11. Any evidence of pigment in placental tissue by histopathology

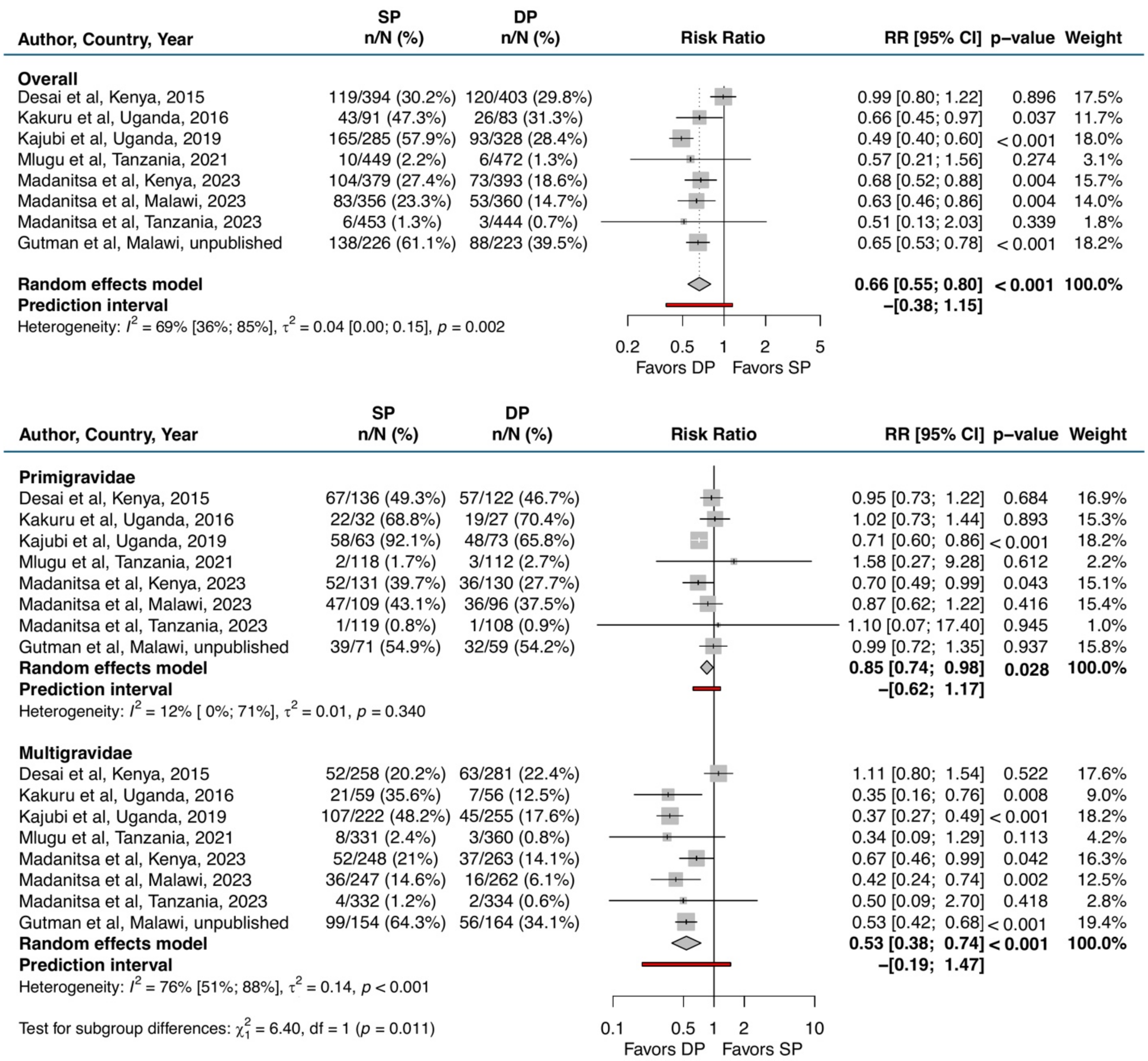

Abbreviations: CI = confidence interval; DP = dihydroartemisinin-piperaquine; RR = risk ratio; SP = sulfadoxine-pyrimethamine

Figure S-12. Any evidence of parasites in placental tissue or blood by histopathology, microscopy, PCR, or RDT

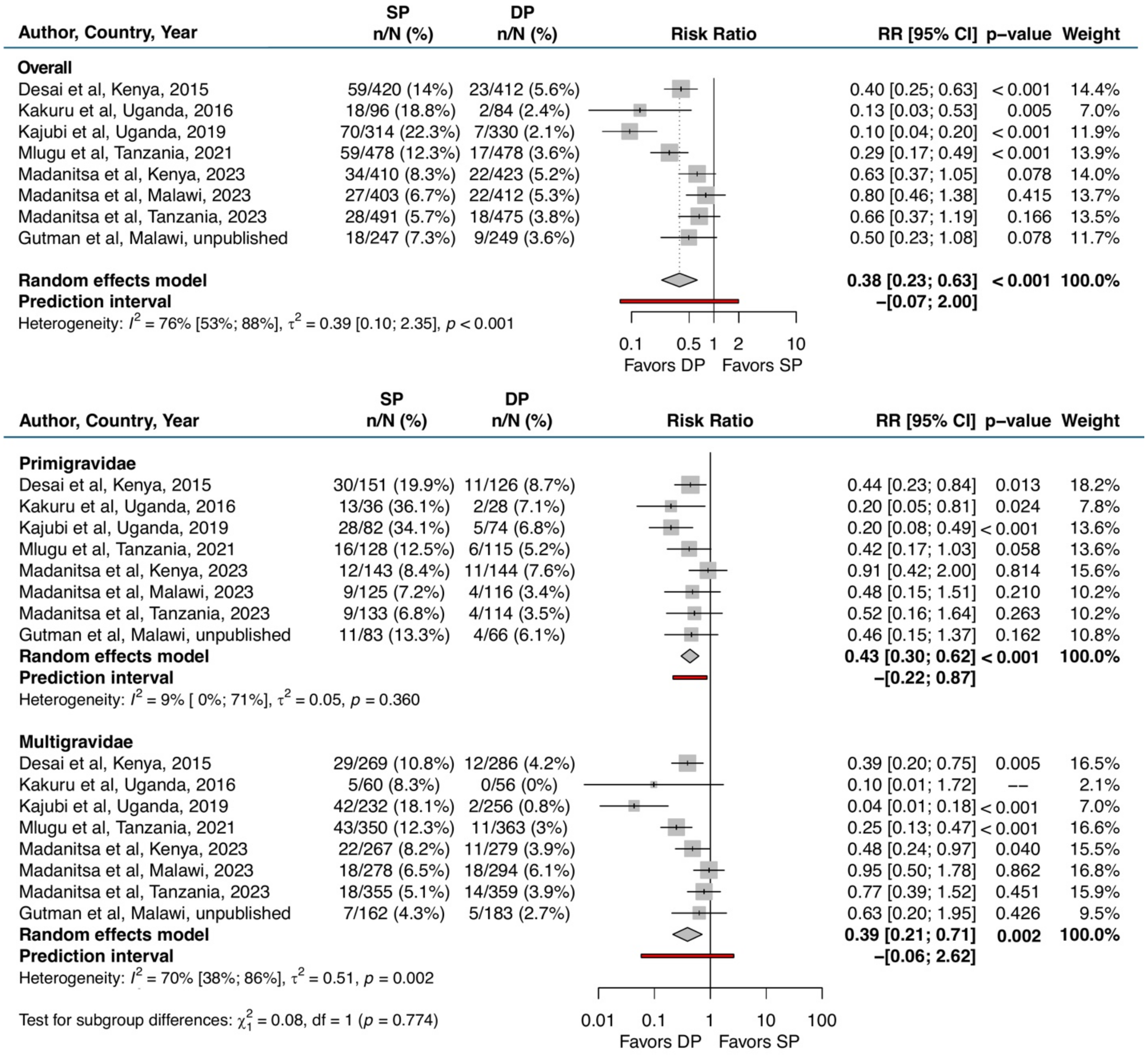

Abbreviations: CI = confidence interval; DP = dihydroartemisinin-piperaquine; PCR = polymerase chain reaction; RDT = rapid diagnostic test; RR = risk ratio; SP = sulfadoxine-pyrimethamine

#### Figure S-13. Any evidence of pigment or parasites in placental tissue or blood by histopathology, microscopy, PCR, or RDT

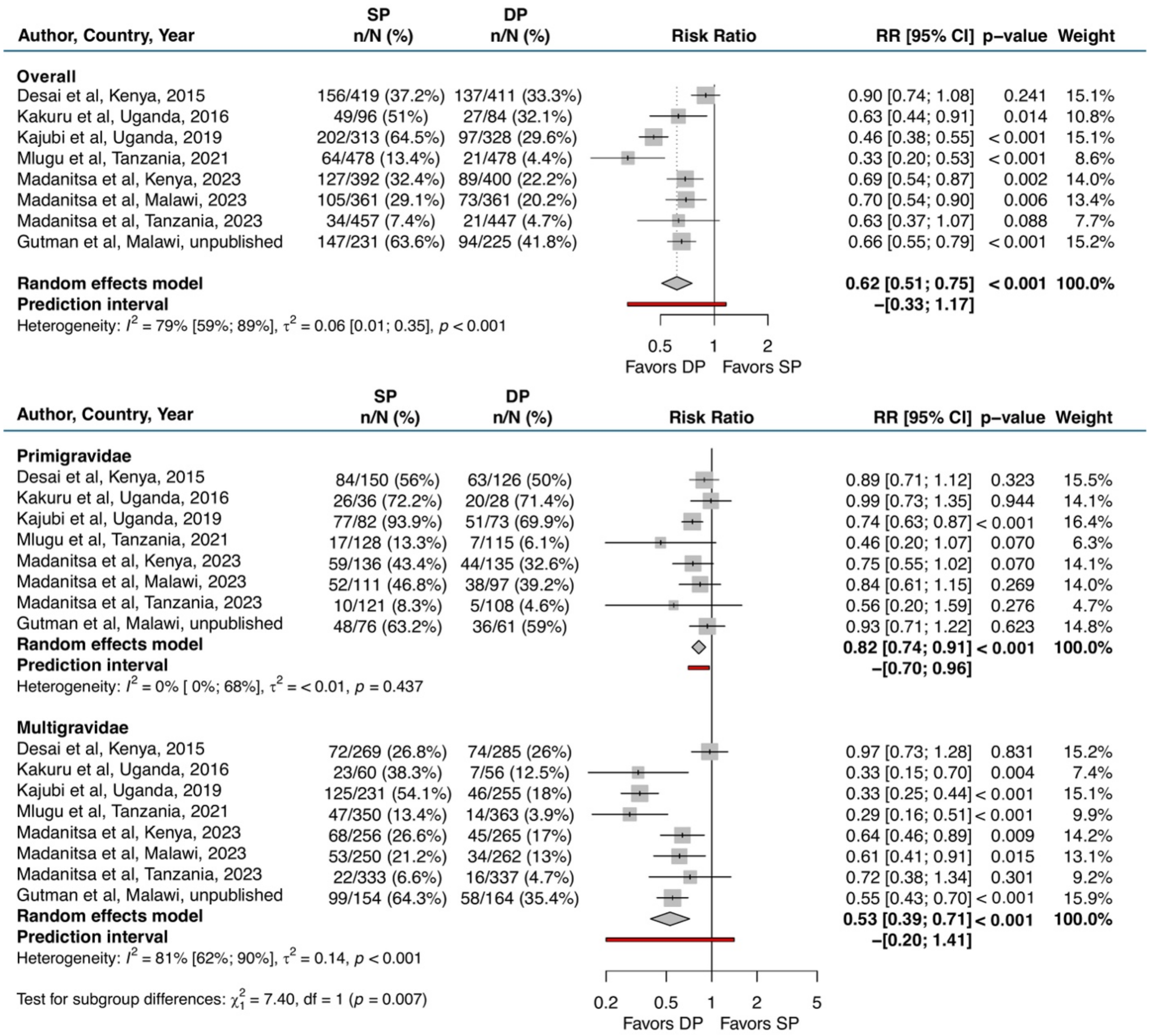

Abbreviations: CI = confidence interval; DP = dihydroartemisinin-piperaquine; PCR = polymerase chain reaction; RDT = rapid diagnostic test; RR = risk ratio; SP = sulfadoxine-pyrimethamine

#### Figure S-14. Any evidence of peripheral malaria at delivery by microscopy, PCR, or RDT

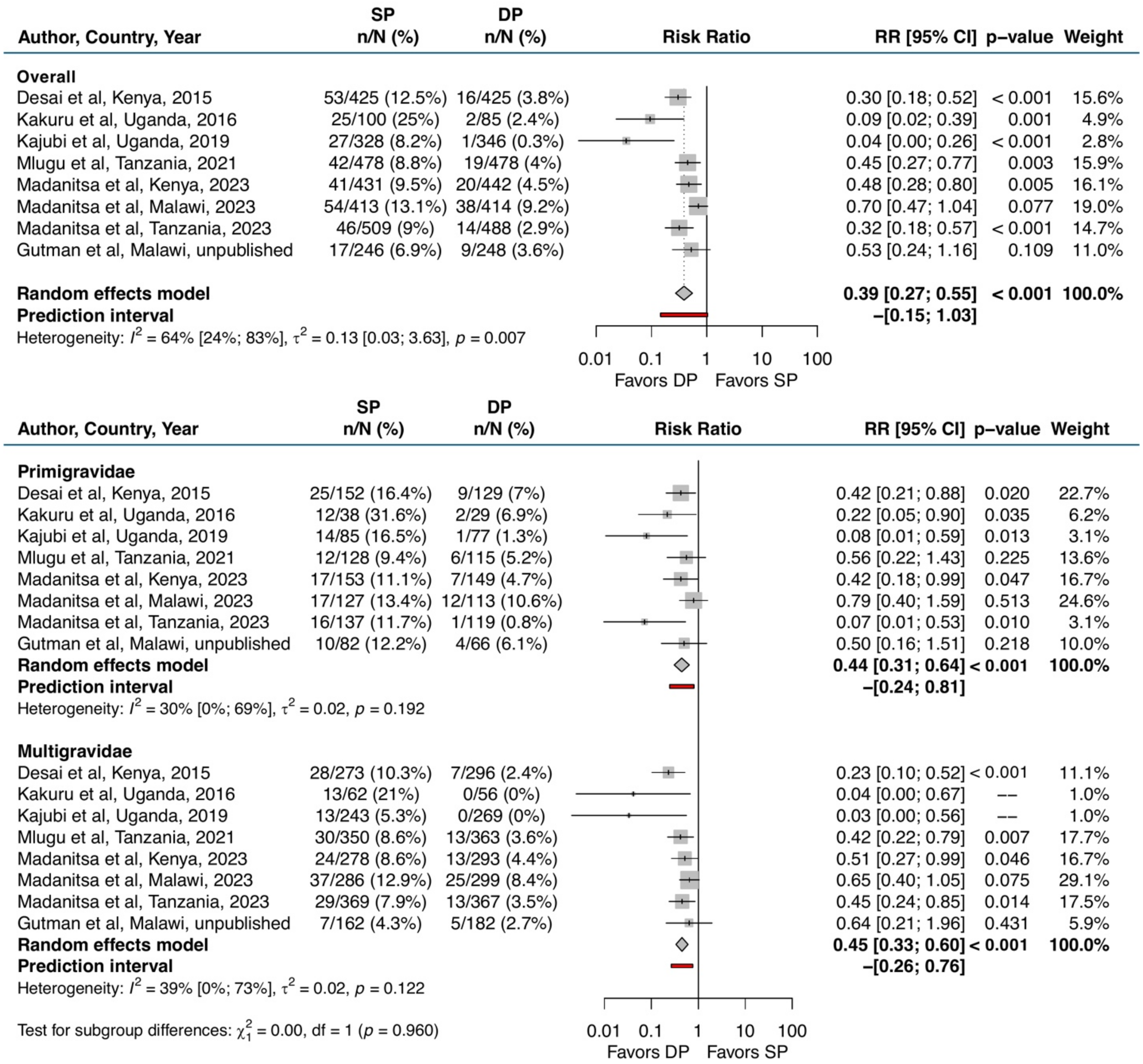

Abbreviations: CI = confidence interval; DP = dihydroartemisinin-piperaquine; PCR = polymerase chain reaction; RDT = rapid diagnostic test; RR = risk ratio; SP = sulfadoxine-pyrimethamine

#### Figure S-15. Any evidence of severe anaemia (Hb <7 g/dL) during pregnancy

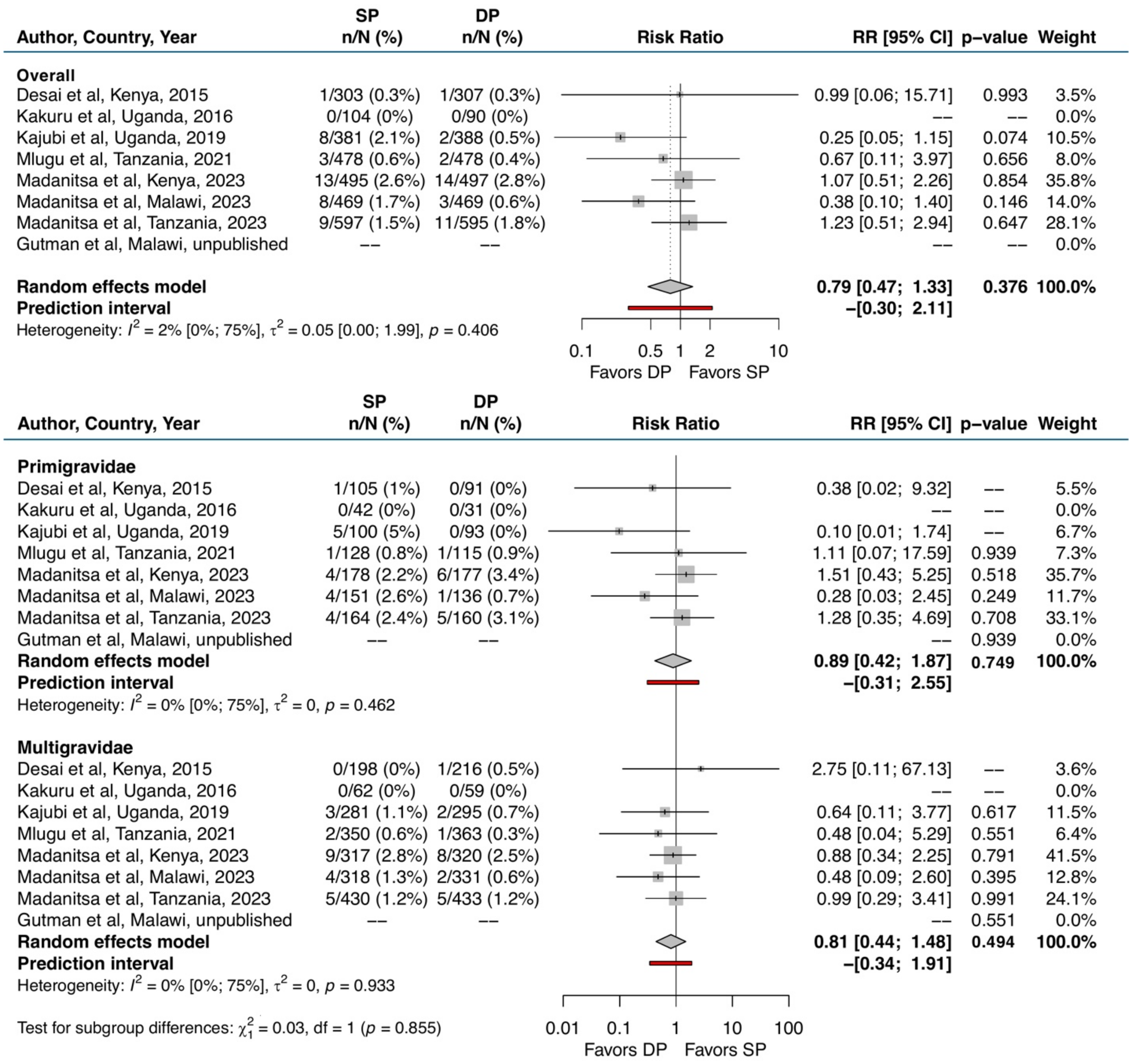

Abbreviations: CI = confidence interval; DP = dihydroartemisinin-piperaquine; g/dL = grams/deciliter; Hb = haemoglobin; RR = risk ratio; SP = sulfadoxine-pyrimethamine

#### Figure S-16. Any evidence of moderate anaemia (Hb <9 g/dL) during pregnancy

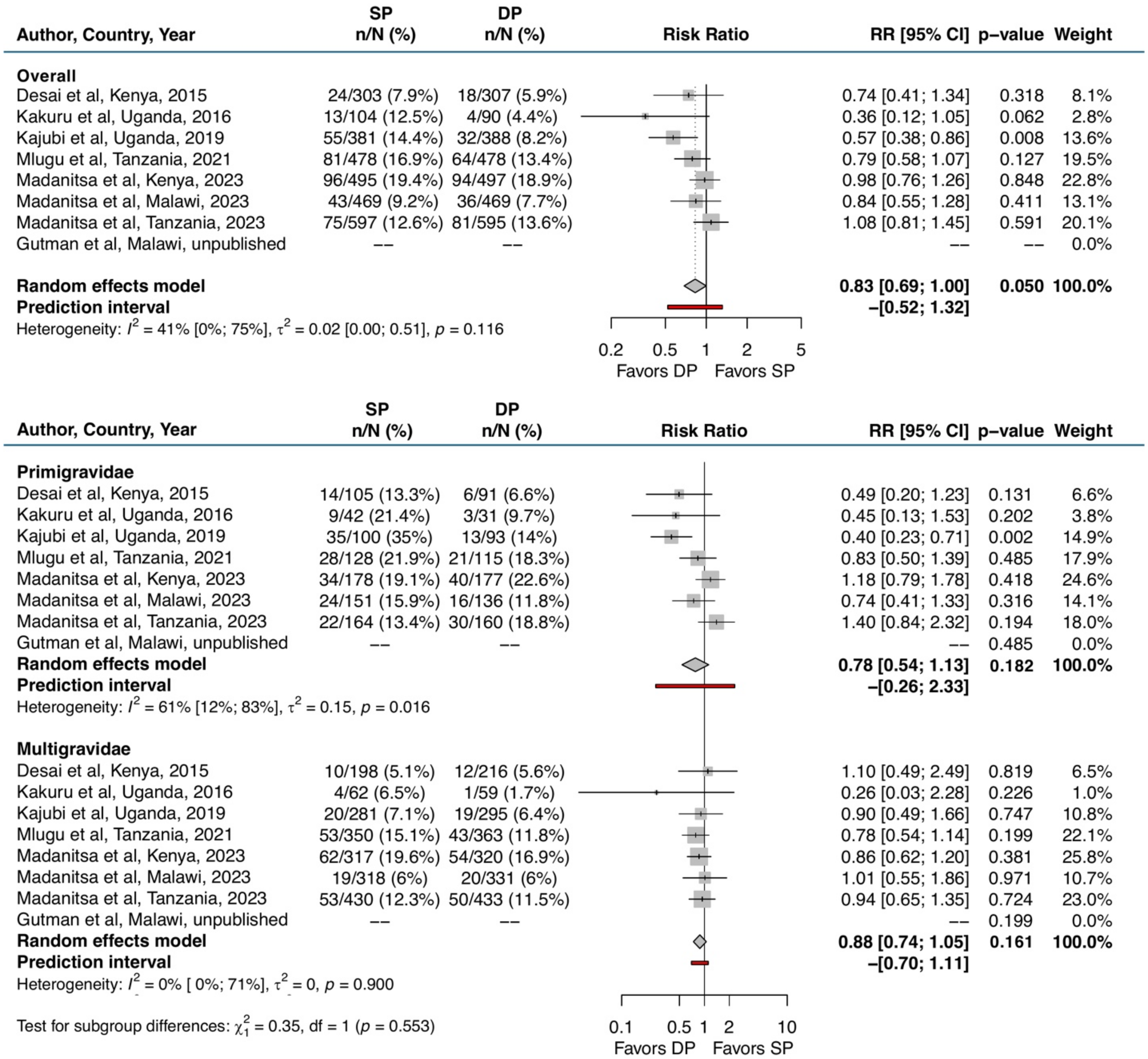

Abbreviations: CI = confidence interval; DP = dihydroartemisinin-piperaquine; g/dL = grams/deciliter; Hb = haemoglobin; RR = risk ratio; SP = sulfadoxine-pyrimethamine

### Figure S-17. Any evidence of mild anaemia (Hb <11 g/dL) during pregnancy

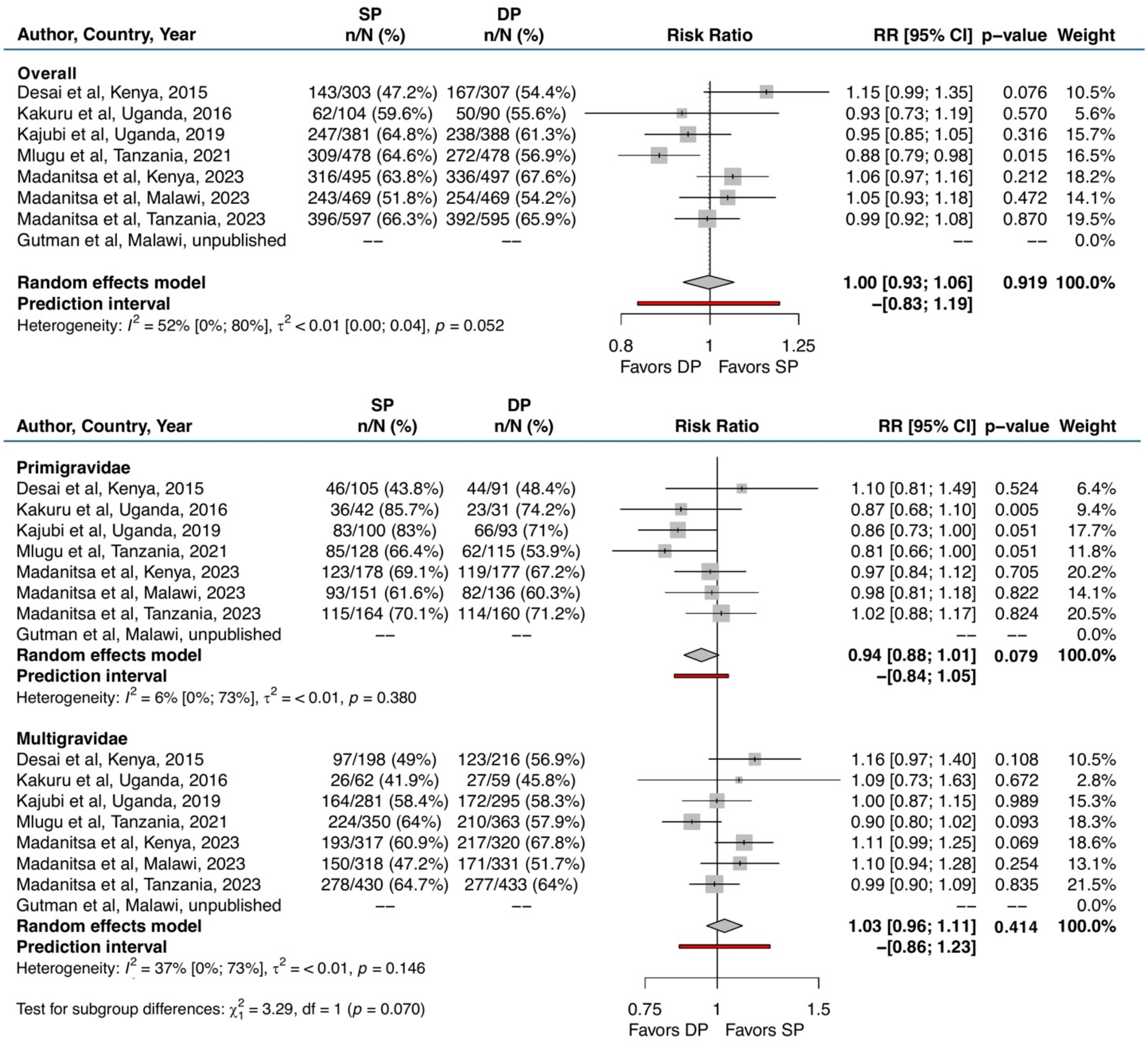

Abbreviations: CI = confidence interval; DP = dihydroartemisinin-piperaquine; g/dL = grams/deciliter; Hb = haemoglobin; RR = risk ratio; SP = sulfadoxine-pyrimethamine

Figure S-18. Mean maternal MUAC in cm at delivery

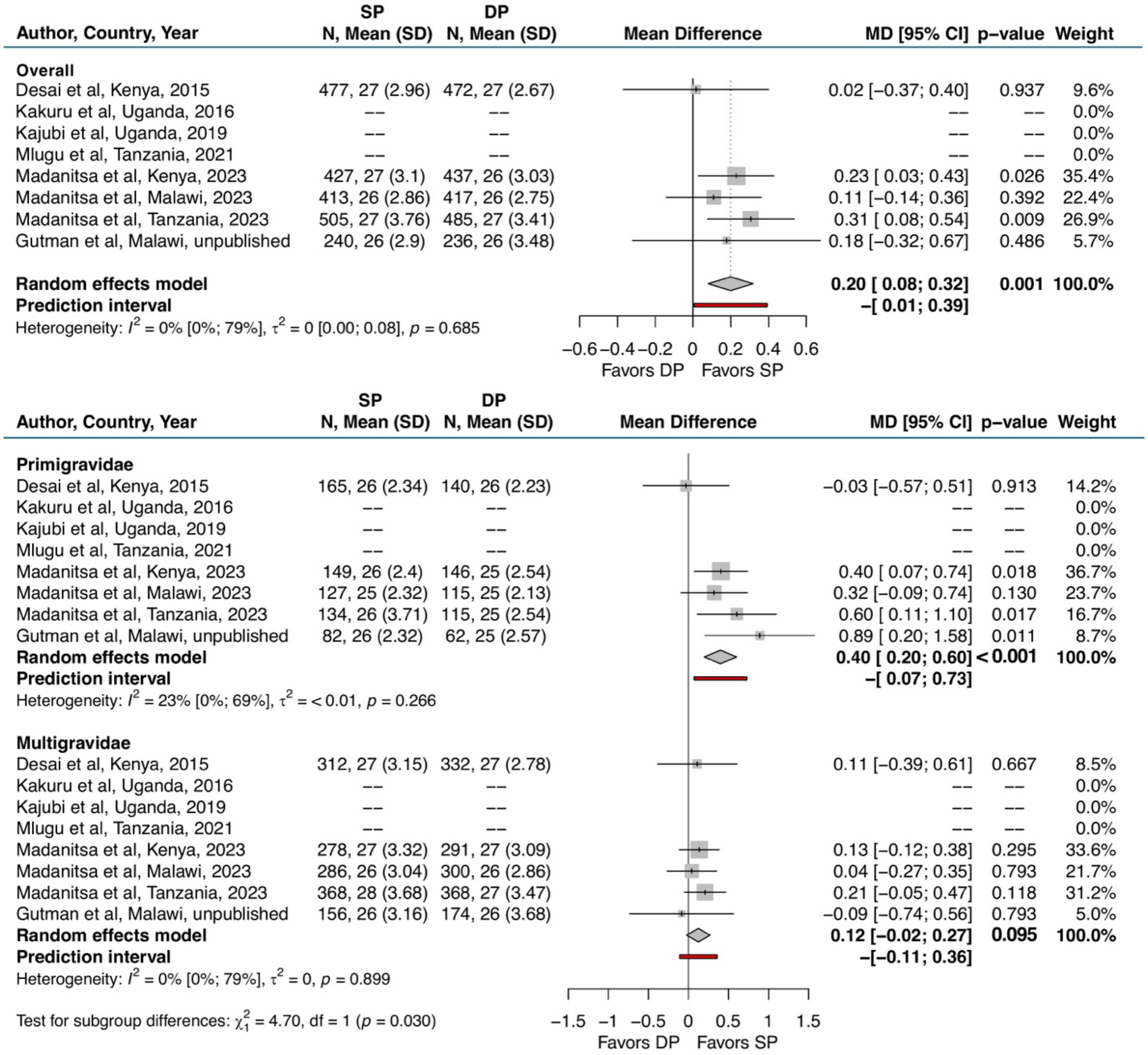

Abbreviations: CI = confidence interval; DP = dihydroartemisinin-piperaquine; MD = mean difference; MUAC = mid-upper arm circumference; SD = standard deviation; SP = sulfadoxine-pyrimethamine

### Figure S-19. Mean maternal weight gain per week in grams

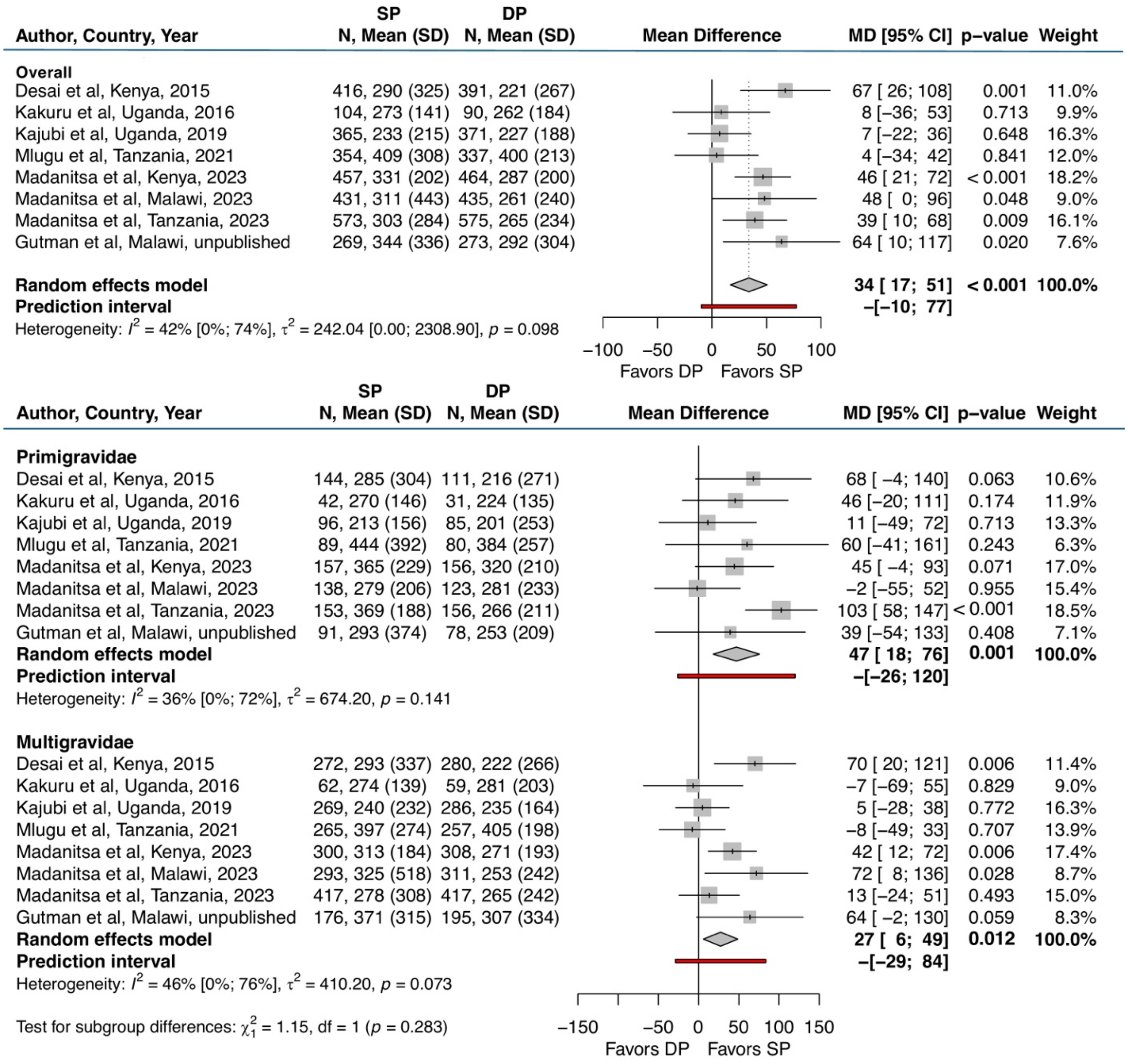

Abbreviations: CI = confidence interval; DP = dihydroartemisinin-piperaquine; MD = mean difference; SD = standard deviation; SP = sulfadoxine-pyrimethamine

### Figure S-20. Any evidence of stunting (LAZ <2 SD) from birth to two months of life

| Author, Country, Year | SP n/N (%) | DP n/N (%) | RR [95% CI] | p–value | Weight |
|---|---|---|---|---|---|
| **Overall** | | | | | |
| Desai et al, Kenya, 2015 | 127/361 (35.2%) | 142/349 (40.7%) | 1.16 [0.96; 1.40] | 0.131 | 20.8% |
| Kakuru et al, Uganda, 2016 | 23/94 (24.5%) | 18/82 (22%) | 0.90 [0.52; 1.54] | 0.694 | 3.4% |
| Kajubi et al, Uganda, 2019 | 77/304 (25.3%) | 110/322 (34.2%) | 1.35 [1.06; 1.72] | 0.017 | 14.0% |
| Mlugu et al, Tanzania, 2021 | –– | –– | –– | –– | 0.0% |
| Madanitsa et al, Kenya, 2023 | 43/408 (10.5%) | 55/427 (12.9%) | 1.22 [0.84; 1.78] | 0.295 | 6.7% |
| Madanitsa et al, Malawi, 2023 | 183/383 (47.8%) | 191/390 (49%) | 1.02 [0.89; 1.19] | 0.740 | 29.3% |
| Madanitsa et al, Tanzania, 2023 | 61/406 (15%) | 77/406 (19%) | 1.26 [0.93; 1.72] | 0.136 | 9.6% |
| Gutman et al, Malawi, unpublished | 71/162 (43.8%) | 87/155 (56.1%) | 1.28 [1.02; 1.60] | 0.030 | 16.2% |
| **Random effects model** | | | **1.16 [1.05; 1.29]** | **0.004** | **100.0%** |
| **Prediction interval** | | | **–[0.95; 1.43]** | | |

Heterogeneity: $I^2$ = 3% [0%; 72%], $\tau^2$ < 0.01 [0.00; 0.06], $p$ = 0.403

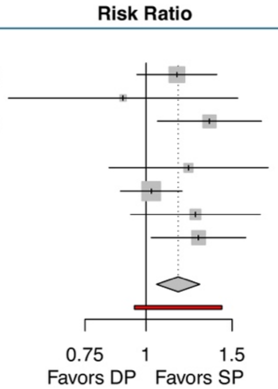

| Author, Country, Year | SP n/N (%) | DP n/N (%) | RR [95% CI] | p–value | Weight |
|---|---|---|---|---|---|
| **Primigravidae** | | | | | |
| Desai et al, Kenya, 2015 | 51/121 (42.1%) | 46/98 (46.9%) | 1.08 [0.73; 1.61] | 0.697 | 22.8% |
| Kakuru et al, Uganda, 2016 | 11/35 (31.4%) | 9/28 (32.1%) | 1.02 [0.42; 2.47] | 0.961 | 4.7% |
| Kajubi et al, Uganda, 2019 | 31/77 (40.3%) | 34/69 (49.3%) | 1.23 [0.76; 2.01] | 0.397 | 15.3% |
| Mlugu et al, Tanzania, 2021 | –– | –– | –– | –– | 0.0% |
| Madanitsa et al, Kenya, 2023 | 16/136 (11.8%) | 24/142 (16.9%) | 1.45 [0.77; 2.72] | 0.248 | 9.0% |
| Madanitsa et al, Malawi, 2023 | 69/118 (58.5%) | 51/109 (46.8%) | 0.79 [0.55; 1.13] | 0.197 | 27.6% |
| Madanitsa et al, Tanzania, 2023 | 18/98 (18.4%) | 18/91 (19.8%) | 1.07 [0.56; 2.06] | 0.838 | 8.5% |
| Gutman et al, Malawi, unpublished | 27/49 (55.1%) | 25/47 (53.2%) | 0.94 [0.55; 1.63] | 0.837 | 12.2% |
| **Random effects model** | | | **1.02 [0.84; 1.23]** | **0.853** | **100.0%** |
| **Prediction interval** | | | **–[0.79; 1.31]** | | |
| Heterogeneity: $I^2$ = 0% [0%; 71%], $\tau^2$ = 0, $p$ = 0.693 | | | | | |
| **Multigravidae** | | | | | |
| Desai et al, Kenya, 2015 | 76/240 (31.7%) | 96/251 (38.2%) | 1.19 [0.88; 1.61] | 0.251 | 20.8% |
| Kakuru et al, Uganda, 2016 | 12/59 (20.3%) | 9/54 (16.7%) | 0.82 [0.35; 1.95] | 0.653 | 2.5% |
| Kajubi et al, Uganda, 2019 | 46/227 (20.3%) | 76/253 (30%) | 1.49 [1.03; 2.15] | 0.031 | 14.1% |
| Mlugu et al, Tanzania, 2021 | –– | –– | –– | –– | 0.0% |
| Madanitsa et al, Kenya, 2023 | 27/272 (9.9%) | 31/285 (10.9%) | 1.10 [0.66; 1.84] | 0.716 | 7.1% |
| Madanitsa et al, Malawi, 2023 | 114/265 (43%) | 139/279 (49.8%) | 1.17 [0.92; 1.51] | 0.201 | 30.7% |
| Madanitsa et al, Tanzania, 2023 | 43/305 (14.1%) | 59/313 (18.8%) | 1.31 [0.89; 1.95] | 0.170 | 12.2% |
| Gutman et al, Malawi, unpublished | 44/111 (39.6%) | 62/108 (57.4%) | 1.44 [0.98; 2.12] | 0.063 | 12.6% |
| **Random effects model** | | | **1.25 [1.09; 1.43]** | **0.002** | **100.0%** |
| **Prediction interval** | | | **–[1.04; 1.50]** | | |

Heterogeneity: $I^2$ = 0% [0%; 71%], $\tau^2$ = 0, $p$ = 0.819

Test for subgroup differences: $\chi^2_1$ = 2.93, df = 1 ($p$ = 0.087)

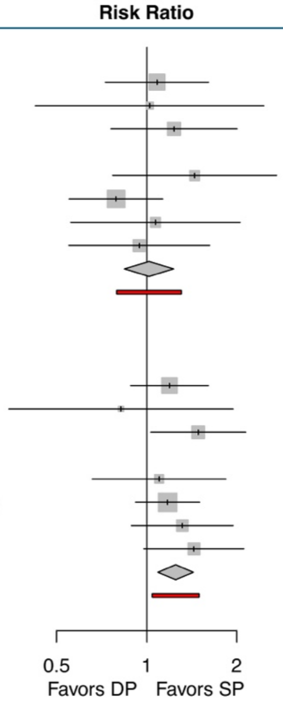

Abbreviations: CI = confidence interval; DP = dihydroartemisinin-piperaquine; LAZ = length-for-age z-score; RR = risk ratio; SD = standard deviation; SP = sulfadoxine-pyrimethamine

Figure S-21. Any evidence of underweight (WAZ <2 SD) from birth to two months of life

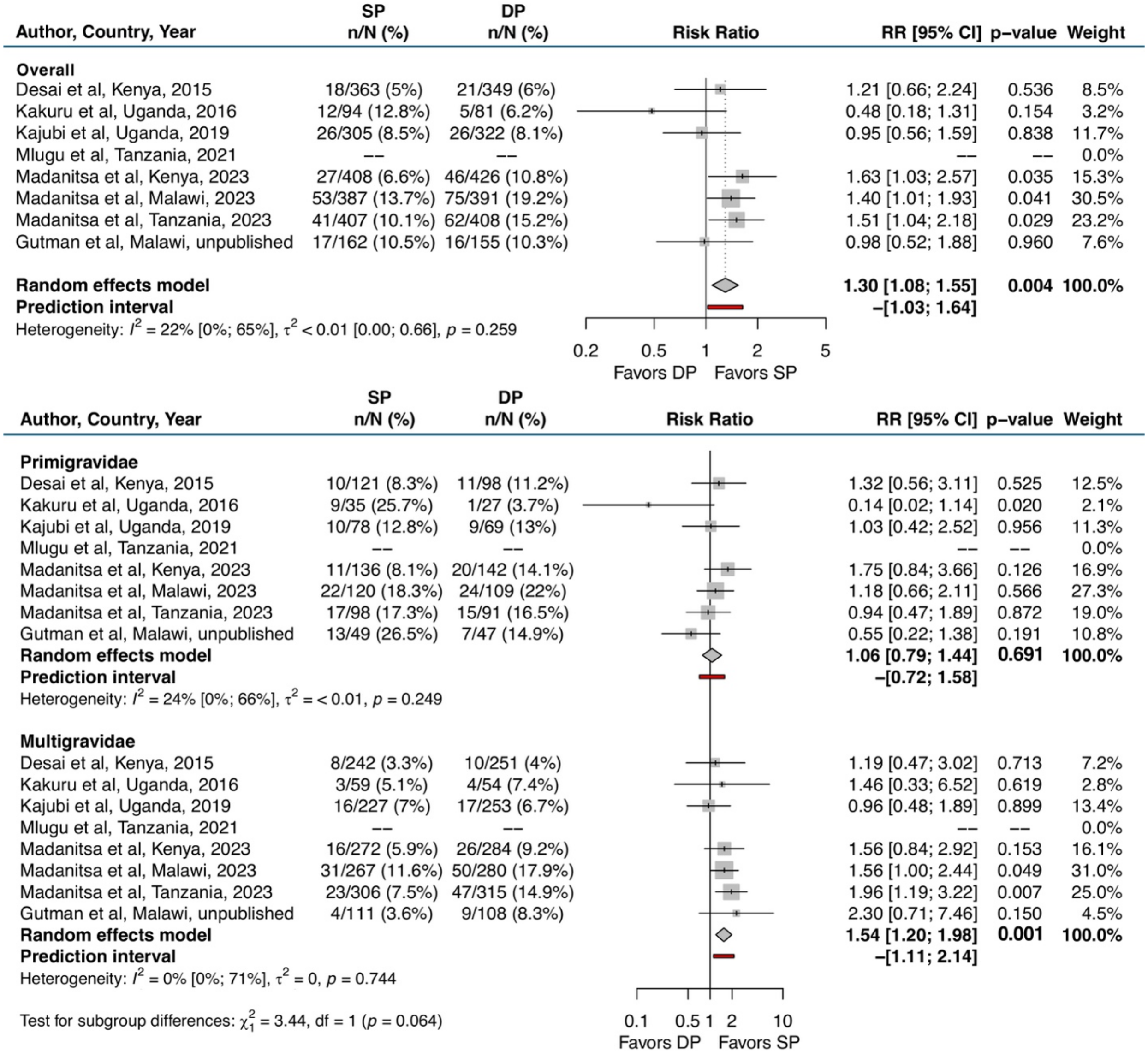

Abbreviations: CI = confidence interval; DP = dihydroartemisinin-piperaquine; RR = risk ratio; SD = standard deviation; SP = sulfadoxine-pyrimethamine; WAZ = weight-for-age z-score

Figure S-22. Any evidence of wasting (WLZ <2 SD) from birth to two months of life

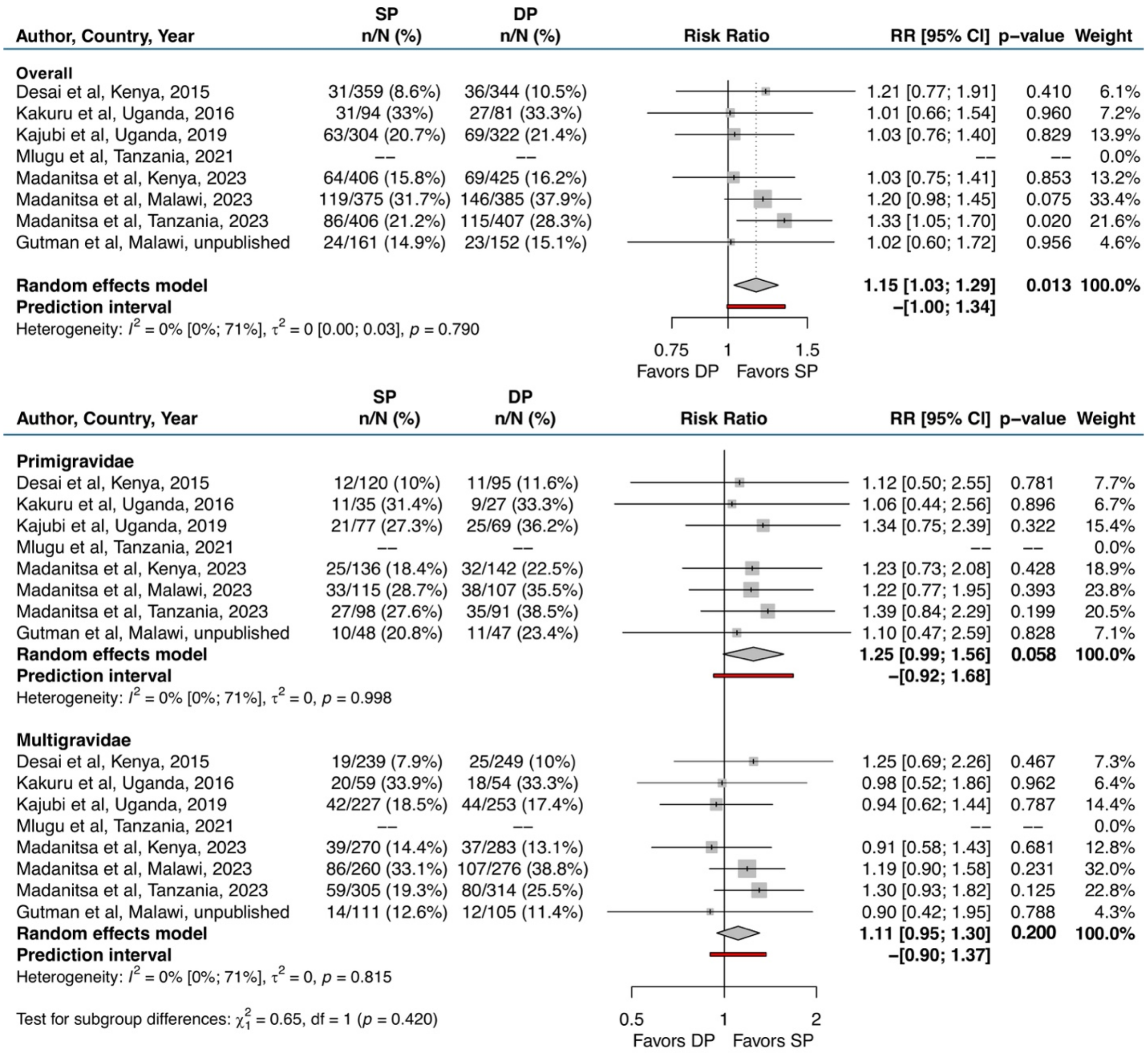

Abbreviations: CI = confidence interval; DP = dihydroartemisinin-piperaquine; RR = risk ratio; SD = standard deviation; SP = sulfadoxine-pyrimethamine; WLZ = weight-for-length z-score

#### Figure S-23. Mean length-for-age z-score at two months of life

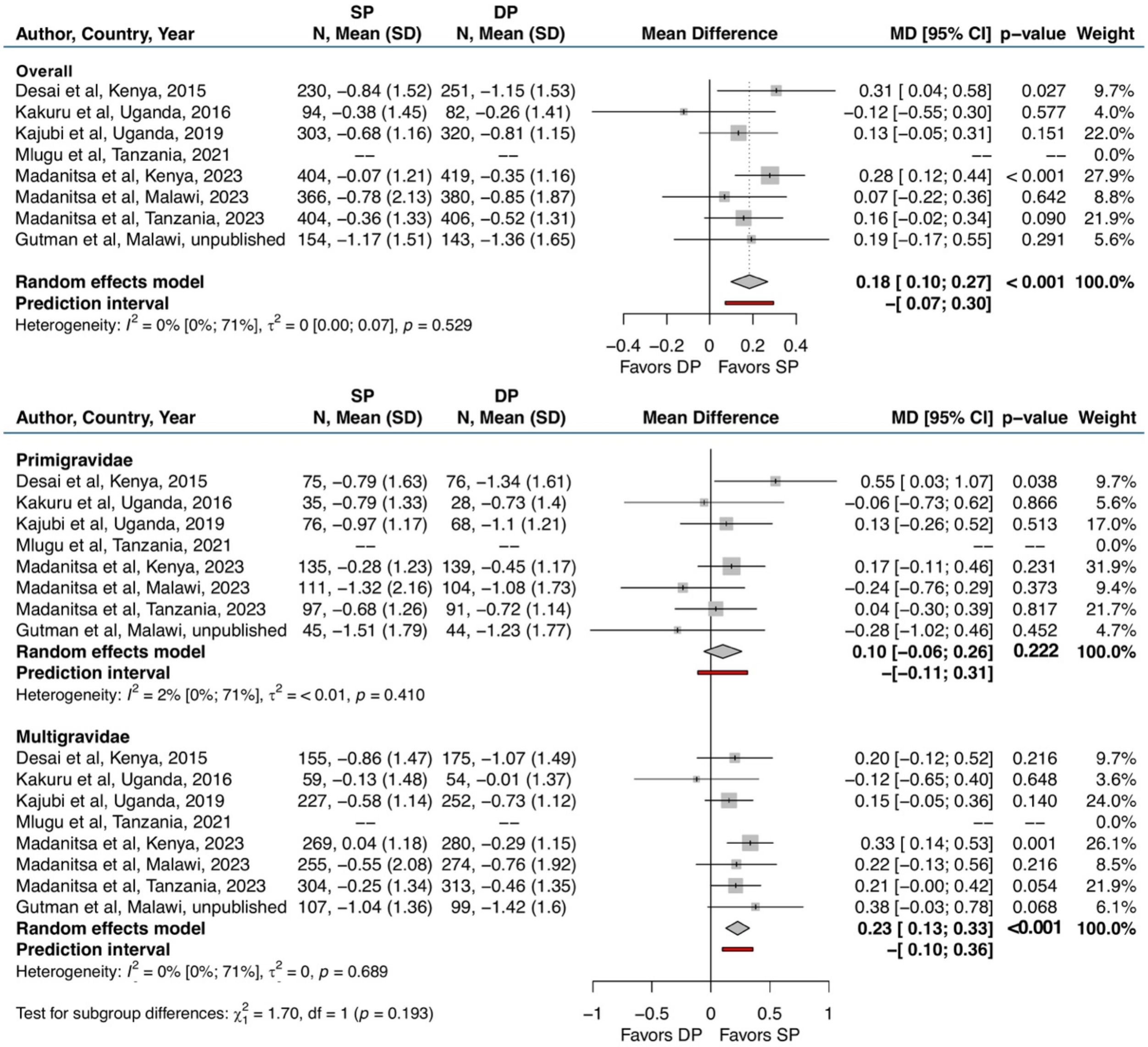

Abbreviations: CI = confidence interval; DP = dihydroartemisinin-piperaquine; MD = mean difference; SD = standard deviation; SP = sulfadoxine-pyrimethamine

### Figure S-24. Mean weight-for-age z-score at two months of life

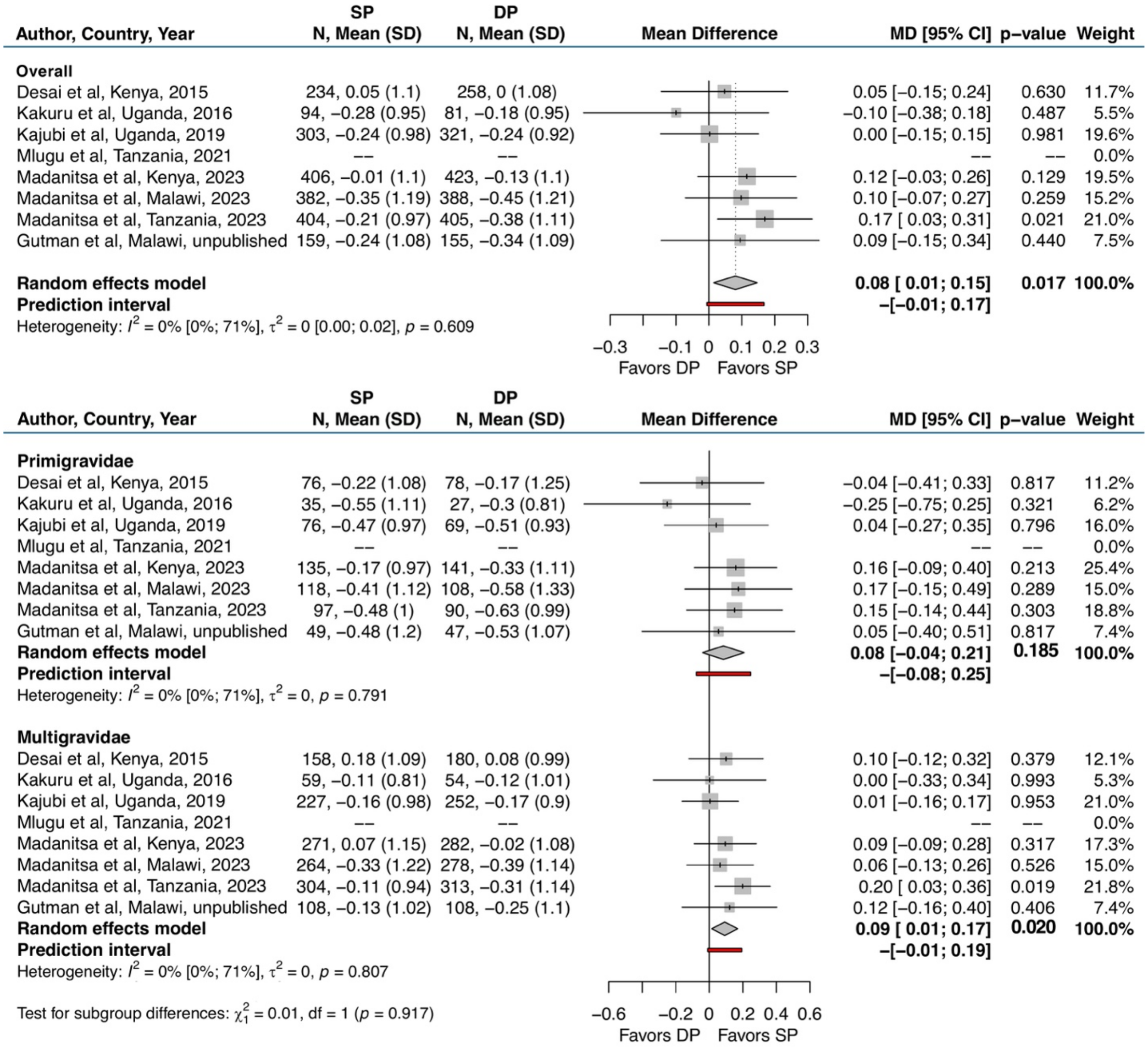

Abbreviations: CI = confidence interval; DP = dihydroartemisinin-piperaquine; MD = mean difference; SD = standard deviation; SP = sulfadoxine-pyrimethamine

### Figure S-25. Mean weight-for-length z-score at two months of life

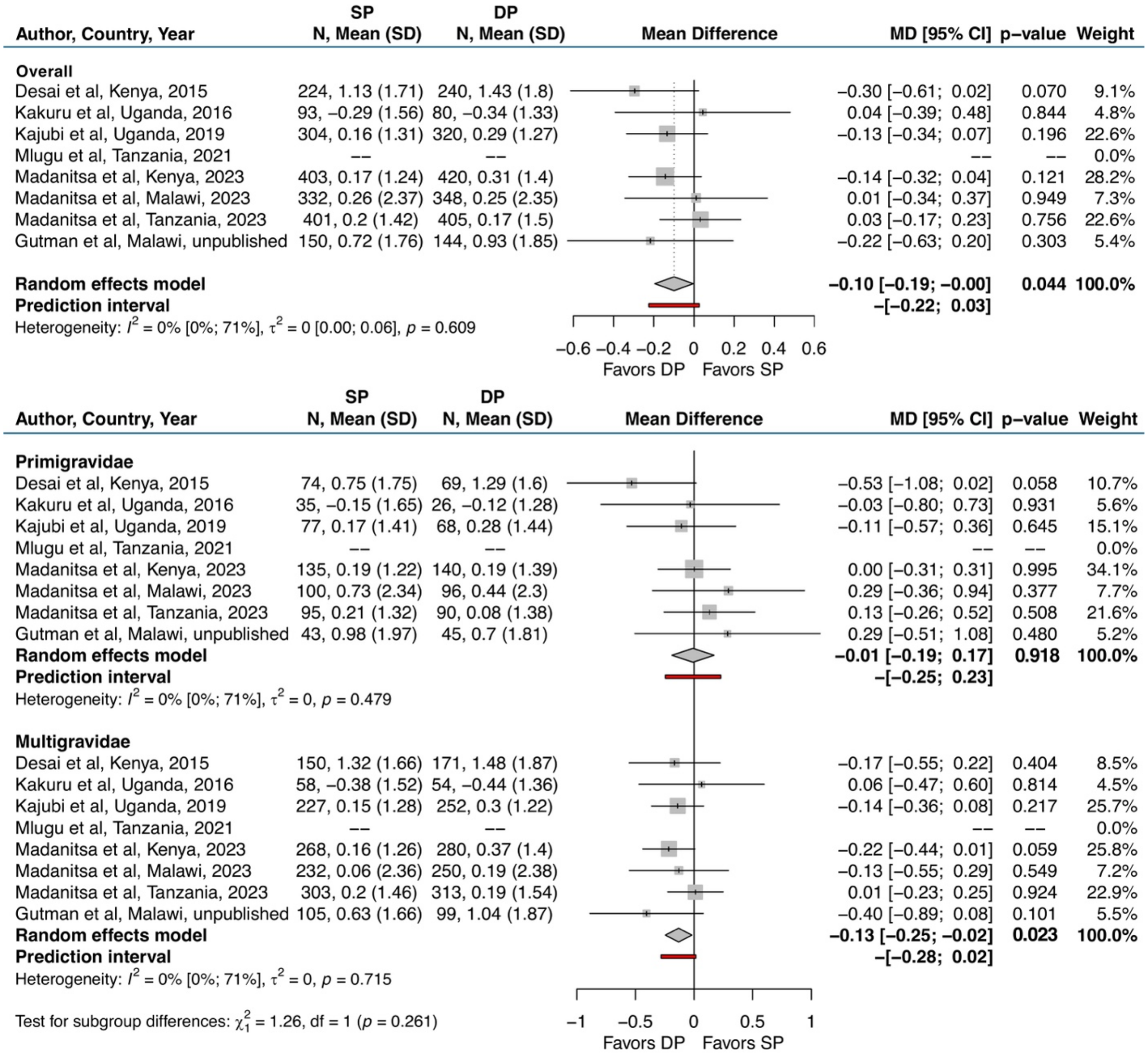

Abbreviations: CI = confidence interval; DP = dihydroartemisinin-piperaquine; MD = mean difference; SD = standard deviation; SP = sulfadoxine-pyrimethamine

### Appendix 8. Study-specific effect estimates from mediation analyses

Figure S-26. IPTp differences in BWGA z-scores mediated by incidence of clinical malaria during pregnancy

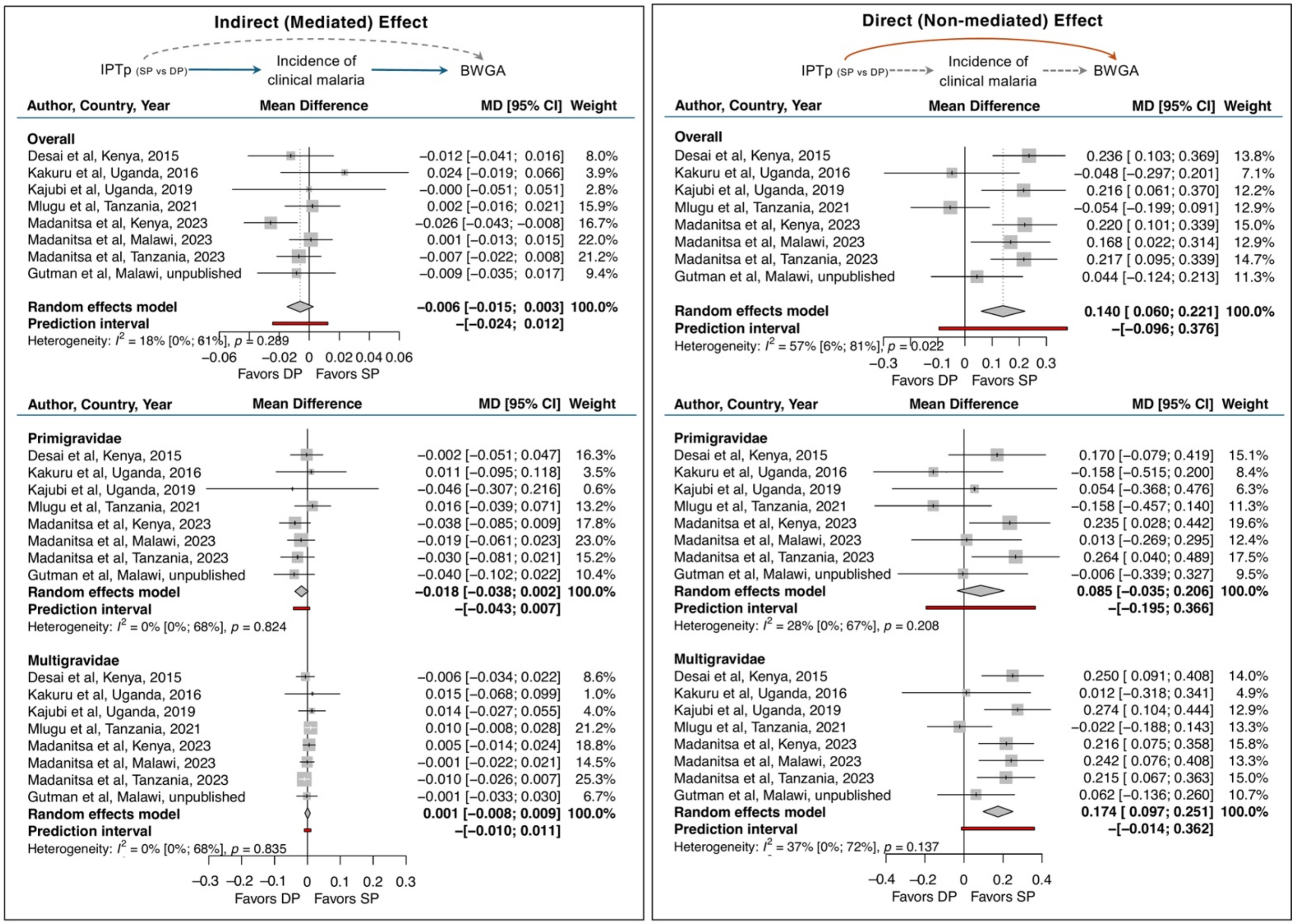

Abbreviations: BWGAz = Birthweight-for-gestational age, CI = confidence interval; DP = dihydroartemisinin-piperaquine; MD = mean difference; SP = sulfadoxine-pyrimethamine

Figure S-27. IPTp differences in BWGA z-scores mediated by placental malaria infection defined as any evidence of pigment or parasites detected by histopathology, microscopy, PCR, or RDT

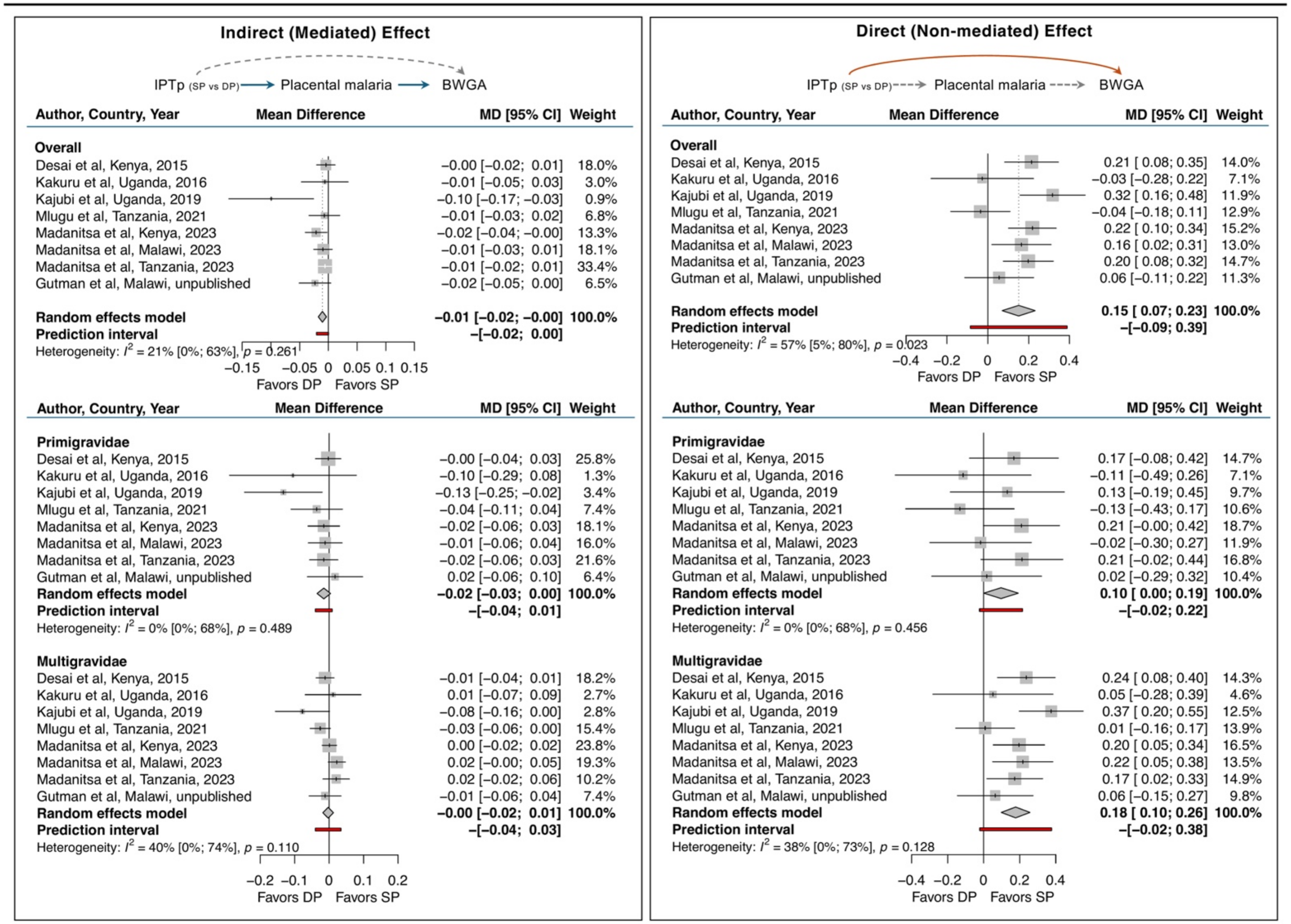

Abbreviations: BWGAz = birthweight-for-gestational age z-score; CI = confidence interval; DP = dihydroartemisinin-piperaquine; MD = mean difference; SP = sulfadoxine-pyrimethamine

#### Figure S-28. IPTp differences in BWGA z-scores mediated by gestational weight gain

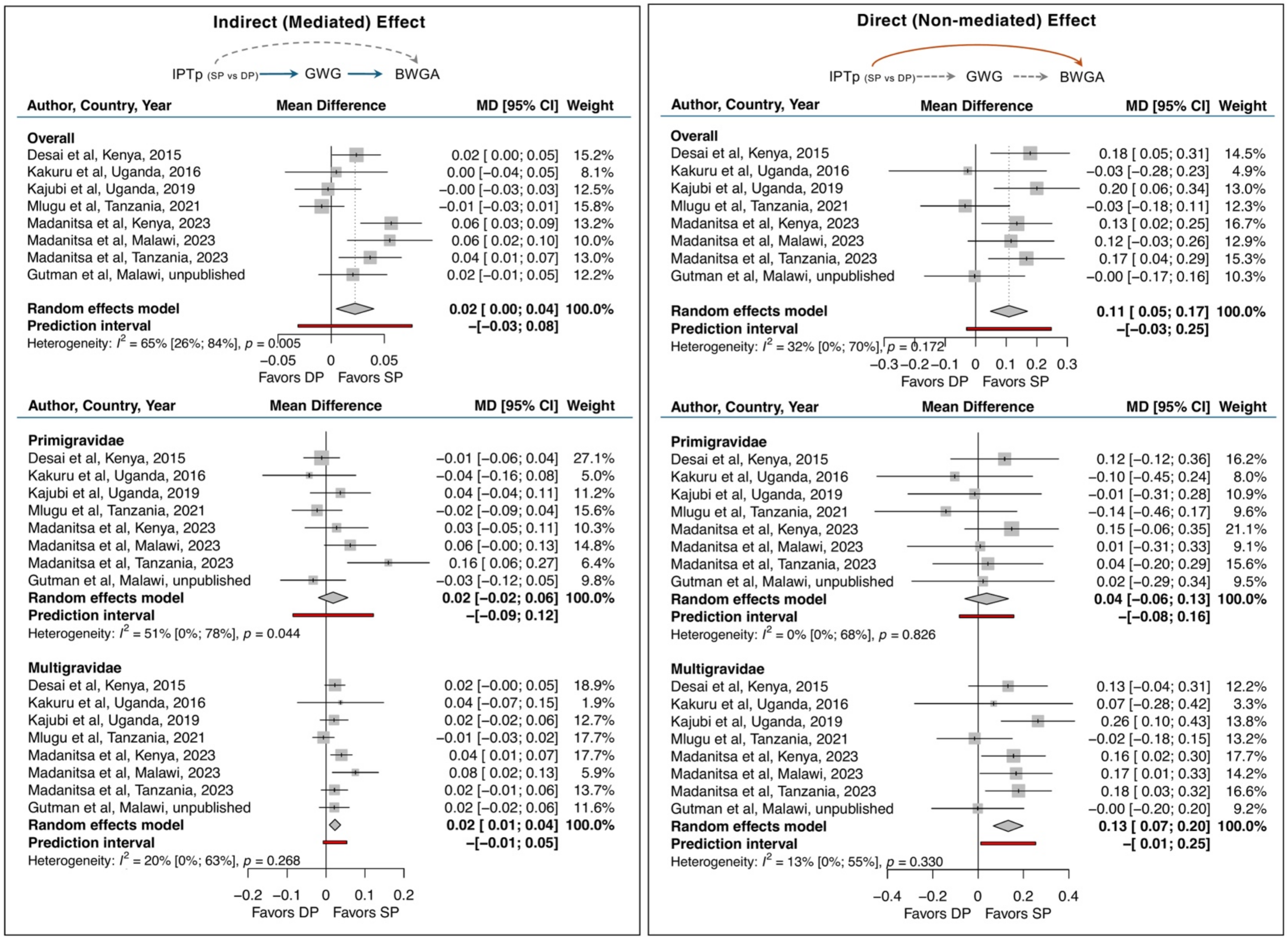

Abbreviations: BWGAz = Birthweight-for-gestational age, CI = confidence interval; DP = dihydroartemisinin-piperaquine; GWG = Gestational weight gain; MD = mean difference; SP = sulfadoxine-pyrimethamine

#### Figure S-29. IPTp differences in BWGA z-scores mediated by MUAC

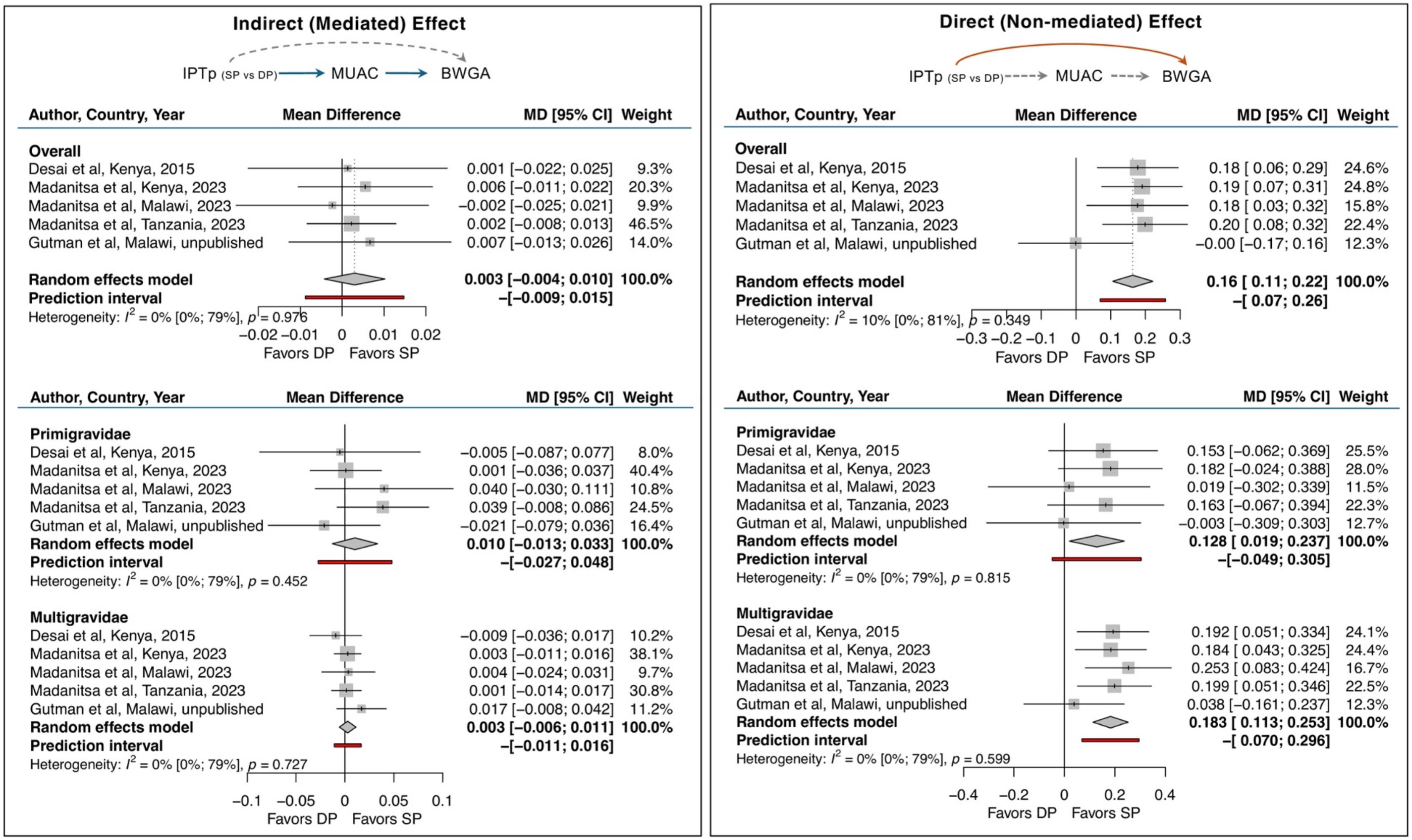

Abbreviations: BWGAz = Birthweight-for-gestational age, CI = confidence interval; DP = dihydroartemisinin-piperaquine; MD = mean difference; MUAC = mid-upper arm circumference; SP = sulfadoxine-pyrimethamine

### Appendix 9. Results for excluded Okoro et al, Nigeria (2023) trial

#### Table S-9. Table 1 Participant characteristics

| Enrolment characteristics | Total<br>N=197 | IPTp-SP<br>N=91 | IPTp-DP<br>N=106 |
|---|---|---|---|
| **Maternal age in years, mean (SD)** | 27·4 (6·0) | 27·7 (6·3) | 27·0 (5·8) |
| **Gestational age in weeks, mean (SD)** | 17·5 (1·6) | 17·7 (1·6) | 17·3 (1·5) |
| **Gravidity categories, n/N (%)** | | | |
| Primigravidae | 68/197 (35%) | 30/91(33%) | 38/106 (36%) |
| Secundigravidae | 25/197 (13%) | 11/91 (12%) | 14/106 (13%) |
| Multigravidae (3+) | 104/194 (53%) | 50/91 (55%) | 54/106 (51%) |
| **Weight in kg, mean (SD)** | 65·0 (15·6) | 66·1 (14·9) | 64·0 (16·1) |
| **Height in cm, mean (SD)** | 158·7 (7·8) | 159·0 (6·7) | 158·3 (8·6) |
| **Maternal MUAC in cm, mean (SD)** | -- | -- | -- |
| **Highest level of schooling completed, n/N (%)** | | | |
| None | -- | -- | -- |
| Primary | -- | -- | -- |
| Secondary | -- | -- | -- |
| Higher | -- | -- | -- |
| **Wealth index tertiles, n/N (%)** | | | |
| Lowest tertile | -- | -- | -- |
| Middle tertile | -- | -- | -- |
| Highest tertile | -- | -- | -- |
| **Slept under a bed net last night, n/N (%)** | -- | -- | -- |
| **Microscopy positivity, n/N (%)** | 82/196 (42%) | 41/91 (48%) | 39/105 (37%) |
| **PCR/LAMP positivity, n/N (%)** | -- | -- | -- |

Abbreviations: LAMP = loop-mediated isothermal amplification method MUAC = mid-upper arm circumference; PCR = polymerase chain reaction

Table S-9. Description of study characteristics and outcomes collected

| Study Information | Okoro et al[22] |
|---|---|
| **Study Details** | |
| Source | PACTR202002644579177 |
| Study site(s) | Tertiary hospital in Maiduguri, Nigeria |
| Prevalence of PfDHPS 540E mutation, %* | 0% |
| Prevalence of PfDHPS 581G mutation, %* | 0% |
| Number of Participants Randomized (Among Singleton Pregnancies) | 197 |
| Sulfadoxine-Pyrimethamine | 91 |
| Dihydroartemisinin-Piperaquine | 106 |
| IPTp dosing regimen | 3-course IPTp given at 16-20, 28, and 36 weeks |
| Number of IPTp doses, median (IQR) | 2 (2-3) |
| Microscopy positivity at enrolment, % | 42% |
| **Birth outcomes** | |
| Foetal Loss | Available (nne observed) |
| Small-for-Gestational Age | Infant sex not collected |
| Preterm Delivery | Available[†] |
| Low Birthweight | Available |
| Neonatal Death | Available (none observed) |
| **Continuous Birth Outcomes** | |
| Mean Birthweight | Available |
| Mean Gestational Age at Delivery | Available[†] |
| Mean Birthweight-for-Gestational Age Z-scores | Infant sex not collected |
| **Malaria Outcomes** | |
| Incidence of Clinical Malaria Episodes in Pregnancy | Not collected |
| Any Evidence of Pigment Only in Placental Tissue by Histopathology | Available |
| Any Evidence of Parasites in Placental Tissue or Blood by Histopathology, PCR, Microscopy, or RDT | Available; Testing of placental blood for parasitemia not done |
| Any Evidence of Parasites or Pigment in Placental Tissue or Blood by Histopathology, PCR, Microscopy, or RDT | Available; Testing of placental blood for parasitemia not done |
| Any Evidence of Parasites in Maternal Peripheral Blood at Delivery by RDT, Microscopy, or PCR | Available; Testing by microscopy only |
| **Maternal Outcomes** | |
| Any Evidence of Severe Anaemia (Hb <7 g/dl) During Pregnancy | Not collected |

| Study Information | Okoro et al[22] |
|---|---|
| Any Evidence of Moderate Anaemia (Hb <9 g/dl) During Pregnancy | Not collected |
| Any Evidence of Mild Anaemia (Hb <11 g/dl) During Pregnancy | Not collected |
| MUAC at Delivery | Not collected |
| Maternal weight gain per week‡ | Not collected |
| **Infant Outcomes** | |
| Any Evidence of Stunting (LAZ <2 SD) from Birth to 2 Months of Life | Not collected |
| Any Evidence of Underweight (WAZ <2 SD) from Birth to 2 Months of Life | Not collected |
| Any Evidence of Wasting (WLZ <2 SD) from Birth to 2 Months of Life | Not collected |
| Mean LAZ at 2 Months of Life | Not collected |
| Mean WAZ at 2 Months of Life | Not collected |
| Mean WLZ at 2 Months of Life | Not collected |

Abbreviations: ANC = antenatal care visit; Hb = haemoglobin; IQR = interquartile range; LAZ = length-for-age z-score; MUAC = mid-upper arm circumference; WAZ = weight-for-age z-score; WLZ = weight-for-length z-score

* Prevalence of polymorphisms were taken from another study from a neighboring site.[23]

† Gestational age dating not confirmed by ultrasound

‡ Maternal weight gain per week calculated using the following formula:

$$\frac{Weight_{last\ ANC\ visit\ before\ delivery} - Weight_{enrollment}}{\#\ of\ weeks\ between\ enrollment\ and\ last\ ANC\ visit}$$

### Figure S-30. Birth outcomes

#### Results from Okoro et al, 2023 trial[22]

##### Binary Birth Outcomes

| Birth Outcomes | Prevalence, n/N (%) DP | Prevalence, n/N (%) SP | DP:SP Summary Estimate RR [95% CI] | p-value | gravidity $p_{interaction}$ |
|---|---|---|---|---|---|
| Any composite adverse pregnancy outcome (foetal loss, preterm birth, SGA, LBW, neonatal death) | 9/106 (8·5%) | 12/91 (13·2%) | 0·64 [0·28, 1·46] | 0·29 | 0·83 |
| | 2/38 (5·3%) | 2/30 (6·7%) | 0·79 [0·12, 5·28] | 0·81 | |
| | 7/68 (10·3%) | 10/61 16·4%) | 0·63 [0·25, 1·55] | 0·31 | |
| Foetal loss (miscarriage, abortion, or stillbirth) | 0/106 (0%) | 0/91 (0%) | -- | -- | |
| | 0/38 (0%) | 0/30 (0%) | -- | -- | |
| | 0/68 (0%) | 0/61 (0%) | -- | -- | |
| Small-for-gestational age (<10th percentile for birthweight-for-gestational age)[1] | -- | -- | -- | -- | |
| | -- | -- | -- | -- | |
| | -- | -- | -- | -- | |
| Preterm Birth (<37 gestational weeks) | 4/106 (3·8%) | 5/91 (5·5%) | 0·69 [0·19, 2·48] | 0·57 | -- |
| | 0/38 (0%) | 0/30 (0%) | -- | -- | |
| | 4/68 (5·9%) | 5/61 (8·2%) | 0·72 [0·20, 2·55] | 0·61 | |
| Low Birthweight (<2500 grams) | 8/106 (7·5%) | 12/91 (13·2%) | 0·57 [0·24, 1·34] | 0·20 | 0·72 |
| | 2/38 (5·3%) | 2/30 (6·7%) | 0·79 [0·12, 5·28] | 0·81 | |
| | 6/68 (8·8%) | 10/61 (16·4%) | 0·54 [0·21, 1·39] | 0·20 | |
| Neonatal Death (death within the first 28 days of life) | 0/106 (0%) | 0/91 (0%) | -- | -- | |
| | 0/38 (0%) | 0/30 (0%) | -- | -- | |
| | 0/68 (0%) | 0/61 (0%) | -- | -- | |

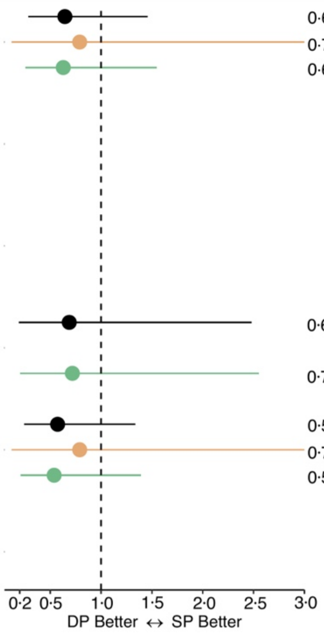

##### Continuous Birth Outcomes[2]

| Outcome | N, Mean (SD) DP | N, Mean (SD) SP | SP:DP Summary Estimate MD [95% CI] | p-value | gravidity $p_{interaction}$ |
|---|---|---|---|---|---|
| Mean newborn birthweight in grams | 106, 3078 (502) | 91, 3012 (589) | -66 [-218, 86] | 0·40 | 0·96 |
| | 38 (3137 (447) | 30, 3080 (381) | -57 [-258, 143] | 0·58 | |
| | 68, 3045 (531) | 61, 2978 (668) | -66 [-274, 141] | 0·53 | |
| Mean gestational age at birth in weeks | 106, 39 (1·6) | 91, 39 (1·4) | -0·08 [-0·51, 0·34] | 0·51 | 0·66 |
| | 38, 39 (1·4) | 30, 39 (0·9) | 0·09 [-0·48, 0·66] | 0·75 | |
| | 68, 39 (1·7) | 61, 39 (1·6) | -0·11 [-0·67, 0·46] | 0·71 | |

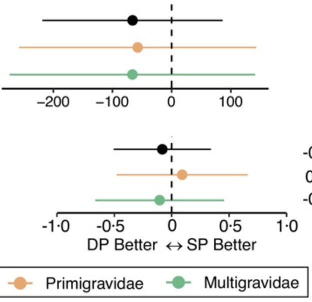

Abbreviations: CI=confidence interval; DP=dihydroartemisinin-piperaquine; MD=mean difference; RR = relative risk ratio; SD=standard deviation; SP=sulfadoxine-pyrimethamine

[1] Did not collect infant sex variable used to determine birthweight percentiles for gestational age

[2] Birthweight-for-gestational age z-scores missing due to unknown infant sex

### Figure S-31. Malaria Outcomes

#### Results from Okoro et al, 2023 trial[22]

**Malaria Outcomes**

| Malaria Outcomes | Prevalence, n/N (%) DP | Prevalence, n/N (%) SP | DP:SP Summary Estimate RR [95% CI] | DP:SP Summary Estimate p-value | gravidity $p_{interaction}$ |
|---|---|---|---|---|---|
| Incidence of clinical malaria episodes during pregnancy[1] | -- | -- | -- | -- | -- |
| | -- | -- | -- | -- | |
| | -- | -- | -- | -- | |
| Any evidence of pigment only in placental tissue by histopathology | 35/75 (49·3%) | 28/71 (39·4%) | 1·25 [0·87, 1·81] | 0·23 | 0·038 |
| | 16/27 (59·3%) | 6/24 (25%) | 2·37 [1·11, 5·07] | 0·026 | |
| | 21/48 (43·8%) | 22/47 (46·8%) | 0·93 [0·60, 1·45] | 0·77 | |
| Any evidence of parasites in placental tissue or blood by histopathology, PCR, microscopy, or RDT | 31/106 (29·2%) | 20/91 (22%) | 1·33 [0·82, 2·17] | 0·25 | 0·82 |
| | 11/38 (28·9%) | 6/30 (20%) | 1·45 [0·61, 3·46] | 0·41 | |
| | 20/68 (29·4%) | 14/61 (23%) | 1·28 [0·71, 2·31] | 0·41 | |
| Any evidence of parasites or pigment in placental tissue or blood by histopathology, PCR, microscopy, or RDT | 68/106 (64·2%) | 48/91 (52·7%) | 1·22 [0·96, 1·55] | 0·11 | 0·053 |
| | 27/38 (71·1%) | 12/30 (40%) | 1·78 [1·10, 2·88] | 0·020 | |
| | 41/68 (60·3%) | 36/61 (59%) | 1·02 [0·77, 1·36] | 0·88 | |
| Any evidence of parasites in maternal peripheral blood at delivery by RDT, microscopy, or PCR | 81/106 (76·4%) | 61/91 (67%) | 1·14 [0·95, 1·36] | 0·15 | 0·087 |
| | 27/38 (71·1%) | 23/30 (76·7%) | 0·93 [0·70, 1·23] | 0·60 | |
| | 54/68 (79·4%) | 38/61 (62·3%) | 1·27 [1·01, 1·60] | 0·038 | |

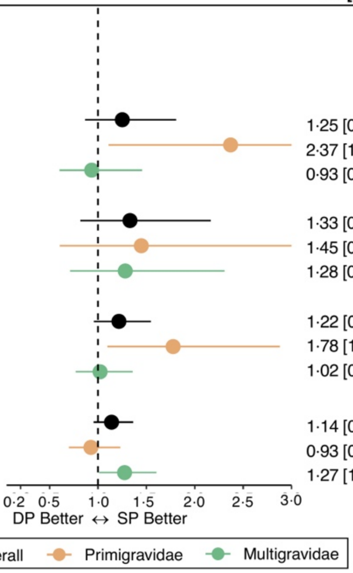

Abbreviations: CI=confidence interval; DP=dihydroartemisinin-piperaquine; PCR=polymerase chain reaction; RDT=rapid diagnostic test; RR=relative risk ratio; SP=sulfadoxine-pyrimethamine

[1] Outcome not collected

### Appendix 10. Appendix References

1. Desai M, Gutman J, L'lanziva A, et al. Intermittent screening and treatment or intermittent preventive treatment with dihydroartemisinin–piperaquine versus intermittent preventive treatment with sulfadoxine–pyrimethamine for the control of malaria during pregnancy in western Kenya: an open-label, three-group, randomised controlled superiority trial. *Lancet* 2015; **386**(10012): 2507-19.
2. Kakuru A, Jagannathan P, Muhindo MK, et al. Dihydroartemisinin–piperaquine for the prevention of malaria in pregnancy. *N Engl J Med* 2016; **374**(10): 928-39.
3. Kajubi R, Ochieng T, Kakuru A, et al. Monthly sulfadoxine-pyrimethamine versus dihydroartemisinin-piperaquine for intermittent preventive treatment of malaria in pregnancy: A randomized controlled trial. *Lancet* 2019.
4. Mlugu EM, Minzi O, Kamuhabwa AA, Aklillu E. Effectiveness of intermittent preventive treatment with dihydroartemisinin-piperaqunine against malaria in pregnancy in Tanzania: A Randomized Controlled Trial. *Clin Pharmacol Ther* 2021.
5. Madanitsa M, Barsosio HC, Minja DT, et al. Effect of monthly intermittent preventive treatment with dihydroartemisinin–piperaquine with and without azithromycin versus monthly sulfadoxine–pyrimethamine on adverse pregnancy outcomes in Africa: a double-blind randomised, partly placebo-controlled trial. *Lancet* 2023; **401**(10381): 1020-36.
6. Gutman J. A Prospective Randomized Open-Label Study on the Efficacy and Safety of Intermittent Preventive Treatment in Pregnancy (IPTp) With Dihydroartemisinin-Piperaquine (DP) Versus IPTp With Sulfadoxine-Pyrimethamine (SP) in Malawi. 2016. Available from https://clinicaltrials.gov/study/NCT03009526 ClinicalTrials.gov Identifer: NCT03009526.
7. Conrad MD, Mota D, Foster M, et al. Impact of Intermittent Preventive Treatment During Pregnancy on Plasmodium falciparum Drug Resistance–Mediating Polymorphisms in Uganda. *J Infect Dis* 2017; **216**(8): 1008-17.
8. Nayebare P, Asua V, Conrad MD, et al. Associations between malaria-preventive regimens and Plasmodium falciparum drug resistance-mediating polymorphisms in Ugandan pregnant women. *Antimicrob Agents Chemother* 2020; **64**(12): 10.1128/aac. 01047-20.
9. Gutman J, Kalilani L, Taylor S, et al. The A581 G mutation in the gene encoding Plasmodium falciparum dihydropteroate synthetase reduces the effectiveness of sulfadoxine-pyrimethamine preventive therapy in Malawian pregnant women. *J Infect Dis* 2015; **211**(12): 1997-2005.
10. Villar J, Ismail LC, Victora CG, et al. International standards for newborn weight, length, and head circumference by gestational age and sex: the Newborn Cross-Sectional Study of the INTERGROWTH-21st Project. *The Lancet* 2014; **384**(9946): 857-68.
11. Rogerson SJ, Hviid L, Duffy PE, Leke RF, Taylor DW. Malaria in pregnancy: pathogenesis and immunity. *Lancet Infect Dis* 2007; **7**(2): 105-17.
12. WHO Multicentre Growth Reference Study Group, de Onis M. WHO Child Growth Standards based on length/height, weight and age. *Acta Paediatr* 2006; **95**: 76-85.

13. Rubin DB. Estimating causal effects of treatments in randomized and nonrandomized studies. *J Educ Psychol* 1974; **66**(5): 688.
14. Pearl J. The causal mediation formula—a guide to the assessment of pathways and mechanisms. *Prev Sci* 2012; **13**(4): 426-36.
15. Pearl J. Direct and indirect effects. Proceedings of the seventeenth conference on uncertainty in artificial intelligence; 2001: Morgan Kaufmann Publishers Inc.; 2001. p. 411-20.
16. Imai K, Keele L, Yamamoto T. Identification, inference and sensitivity analysis for causal mediation effects. *Stat Sci* 2010: 51-71.
17. Rudolph KE, Williams NT, Diaz I. Practical causal mediation analysis: extending nonparametric estimators to accommodate multiple mediators and multiple intermediate confounders. *Biostatistics* 2024: kxae012.
18. Coyle JR, Hejazi NS, Malenica I, Phillips RV, Sofrygin O. Modern Pipelines for Machine Learning and Super Learning. In: https://github.com/tlverse/sl3, editor. R package version 1.4.2 ed; 2021.
19. Zheng W, Van Der Laan MJ. Targeted maximum likelihood estimation of natural direct effects. *The international journal of biostatistics* 2012; **8**(1): 1-40.
20. Hejazi N, Rudolph K, Díaz I. medoutcon: Nonparametric efficient causal mediation analysis with machine learning in R. *JOSS* 2022; **7**(69).
21. McGuinness LA, Higgins JP. Risk-of-bias VISualization (robvis): an R package and Shiny web app for visualizing risk-of-bias assessments. *Res Syn Meth* 2021; **12**(1): 55-61.
22. Okoro RN, Geidam AD, Bukar AA, et al. Superiority trial of intermittent treatment with dihydroartemisinin–piperaquine versus sulfadoxine–pyrimethamine for the prevention of malaria during pregnancy. *Futur J Pharm Sci* 2023; **9**(1): 8.
23. Balogun ST, Sandabe UK, Sodipo OA, Okon KO, Akanmu AO. Single nucleotide polymorphisms of Pfdhfr and Pfdhps genes: implications for malaria prophylactic strategies in Maiduguri, Northeast Nigeria. *J Trop Med* 2021; **2021**: 1-7.
